# Superior antibody and membrane protein-specific T cell responses to CoronaVac by intradermal versus intramuscular routes in adolescents

**DOI:** 10.1101/2023.04.05.23288005

**Authors:** Jaime S. Rosa Duque, Samuel M.S. Cheng, Carolyn A. Cohen, Daniel Leung, Xiwei Wang, Xiaofeng Mu, Yuet Chung, Tsun Ming Lau, Manni Wang, Wenyue Zhang, Yanmei Zhang, Howard H.W. Wong, Leo C.H. Tsang, Sara Chaothai, Tsz Chun Kwan, John K.C. Li, Karl C.K. Chan, Leo L.H. Luk, Jenson C.H. Ho, Wing Yan Li, Amos M.T. Lee, Jennifer H.Y. Lam, Sau Man Chan, Wilfred H.S. Wong, Issan Y.S. Tam, Masashi Mori, Sophie A. Valkenburg, Malik Peiris, Wenwei Tu, Yu Lung Lau

**Affiliations:** Department of Paediatrics and Adolescent Medicine, The University of Hong Kong, Hong Kong, China; School of Public Health, The University of Hong Kong, Hong Kong, China; HKU-Pasteur Research Pole, School of Public Health, The University of Hong Kong, Hong Kong, China; Research Institute for Bioresources and Biotechnology, Ishikawa Prefectural University, Nonoichi, Japan; Department of Microbiology and Immunology, Peter Doherty Institute for Infection and Immunity, University of Melbourne, Melbourne, VIC, Australia; Centre for Immunology & Infection C2i, Hong Kong, China

**Keywords:** COVID-19, vaccine, CoronaVac, intradermal, immunity, adolescent

## Abstract

Strategies to improve the immunogenicity of COVID-19 vaccines are necessary to optimise their protection against disease. Fractional dosing by intradermal administration (ID) has been shown to be equally immunogenic as intramuscular (IM) for several vaccines, but the immunogenicity of ID inactivated whole-virus SARS-CoV-2 at the full dose is unknown. This study (NCT04800133) investigated the superiority of antibody and T cell responses of full-dose CoronaVac by ID over IM in adolescents. Participants aged 11-17 years received 2 doses IM or ID, followed by the 3^rd^ dose 13-42 days later. Humoral and cellular immunogenicity outcomes were measured post-dose 2 (IM-CC versus ID-CC) and post-dose 3 (IM-CCC versus ID-CCC). Doses 2 and 3 were administered to 173 and 104 adolescents, respectively. S IgG, S-RBD IgG, S IgG Fc*γ*RIIIa-binding, SNM-specific IL-2^+^CD4^+^, SNM-specific IL-2^+^CD8^+^, S-specific IL-2^+^CD8^+^, N-specific IL-2^+^CD4^+^, N-specific IL-2^+^CD8^+^ and M-specific IL-2^+^CD4^+^ responses fulfilled the superior and non-inferior criteria for ID-CC compared to IM-CC, whereas IgG avidity was inferior. For ID-CCC, S-RBD IgG, surrogate virus neutralisation test (sVNT), 90% plaque reduction neutralisation titre (PRNT90), PRNT50, S IgG avidity, S IgG Fc*γ*RIIIa-binding, M-specific IL-2^+^CD4^+^, interferon-*γ*^+^CD8^+^ and IL-2^+^CD8^+^ responses were superior and non-inferior to IM-CCC. The estimated vaccine efficacies were 49%, 52%, 66% and 79% for IM-CC, ID-CC, IM-CCC and ID-CCC, respectively. More in the ID groups reported local, mild adverse reactions. This is the first study to demonstrate superior antibody and M-specific T cell responses by ID inactivated SARS-CoV-2 vaccination and serves as the basis for future research to improve immunogenicity of inactivated vaccines.

## INTRODUCTION

The coronavirus disease 2019 (COVID-19) pandemic caused by the severe acute respiratory syndrome coronavirus 2 (SARS-CoV-2) remains of major global public health concern. Although hospitalisations were rarer for adolescents, severe disease still occurred.^1^ During an outbreak by Omicron variants in Hong Kong (HK) in 2022, paediatric hospitalisations increased, with acute neurological and respiratory complications, multisystem inflammatory syndrome in children (MIS-C), long COVID and mental health issues being reported in children and young people, outcomes which can be ameliorated by vaccination.^1–5^

Initial landmark trials demonstrated that the nucleoside-modified mRNA vaccine, BNT162b2, and inactivated whole-virus vaccine, CoronaVac, had ∼90-95% and ∼50-85% efficacies against symptomatic COVID-19 in persons aged ≥16 and ≥18 years old, respectively.^6–8^ The efficacy for BNT162b2 in another phase 3 study for 12 to 15-year-old adolescents was 100%.^9^ These vaccines, with efficacies >50%, have been approved for emergency use since 2021. In a phase 2 trial on CoronaVac for adolescents, 2 doses induced 100% seroconversion in those 12-17 years old.^10^ However, the real-life effectiveness for CoronaVac in the prevention of hospitalisation for adolescents was lower, at ∼90% in Chile and HK, and it is further reduced against infection.^5, 11, 12^

As inactivated vaccines, namely CoronaVac, has been amongst the most widely used COVID-19 vaccines for individuals *≥*3 years old globally, our group performed a humoral and cellular immunobridging study during the early phase of vaccine availability and found non-inferior immunogenicity for CoronaVac in adolescents compared to adults.^13, 14^ However, CoronaVac induced lower antibody responses than BNT162b2 in adolescents and performed similarly in adolescents as adults.^14^ These findings were consistent with the observation that CoronaVac’s efficacies against infection appeared lower than BNT162b2 in separate pivotal clinical trials.^6–8^

Moreover, emerging VOCs, such as various Omicron subvariants, developed mutations at numerous sites that allow neutralising antibody escape for previously infected or vaccinated individuals, further raising concerns for reduced efficacies.^15^ In our recent study, the immunogenicity of CoronaVac against Omicron was markedly lower than the wild type (WT) strain in adolescents.^16^ As a result of immune evasion by these VOCs and waning antibodies, inclusion of the third dose as part of the primary series of CoronaVac had been recommended in Hong Kong and Singapore for most age groups. It is apparent that all feasible strategies on optimizing immunological responses to CoronaVac need to be urgently explored.

Intradermal vaccination (ID) has been shown to be safe and can enhance immunogenicity compared to the intramuscular route (IM).^17^ Introduction of viral antigens and adjuvant into the skin activates resident innate cells, including dermal CD14^+^ dendritic cells, Langerhans cells and mast cells that secrete cytokines, cross present to CD8^+^ T cells and prime CD4^+^ T cells to induce switching of naïve B cells into IgG- and IgA-producing isotypes.^18–21^ Fractional vaccine dosing has been studied extensively for ID to mitigate vaccine inequity during supply shortages or unaffordable costs, especially for the inactivated poliovirus (IPV), hepatitis B (HBV) and human papillomavirus (HPV) vaccines, which showed similar humoral immunogenicity as full doses of IM.^22–24^

Inactivated influenza vaccines, produced by similar technology as CoronaVac, also induced similar IgG titres with fractional dosing by the ID as the full IM dose.^17^ In 2 separate studies, our group showed children 6 months to 17 years old who received inactivated influenza vaccination intradermally at one-fifth of the IM dose developed similar antibody responses to the conventional IM dose.^25, 26^ For full ID dosing, two trials found superior geometric mean (GM) haemagglutination inhibition antibody titres and seroprotection rates compared to IM of the inactivated influenza vaccines in older adults.^27, 28^

Several investigators recently compared fractional ID ChAdOx1/AZD-1222 with BNT162b2 boosters after 2 IM injections of CoronaVac, which induced similar antibody and T cell responses as IM boosters of the same respective vaccine.^29, 30^ However, ID usage of full doses of COVID-19 vaccines has not been studied thus far. Based on the collective available scientific data, we postulated that ID with the full dose of CoronaVac can induce greater immunogenicity against SARS-CoV-2 than the currently recommended IM. This study aimed to compare the reactogenicity of 2 and 3 full doses of CoronaVac between ID and IM and show superior immunogenicity with ID for adolescents 11-17 years old. The current study presents a pre-specified interim analysis of the immunogenicity against the WT and Omicron SARS-CoV-2, reactogenicity and safety results at 1 month after 2 and 3 doses of CoronaVac.

## RESULTS

### Study participants

A total of 185 adolescents aged 11-17 years received at least 1 dose of IM or ID CoronaVac at the screening visit (V1) from 27 April 2021 to 06 August 2022 (see Methods; Supp. Fig. 1). There were 185 and 178 who returned for subsequent follow-up visits 2 (V2) and 3 (V3), respectively, and those who attended V3 were included in the reactogenicity and safety analyses (healthy safety population; see Methods, and Protocol and Statistical Analysis Plan in Supplementary Information; Supp. Fig. 1). The evaluable analysis population included those uninfected based on the clinical history obtained and negative ORF8 IgG (a serological marker of past natural SARS-CoV-2 infection) at every visit, who had negative baseline S-RBD IgG, no major protocol deviations and a valid immunogenicity result (see Methods; Supp. Fig. 1). Of the 173 who completed the 2-dose series (IM-CC and ID-CC), 104 received 3 vaccine doses (IM-CCC and ID-CCC) and returned for the follow-up visit (V4), all within the evaluable intervals (Supp. Fig. 1). A total of 119 IM-CC and 54 ID-CC were included in the evaluable analysis population, with 60 IM and 44 ID recipients for 3 doses. We confirmed these findings by performing an analysis with the expanded analysis population that consisted of 119 IM-CC, 59 ID-CC, 82 IM-CCC and 43 ID-CCC who had wider time intervals of vaccination and blood sampling (see Methods; Supp. Fig. 1). There was an even distribution of demographic characteristics between the IM and ID groups (Supp. Table 1).

### Humoral immunogenicity analyses between IM and ID administrations

The primary humoral immunogenicity outcomes in this study were SARS-CoV-2 S IgG, S-RBD IgG by enzyme-linked immunosorbent assay (ELISA), surrogate virus neutralisation test (sVNT), plaque reduction neutralisation titres (PRNT), spike (S) IgG avidity and S IgG Fc*γ* receptor IIIa (Fc*γ*RIIIa)-binding on ELISA performed for healthy, uninfected adolescents 13-42 days after dose 2 or 3 of CoronaVac by IM or ID (see Methods). Evaluable IM achieved 96.6% S-RBD IgG seropositivity, with geometric mean (GM) optical density-450 (OD450) and sVNT inhibition of 1.20 and 71.2% post-dose 2, respectively (Table 1). 100.0% of evaluable ID-CC had positive (S receptor-binding domain) S-RBD IgG, with GM OD450 value of 2.16 and sVNT inhibition of 78.4%. GM for PRNT90 was 9.83 and 10.7 after IM-CC and ID-CC, respectively. GM for PRNT50 against WT was 26.8 and 30.2 after IM-CC and ID-CC, respectively. After IM-CC and ID-CC, GM avidity was 20.5% and 6.95%, and GM OD450 of S IgG Fc*γ*RIIIa-binding were 0.749 and 1.10, respectively.

**Table 1.** Humoral immunogenicity outcomes against wild type SARS-CoV-2 post-dose 2 and post-dose 3 of CoronaVac in the evaluable analysis population.

|  |  |  |  |  |
| --- | --- | --- | --- | --- |
|  | **Intramuscular** |  | **Intradermal** |  |
|  | **2 doses** | **3 doses** | **2 doses** | **3 doses** |
| **S IgG on ELISA** |  |  |  |  |
| N | 116 | 56 | 47 | 36 |
| GM OD450 value (95% CI) | 0.536 (0.493-0.582) | 0.930 (0.830-1.04) | 0.634 (0.562-0.716) | 1.05 (0.983-1.12) |
| % positive (≥LOD at 0.3) | 94.0% | 98.2% | 95.7%, $P>0.9999$ | 100%, $P>0.9999$ |
| **S-RBD IgG on ELISA** |  |  |  |  |
| N | 119 | 60 | 54 | 42 |
| GM OD450 value (95% CI) | 1.20 (1.10-1.31) | 1.77 (1.68-1.87) | 2.16 (2.04-2.29) | 2.45 (2.33-2.57) |
| % positive (≥LOD at 0.5) | 96.6% | 100% | 100%, $P=0.311$ | 100%, $P>0.9999$ |
| **S-RBD ACE2-blocking antibody on sVNT** |  |  |  |  |
| N | 119 | 60 | 54 | 42 |
| GM % inhibition (95% CI) | 71.2 (66.7-76.0) | 84.9 (81.3-88.6) | 78.4 (74.6-82.5) | 91.3 (88.3-94.4) |
| % positive (≥LOQ at 30%) | 96.6% | 100% | 100%, $P=0.311$ | 100%, $P>0.9999$ |
| **Neutralising antibody on PRNT** |  |  |  |  |
| N | 119 | 60 | 54 | 42 |
| GM PRNT90 (95% CI) | 9.83 (8.67-11.1) | 17.8 (13.7-23.3) | 10.7 (8.09-14.1) | 38.7 (27.6-54.2) |
| % positive (≥LOD at 10) | 65.6% | 78.3% | 61.1%, $P=0.610$ | 97.6%, $P=0.0068$ |
| GM PRNT50 (95% CI) | 26.8 (23.0-31.1) | 55.3 (43.2-70.7) | 30.2 (23.5-38.8) | 110 (82.5-145) |
| % positive (≥LOD at 10) | 96.6% | 100% | 98.2%, $P>0.9999$ | 100%, $P>0.9999$ |
| **S IgG avidity on ELISA** |  |  |  |  |
| N | 109 | 55 | 45 | 36 |
| GM avidity index (95% CI) | 20.5 (19.1-22.1) | 38.5 (34.6-42.8) | 6.95 (5.03-9.60) | 53.3 (48.3-58.8) |
| **S IgG FcγRIIIa-binding on ELISA** |  |  |  |  |
| N | 116 | 56 | 47 | 36 |
| GM OD450 value (95% CI) | 0.749 (0.649-0.864) | 1.41 (1.21-1.63) | 1.10 (0.955-1.27) | 1.79 (1.71-1.88) |
| % positive (≥LOD at 0.28) | 87.1% | 98.2% | 95.7%, $P=0.156$ | 100%, $P>0.9999$ |
| GM, geometric mean; OD, optical density; LOD, limit of detection; LOQ, limit of quantification; CI, confidence interval; S, spike protein; ELISA, enzyme-linked immunosorbent assay; RBD, receptor-binding domain; ACE-2, angiotensin-converting enzyme-2; sVNT, surrogate virus neutralisation test; PRNT, plaque reduction neutralisation titre; PRNT90, 90% plaque reduction neutralisation titre; PRNT50, 50% plaque reduction neutralisation titre; FcγRIIIa, Fc gamma receptor III-a. $P$-values compare the proportion of positive responses between intramuscular and intradermal administration by Fisher's exact test. | | | | |

Compared to IM-CC, humoral responses when measured by S IgG (GM ratio (GMR) 1.18, 95%CI 1.02-1.38, *P*=0.028), S-RBD IgG (GMR 1.80, 95%CI 1.58-2.05, *P*<0.0001) and S IgG Fc*γ*RIIIa-binding (GMR 1.47, 95%CI 1.16-1.87, *P*=0.002) (Fig. 1A) satisfied the superior and non-inferior criteria for evaluable ID-CC. ID-CC mounted non-inferior humoral responses by sVNT (GMR 1.10, 95%CI 0.99-1.22, *P*=0.065), PRNT90 (GMR 1.09, 95%CI 0.84-1.41, *P*=0.536), PRNT50 (GMR 1.13, 95%CI 0.85-1.49, *P*=0.397) and inferior S IgG avidity (GMR 0.34, 95%CI 0.27-0.43, *P*<0.0001).

**Fig. 1.**
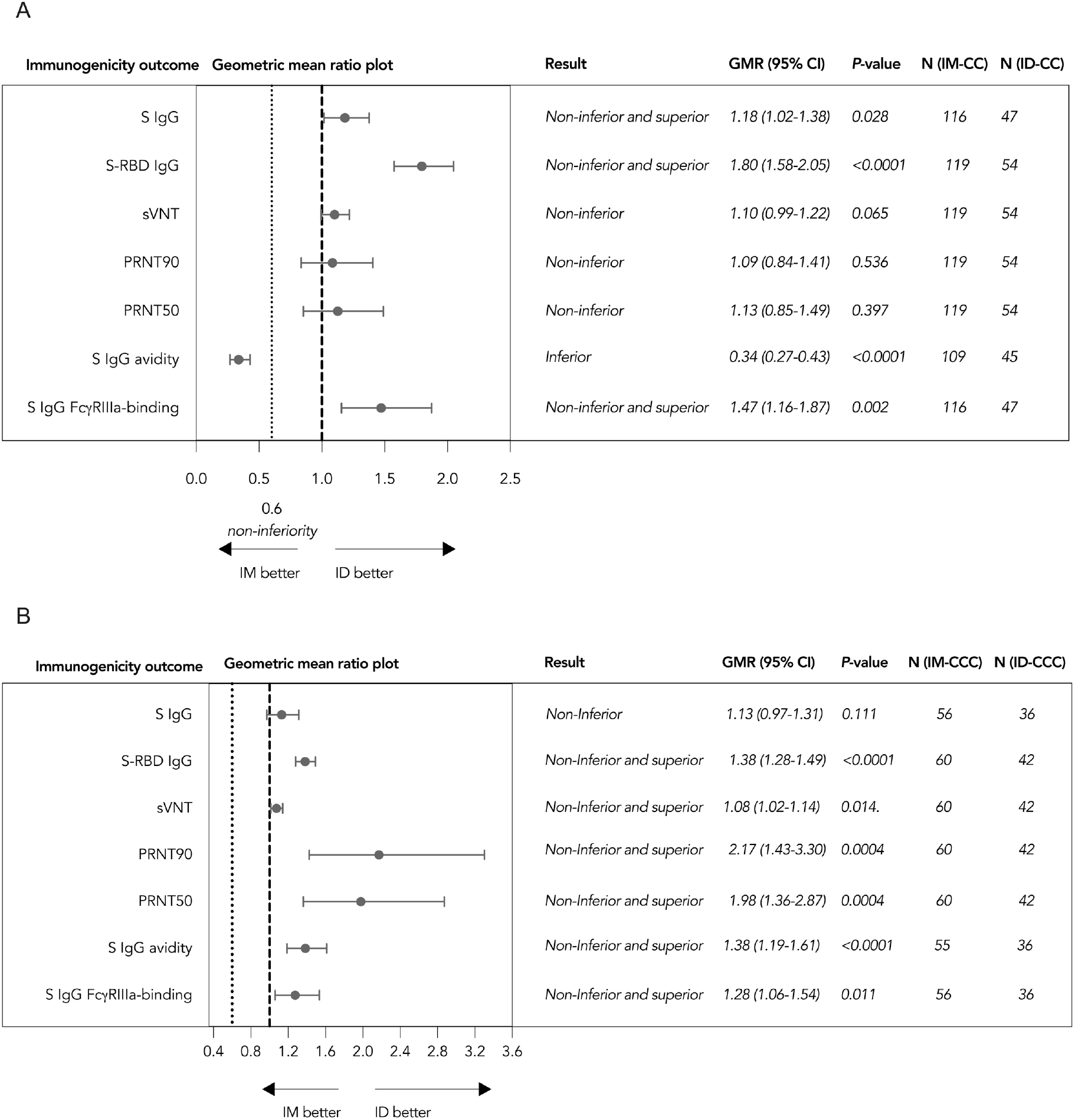
Superiority and non-inferiority hypotheses testing of humoral immunogenicity against wild type SARS-CoV-2 post-dose 2 and post-dose 3 of vaccination in the evaluable analysis population. Adolescents receiving (A) 2 doses of CoronaVac administered intramuscularly (IM-CC) or intradermally (ID-CC) and (B) 3 doses of CoronaVac administered intramuscularly (IM-CCC) or intradermally (ID-CCC) were tested for humoral immunogenicity outcomes. Dots and error bars show GMR estimates and two-sided 95% CI, respectively. GMR, geometric mean ratio; CI, confidence interval; ID, intradermal injection; IM, intramuscular injection; S, spike protein; RBD, receptor-binding domain; sVNT, surrogate virus neutralisation test; PRNT, plaque reduction neutralisation titres; FcγRIIIa, Fcγ receptor IIIa.

Since IM-CCC had been recommended as the primary vaccination series in HK and Singapore, this regimen was also compared with ID-CCC. Evaluable IM-CCC achieved 100.0% S-RBD IgG seropositivity, with GM OD450 and sVNT inhibition of 1.77 and 84.9% post-dose 3, respectively (Table 1). 100.0% of evaluable ID-CCC had positive S-RBD IgG, with GM OD450 value of 2.45 and sVNT inhibition of 91.3%. Neutralisation titres demonstrated GM of 17.8 and 38.7 for PRNT90 after IM-CCC and ID-CCC, respectively. GM for PRNT50 was 55.3 and 110 after IM-CCC and ID-CCC, respectively. GM avidity was 38.5% and 53.3%, and the GM OD450 results of S IgG Fc*γ*RIIIa-binding were 1.41 and 1.79, after IM-CCC and ID-CCC, respectively.

ID-CCC satisfied the superior and non-inferior criteria by S-RBD IgG (GMR 1.38, 95%CI 1.28-1.49, *P*<0.0001), sVNT (GMR 1.08, 95%CI 1.02-1.14, *P*=0.014), PRNT90 (GMR 2.17, 95%CI 1.43-3.30, *P*=0.0004), PRNT50 (GMR 1.98, 95%CI 1.36-2.87, *P*=0.0004), S IgG avidity (GMR 1.38, 95%CI 1.19-1.61, *P*<0.0001) and S IgG Fc*γ*RIIIa-binding (GMR 1.28, 95%CI 1.06-1.54, *P*=0.011), but not by S IgG (GMR 1.13, 95%CI 0.97-1.31, *P*=0.111), which satisfied the non-inferiority criterion only (Fig. 1B). Superiority and non-inferiority testing in the expanded analysis populations for post-doses 2 and 3 were analogous to these results (Supp. Table 2; Supp. Fig. 2).

### Cellular immunogenicity analyses between IM and ID administrations

The primary cellular immunogenicity outcomes for this study were interferon-*γ* (IFN-*γ*)^+^ and interleukin-2 (IL-2)^+^ CD4^+^ and CD8^+^ T cells responses to S, N (nucleocapsid protein) and M (membrane protein) after IM-CC and ID-CC, which were analysed using intracellular cytokine staining by flow cytometry (see Methods). For the 60 CD-IM and 48 CC-ID evaluable adolescents, more than half of participants had detectable responses for S-specific IFN-*γ*^+^CD4^+^ or IL-2^+^CD4^+^ T cells after 2 ID or IM doses (Table 2). There were 45.8-52.1% of those with IFN-*γ*^+^CD8^+^ and IL-2^+^CD8^+^ T cell responses to S after 2 doses of either administration routes. The remainder of the T cell responses to the peptide pools S, N, M and SNM (sum of individual S, N, and M peptide pools) after IM-CC, ID-CC, IM-CCC and ID-CCC also are shown in Table 2.

**Table 2.** Cellular immunogenicity outcomes against wild type SARS-CoV-2 S, N and M peptide pools post-dose 2 and post-dose 3 of CoronaVac in the evaluable analysis population.

|  | Intramuscular |  | Intradermal |  |
| --- | --- | --- | --- | --- |
|  | 2 doses | 3 doses | 2 doses | 3 doses |
| **Total SNM-specific T cell responses on flow cytometry** |  |  |  |  |
| N | 60 | 58 | 48 | 40 |
| **GM** % IFN-γ<sup>+</sup>CD4<sup>+</sup> T cells (95% CI) | 0.0581%<br>(0.0405-0.0833%) | 0.0662%<br>(0.0414-0.106%) | 0.107%<br>(0.0628-0.183%) | 0.102%<br>(0.0530-0.197%) |
| % positive (≥cut-off at 0.0075%) | 83.3% | 74.1% | 81.3%, $P=0.804$ | 77.5%, $P=0.813$ |
| **GM** % IL-2<sup>+</sup>CD4<sup>+</sup> T cells (95% CI) | 0.0395%<br>(0.0301-0.0517%) | 0.0732%<br>(0.0491-0.109%) | 0.112%<br>(0.0680-0.183%) | 0.143%<br>(0.0772-0.264%) |
| % positive (≥cut-off at 0.0075%) | 83.3% | 79.3% | 79.2%, $P=0.624$ | 80.0%, $P>0.9999$ |
| **GM** % IFN-γ<sup>+</sup>CD8<sup>+</sup> T cells (95% CI) | 0.0501%<br>(0.0328-0.0766%) | 0.0708%<br>(0.0400-0.125%) | 0.0589%<br>(0.0333-0.104%) | 0.0623%<br>(0.0315-0.123%) |
| % positive (≥cut-off at 0.0075%) | 65.0% | 62.1% | 62.5%, $P=0.842$ | 62.5%, $P>0.9999$ |
| **GM** % IL-2<sup>+</sup>CD8<sup>+</sup> T cells (95% CI) | 0.0173%<br>(0.0138-0.0216%) | 0.0414%<br>(0.0273-0.0628%) | 0.0498%<br>(0.0309-0.0803%) | 0.0664%<br>(0.0344-0.128%) |
| % positive (≥cut-off at 0.0075%) | 58.3% | 65.5% | 68.8%, $P=0.318$ | 65.0%, $P>0.9999$ |
| **S-specific T cell responses on flow cytometry** |  |  |  |  |
| N | 60 | 59 | 48 | 41 |
| **GM** % IFN-γ<sup>+</sup>CD4<sup>+</sup> T cells (95% CI) | 0.0233%<br>(0.0149-0.0362%) | 0.0162%<br>(0.00979-0.0268%) | 0.0220%<br>(0.0120-0.0405%) | 0.0251%<br>(0.0130-0.0484%) |
| % positive (≥cut-off at 0.005%) | 70.0% | 57.6% | 58.3%, $P=0.229$ | 68.3%, $P=0.303$ |
| **GM** % IL-2<sup>+</sup>CD4<sup>+</sup> T cells (95% CI) | 0.0147%<br>(0.0106-0.0204%) | 0.0168%<br>(0.0105-0.0267%) | 0.0205%<br>(0.0119-0.0353%) | 0.0313%<br>(0.0166-0.0589%) |
| % positive (≥cut-off at 0.005%) | 73.3% | 61.0% | 60.4%, $P=0.214$ | 68.3%, $P=0.528$ |
| **GM** % IFN-γ<sup>+</sup>CD8<sup>+</sup> T cells (95% CI) | 0.0142%<br>(0.00861-0.0235%) | 0.0171%<br>(0.00951-0.0309%) | 0.0138%<br>(0.00772-0.0247%) | 0.00986%<br>(0.00524-0.0186%) |
| % positive (≥cut-off at 0.005%) | 48.3% | 49.2% | 45.8%, $P=0.848$ | 39.0%, $P=0.414$ |
| **GM** % IL-2<sup>+</sup>CD8<sup>+</sup> T cells (95% CI) | 0.00625%<br>(0.00476-0.00822%) | 0.00948%<br>(0.00606-0.0148%) | 0.0121%<br>(0.00727-0.0202%) | 0.00952%<br>(0.00514-0.0176%) |
| % positive (≥cut-off at 0.005%) | 48.3% | 47.5% | 52.1%, $P=0.847$ | 41.5%, $P=0.683$ |
| **N-specific T cell responses on flow cytometry** |  |  |  |  |
| N | 60 | 58 | 48 | 40 |
| **GM** % IFN-γ<sup>+</sup>CD4<sup>+</sup> T cells (95% CI) | 0.0113%<br>(0.00752-0.0170%) | 0.0140%<br>(0.00826-0.0239%) | 0.0222%<br>(0.0114-0.0433%) | 0.0233%<br>(0.0116-0.0470%) |
| % positive (≥cut-off at 0.005%) | 55.0% | 53.5% | 54.2%, $P>0.9999$ | 60.0%, $P=0.542$ |
| **GM** % IL-2<sup>+</sup>CD4<sup>+</sup> T cells (95% CI) | 0.0126%<br>(0.00899-0.0176%) | 0.0199%<br>(0.0122-0.0326%) | 0.0281%<br>(0.0149-0.0531%) | 0.0298%<br>(0.0150-0.0592%) |
| % positive (≥cut-off at 0.005%) | 66.7% | 60.3% | 60.4%, $P=0.549$ | 65.0%, $P=0.677$ |
| **GM** % IFN-γ<sup>+</sup>CD8<sup>+</sup> T cells (95% CI) | 0.00767%<br>(0.00485-0.0121%) | 0.0148%<br>(0.00794-0.0275%) | 0.0163%<br>(0.00842-0.0314%) | 0.0126%<br>(0.00612-0.0259%) |
| % positive (≥cut-off at 0.005%) | 31.7% | 41.4% | 43.8%, $P=0.232$ | 40.0%, $P>0.9999$ |
| **GM** % IL-2<sup>+</sup>CD8<sup>+</sup> T cells (95% CI) | 0.00419%<br>(0.00330-0.00532%) | 0.0101%<br>(0.00628-0.0164%) | 0.0135%<br>(0.00754-0.0241%) | 0.0141%<br>(0.00737-0.0271%) |
| % positive (≥cut-off at 0.005%) | 28.3% | 41.4% | 47.9%, $P=0.0457$ | 50.0%, $P=0.417$ |
| **M-specific T cell responses on flow cytometry** |  |  |  |  |
| N | 60 | 59 | 48 | 40 |
| **GM** % IFN-γ<sup>+</sup>CD4<sup>+</sup> T cells (95% CI) | 0.00706%<br>(0.00483-0.0103%) | 0.00570%<br>(0.00369-0.00881%) | 0.00827%<br>(0.00479-0.0143%) | 0.00967%<br>(0.00462-0.0202%) |
| % positive (≥cut-off at 0.005%) | 36.7% | 23.7% | 35.4%, $P>0.9999$ | 32.5%, $P=0.365$ |
| **GM** % IL-2<sup>+</sup>CD4<sup>+</sup> T cells (95% CI) | 0.00571%<br>(0.00449-0.00728%) | 0.00599%<br>(0.00399-0.00899%) | 0.0107%<br>(0.00617-0.0187%) | 0.0137%<br>(0.00647-0.0288%) |
| % positive (≥cut-off at 0.005%) | 46.7% | 25.4% | 41.7%, $P=0.698$ | 45.0%, $P=0.0522$ |
| **GM** % IFN-γ<sup>+</sup>CD8<sup>+</sup> T cells (95% CI) | 0.00588%<br>(0.00390-0.00886%) | 0.00506%<br>(0.00314-0.00815%) | 0.00661%<br>(0.00399-0.0110%) | 0.0121%<br>(0.00604-0.0242%) |
| % positive (≥cut-off at 0.005%) | 25.0% | 13.6% | 29.2%, $P=0.667$ | 42.5%, $P=0.0019$ |
| **GM** % IL-2<sup>+</sup>CD8<sup>+</sup> T cells (95% CI) | 0.00383%<br>(0.00309-0.00475%) | 0.00454%<br>(0.00321-0.00643%) | 0.00521%<br>(0.00348-0.00778%) | 0.0120%<br>(0.00592-0.0241%) |
| % positive (≥cut-off at 0.005%) | 23.3% | 18.6% | 27.1%, $P=0.662$ | 47.5%, $P=0.0035$ |
| GM, geometric mean; CI, confidence interval; S, spike; N, nucleocapsid protein; M, membrane protein; IFN-γ, interferon-gamma; IL-2, interleukin-2. $P$-values compare the proportion of positive responses between intramuscular and intradermal administration by Fisher's exact test. | | | | |

After 2 doses of ID CoronaVac, SNM-specific IL-2^+^CD4^+^(GMR 2.83, 95%CI 1.67-4.79, *P*=0.0002), SNM-specific IL-2^+^CD8^+^ (GMR 2.88, 95%CI 1.77-4.70, *P*<0.0001), S-specific IL-2^+^CD8^+^ (GMR 1.94, 95%CI 1.13-3.34, *P*=0.017), N-specific IL-2^+^CD4^+^ (GMR 2.24, 95%CI 1.14-4.38, *P*=0.019), N-specific IL-2^+^CD8^+^ (GMR 3.21, 95%CI 1.81-5.71, *P*=0.0001) and M-specific IL-2^+^CD4^+^ (GMR 1.88, 95%CI 1.08-3.28, *P*=0.027) T cell responses were superior and non-inferior to IM (Fig. 2A). SNM-specific IFN-*γ*^+^CD8^+^, S-specific IFN-*γ*^+^CD4^+^ and S-specific IFN-*γ*^+^CD8^+^ T cell responses were inconclusive, while other T cell responses were non-inferior. Additionally, evaluable ID-CCC satisfied superior and non-inferior criteria for M-specific IL-2^+^CD4^+^(GMR 2.28, 95%CI 1.05-4.96, *P*=0.038), IFN-*γ*^+^CD8^+^ (GMR 2.39, 95%CI 1.07-5.33, *P*=0.034) and IL-2^+^CD8^+^ (GMR 2.63, 95%CI 1.30-5.32, *P*=0.008) T cell responses compared to IM-CCC (Fig. 2B). SNM-specific IFN-*γ*^+^CD8^+^, S-specific IFN-*γ*^+^CD8^+^, S-specific IL-2^+^CD8^+^ and N-specific IFN-*γ*^+^CD8^+^ T cell responses were inconclusive, while the other T cell responses were non-inferior. These results were consistent with superiority and non-inferiority testing in the expanded analysis populations (Supp. Table 3; Supp. Fig. 3). Overall, none of the cellular immunogenicity outcomes tested for groups that received ID was inferior compared to IM.

**Fig. 2.**
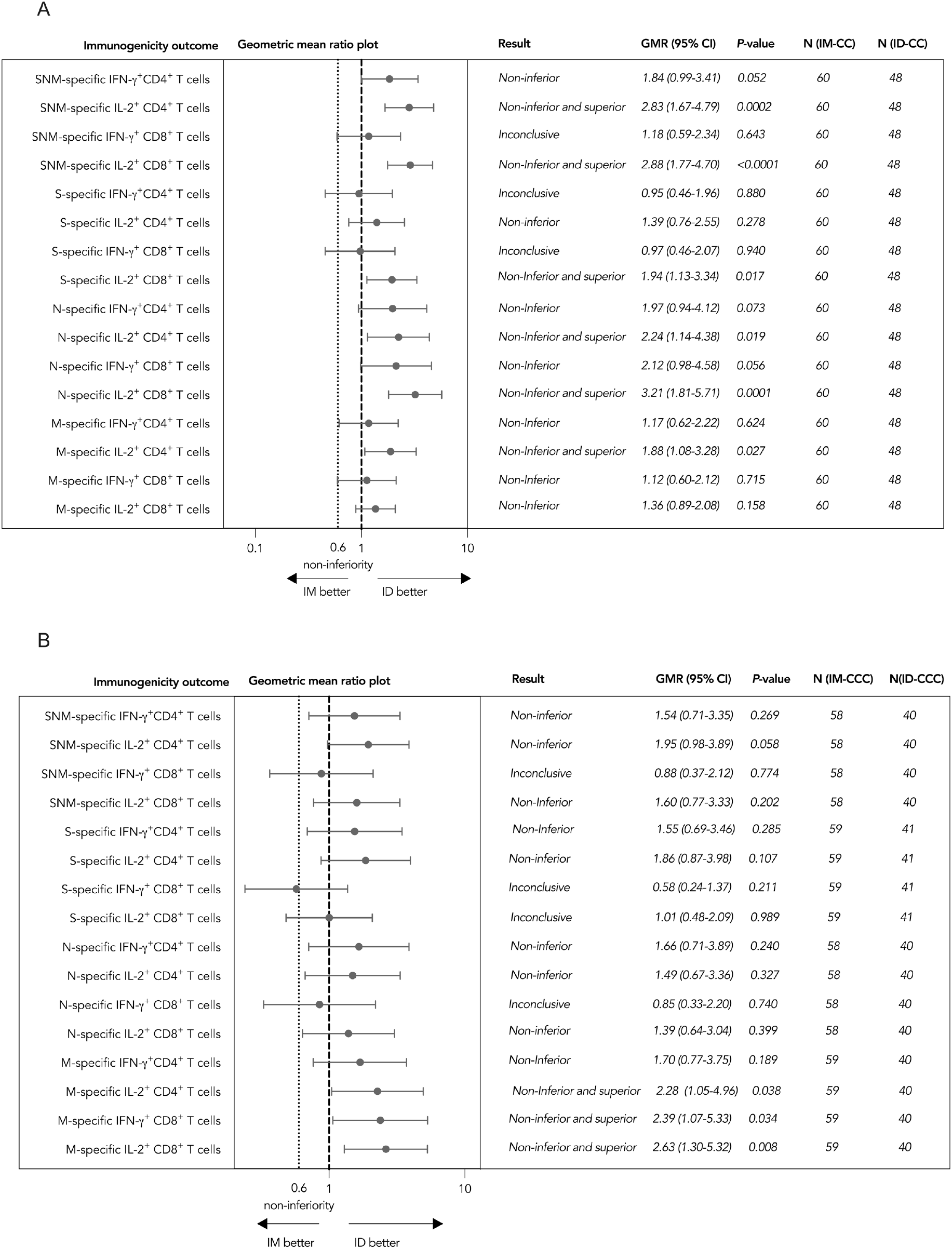
Superiority and non-inferiority hypotheses testing of cellular immunogenicity against wild type SARS-CoV-2 post-dose 2 and post-dose 3 of vaccination in the evaluable analysis population. Adolescents receiving (A) 2 doses of CoronaVac administered intramuscularly (IM-CC) or intradermally (ID-CC) and (B) 3 doses of CoronaVac administered intramuscularly (IM-CCC) or intradermally (ID-CCC) were tested for T cell responses by flow-cytometry-based intracellular cytokine staining assays specific to S, N and M post-dose 2 or post-dose 3. The results of SNM-specific T cell responses were calculated from the sum of responses of the individual S, N and M peptide pools. Dots and error bars show GMR estimates and two-sided 95% CI respectively. GMR, geometric mean ratio; CI, confidence interval; ID, intradermal injection; IM, intramuscular injection; S, spike protein; N, nucleocapsid protein; M, membrane protein; IFN-γ, interferon-γ; IL-2, interleukin-2.

### Longitudinal immunogenicity changes between doses over time within the same group of IM or ID

Antibody and T cell responses were compared between post-doses 2 and 3 in the evaluable analysis populations. Overall, dose 3 induced higher humoral responses for S IgG, S-RBD IgG, sVNT, PRNT90, PRNT50, S IgG avidity and S IgG Fc*γ*RIIIa-binding than dose 2 by the respective IM or ID routes (Supp. Fig. 4). Additionally, there were higher SNM-specific IL-2^+^CD4^+^, SNM-specific IL-2^+^CD8^+^ and N-specific IL-2^+^CD8^+^ T cell responses after IM-CCC compared to IM-CC (Supp. Fig. 5). ID-CCC induced a higher M-specific IL-2^+^CD8^+^ T cell response than ID-CC.

### Estimation of vaccine efficacies from different doses and routes of administration of CoronaVac based on neutralisation titres

Levels of neutralising antibodies have been regarded as a correlate of protection. Hence, we extrapolated our PRNT50 results from evaluable adolescents who received 2 or 3 doses of IM or ID CoronaVac with vaccine efficacies against symptomatic COVID-19 by normalization to convalescent sera, as previously described.^14, 16, 31^ The mean neutralisation levels of IM-CC, ID-CC, IM-CCC and ID-CCC were 0.19, 0.22, 0.40 and 0.79, corresponding to 49%, 52%, 66% and 79% vaccine efficacies, respectively (Fig. 3). Using PRNT90 instead of PRNT50 yielded similar findings (data not shown).

**Fig. 3.**
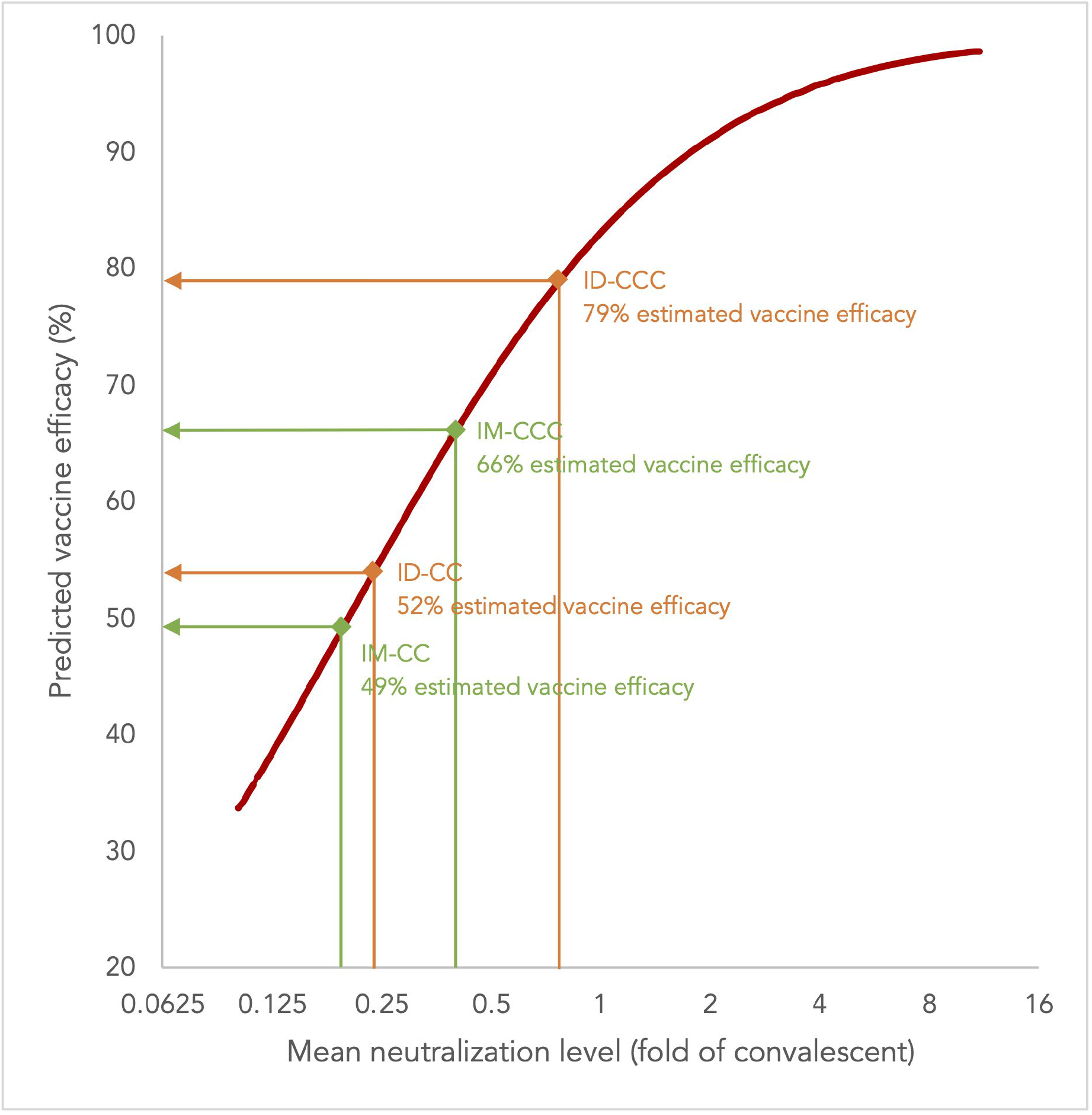
Estimation of vaccine efficacies for 2 doses and 3 doses of CoronaVac by intramuscular or intradermal administration vaccination in the evaluable analysis population based on neutralisation titres against SARS-CoV-2. The vaccine efficacy estimates were based on neutralising antibody titres (PRNT50, or plaque reduction neutralisation test based on the reciprocal of the highest serum dilution that resulted in the cut-off of >50%) post-dose 2 or post-dose 3 of CoronaVac administered intramuscularly (IM-CC or IM-CCC) or intradermally (ID-CC or ID-CCC), as neutralising antibodies have been established as a reliable correlate of protection that can predict vaccine efficacies against symptomatic COVID-19. Dividing the geometric mean titres of PRNT50 who received vaccination by titres from 102 convalescent sera collected on days 28-59 post-onset of illness in patients aged ≥18 years yielded the mean neutralising levels (fold of convalescent). Extrapolation of the point estimates of the vaccine efficacies from the best fit of the logistic model was performed as previously described.^14, 16, 31^ IM-CC (*N*=119) and IM-CCC (*N*=60), post-dose 2 and post-dose 3 of vaccine administered intramuscularly, respectively; ID-CC (*N*=54) and ID-CCC (*N*=42), post-dose 2 and post-dose 3 of vaccine administered intradermally, respectively.

### Omicron-specific humoral and cellular immunogenicity post-dose 2 and post-dose 3 of ID CoronaVac

For ID-CCC, humoral responses tested against the Omicron variant were significantly lower than WT SARS-CoV-2 for S IgG (GM OD450 0.81 vs 1.05, *P*<0.0001) and S IgG Fc*γ*RIIIa-binding (GM OD450 1.33 vs 1.79, *P*<0.001), respectively (Fig. 4A). S IgG avidity and all the cellular immunogenicity outcomes were similar between Omicron and WT (Fig. 4A-D).

**Fig. 4.**
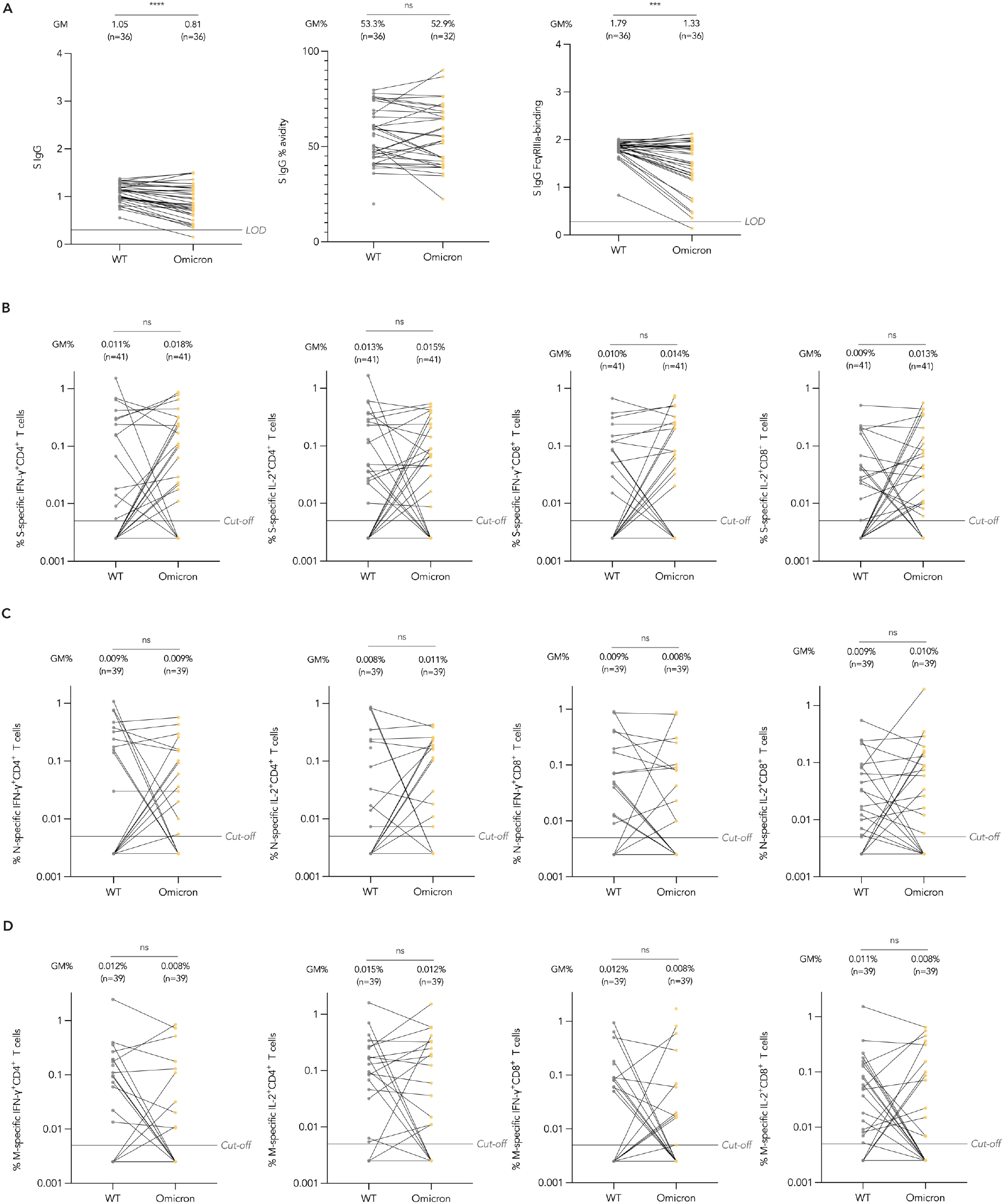
Omicron variant-specific humoral and cellular immunogenicity post-dose 3 of CoronaVac administered intradermally in the evaluable analysis population. (A) Humoral and (B-D) cellular immunogenicity outcomes against WT SARS-CoV-2 and the Omicron variant post-dose 3 of CoronaVac administered intradermally. Data labels and centre lines show GM estimates, and error bars show 95% CI. *P*-values were derived from two-tailed unpaired *t* test after natural logarithmic transformation. GM, geometric mean; CI, confidence interval; WT, wild type; S, spike protein; FcγRIIIa, Fcγ receptor IIIa; IFN-γ interferon-γ, IL-2 interleukin-2; N, nucleocapsid protein; M, membrane protein; IFN-γ, interferon-γ; IL-2, interleukin-2. \*\*\**P*<0.001; \*\*\*\**P*<0.0001; ns, no significant difference.

### Reactogenicity and safety of IM or ID CoronaVac

In the healthy safety population, pain at the injection site was the most reported AR for IM, which was similar to ID (Fig. 5A). Greater proportions of ID reported swelling, erythema and induration (IM-C vs ID-C: 1.7vs 52.5%, *P*<0.0001; IM-CC vs ID-CC: 0.8% vs 64.4%, *P*<0.0001; IM-CCC vs ID-CCC: 1.1% vs 65.1%, *P*<0.0001) and pruritis (IM-C vs ID-C: 0.8% vs 62.7%, *P*<0.0001; IM-CC vs ID-CC: 0.8% vs 50.9%, *P*<0.0001; IM-CCC vs ID-CCC: 1.1% vs 60.5%, *P*<0.0001) at the injection site than IM. Most recipients developed these symptoms within minutes after ID, which progressed locally at the site of inoculation over 1-2 weeks and subsided over several weeks (Fig. 5B-D; Supp. Fig. 6). Systemic ARs were similar between IM and ID (Fig. 6). There were 13 adverse events (AEs) reported within 28 days after vaccination (8 for IM and 5 for ID) and no serious adverse event (SAE) for either administration route (Supp. Table 4).

**Fig. 5.**
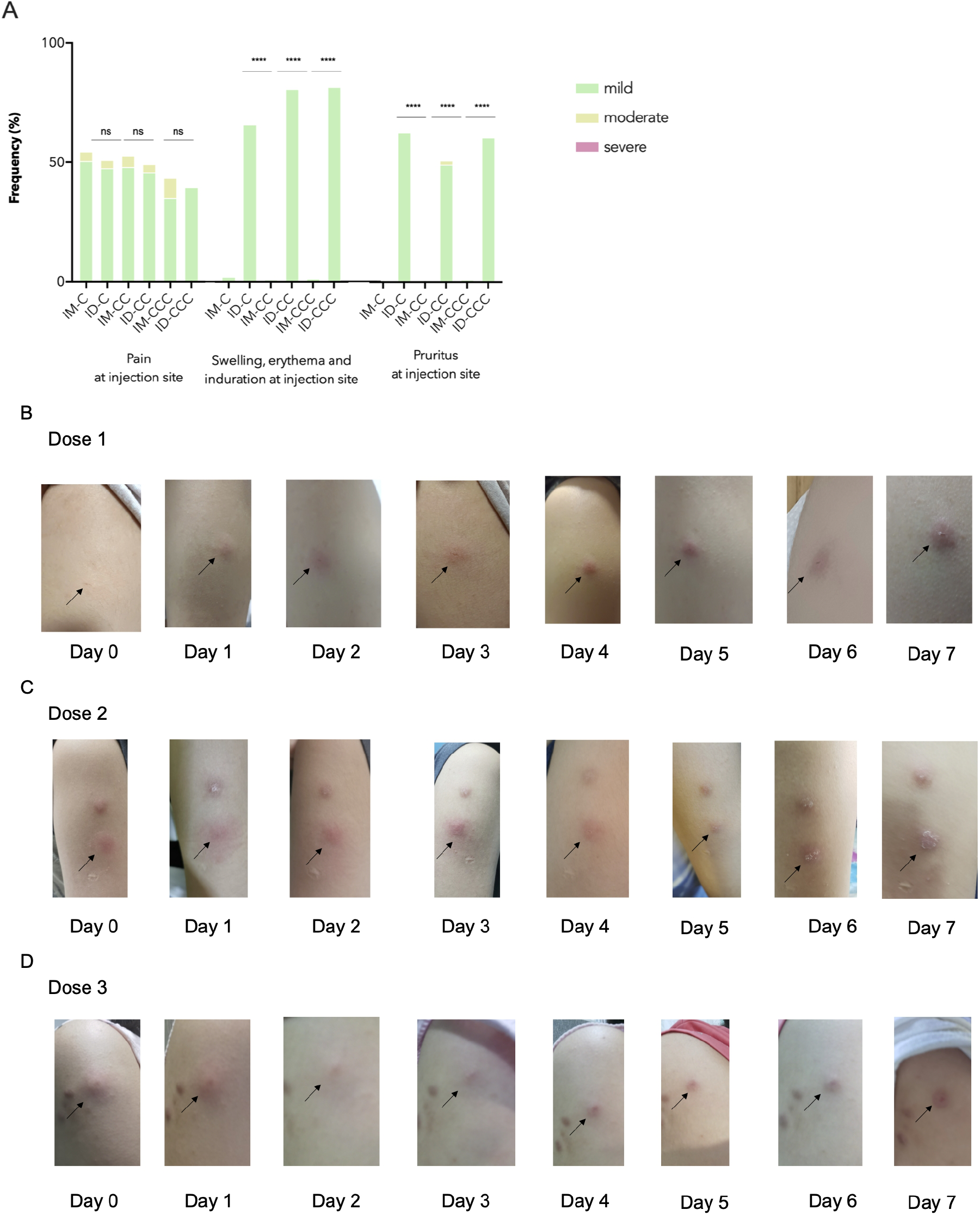
Local adverse reactions in the healthy safety population after doses 1, 2 and 3 of CoronaVac by intramuscular or intradermal administration. (A) Local adverse reactions 7 days after each dose of CoronaVac administered by intramuscular or intradermal injections were solicited from participants in the healthy safety population. Data are shown as percentages of the respective adverse reaction of any severity. (B-D) Photos are representative of the typical injection site reactions manifested for 7 days after doses (B) 1, (C) 2 or (D) 3 of CoronaVac administered intradermally. IM-CC (*N*=119) and IM-CCC (*N*=94), post-dose 2 and post-dose 3 of vaccine administered intramuscularly, respectively; ID-CC (*N*=59) and ID-CCC (*N*=45), post-dose 2 and post-dose 3 of vaccine administered intradermally, respectively. \*\*\*\**P*<0.0001; ns, no significant difference.

**Fig. 6.**
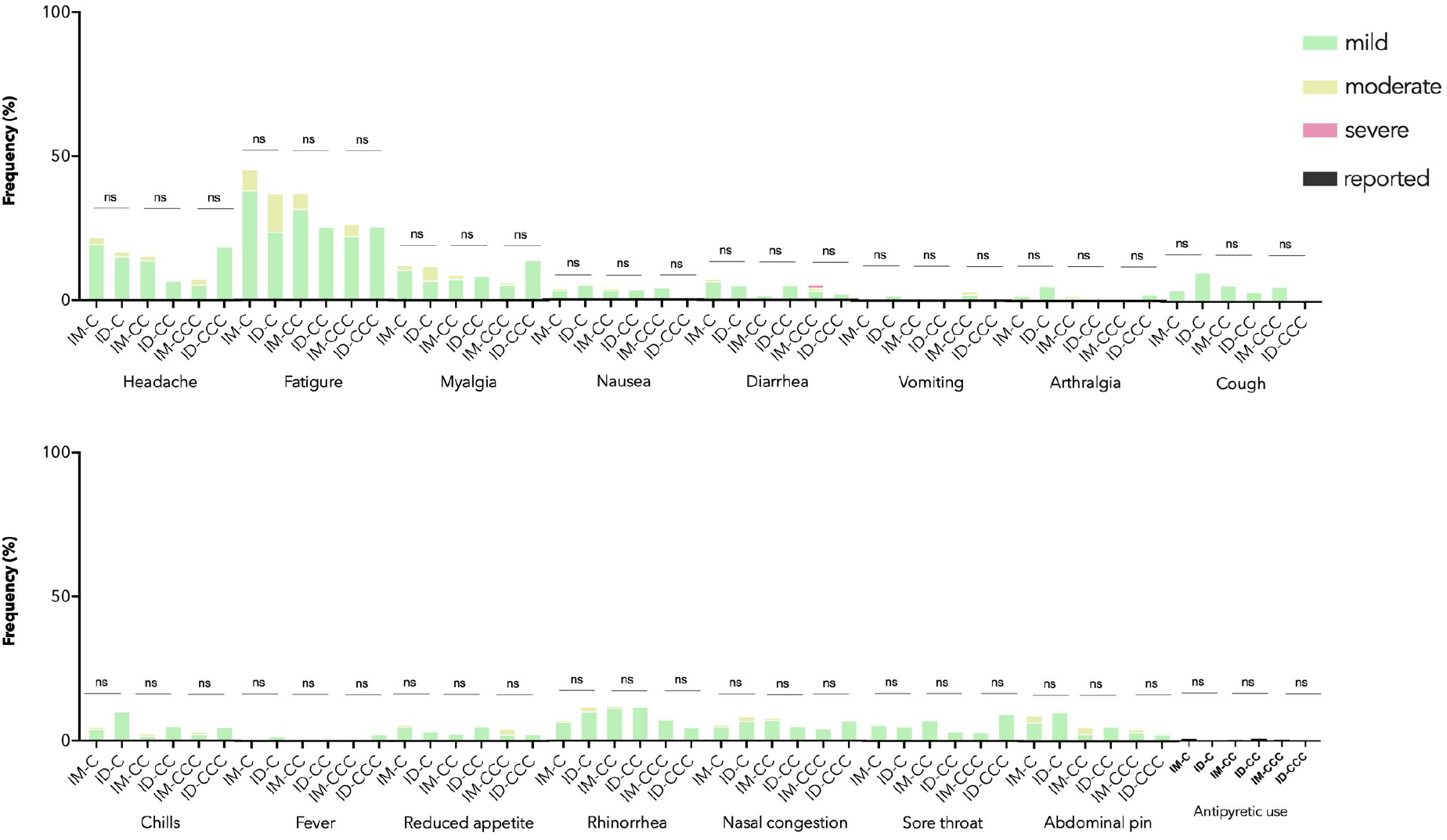
Systemic adverse reactions in the healthy safety population after doses 1, 2 and 3 of CoronaVac by intramuscular or intradermal administration. Adverse reactions 7 days after each dose of CoronaVac administered by intramuscular or intradermal injections were solicited from participants in the healthy safety population. Data are shown as percentages of the respective adverse reaction of any severity. IM-CC (*N*=119) and IM-CCC (*N*=94), post-dose 2 and post-dose 3 of vaccine administered intramuscularly, respectively; ID-CC (*N*=59) and ID-CCC (*N*=45), post-dose 2 and post-dose 3 of vaccine administered intradermally, respectively. ns, no significant difference.

## DISCUSSION

This is the first study to assess the immunogenicity, reactogenicity and safety of intradermal administration of an inactivated COVID-19 vaccine with the full dose, which demonstrated superior antibody responses and T cell responses against the SARS-CoV-2 membrane protein in adolescents who received 3 injections. ID-CCC elicited higher antibody responses across all parameters tested and M-specific IL-2^+^CD8^+^ T cell response than ID-CC. These data estimated the vaccine efficacy of ID-CCC to be 79%, which was ∼30% higher than 2 doses of IM observed in this study (49%), our recent immunobridging publication (50%) and CoronaVac’s initial study in Brazil (51%).^7, 14^ Whether the observed superior immunogenicity and estimated efficacy from ID translate to actual higher clinical efficacy and effectiveness needs to be further studied.

The long-term immune protection from different types and routes of vaccination, particularly against VOCs, is incompletely established. In this study, the levels of anti-spike antibodies and its Fc*γ*RIIIa-binding against Omicron was lower than WT after ID-CCC. However, S IgG avidity and cellular immunogenicity against Omicron were maintained, which can possibly explain the persistently high real-life vaccine effectiveness even when it is known that there is waning of quantitative serum antibody concentrations against VOCs.^5, 12^ Indeed, several human studies have shown that T cell immunity can persist for years after prior exposure and mitigate disease severity when neutralising antibodies are reduced, and the pre-existing antigen-specific T cells are protective against influenza viral infections, severity of symptoms and viral shedding.^32–36^ In addition, vaccine-induced T cell responses against the highly conserved structural SARS-CoV-2 membrane protein confers partial protection from lung pathology in a murine model.^37^

Interestingly, S IgG avidity for ID-CC was inferior compared to IM-CC. However, this was reversed after 3 doses. This was accompanied by a simultaneous shift from non-inferior to superior neutralising antibodies and sVNT, while the differences in S IgG and S-RBD IgG between IM and ID became less pronounced. We speculate despite reduced differences in the quantitative anti-S IgG concentrations after dose 3 between ID and IM, there was greater antibody function and quality from the higher avidity that correspondingly enhanced the neutralisation of SARS-CoV-2.^38^ The high-affinity antibodies that resulted in higher avidity binding are indicative of class switching in germinal centres, affinity maturation and longer lasting functional antibody responses. Furthermore, Fc*γ*RIIIa functions were also increased by ID vaccination. Our recent publication on BNT162b2 by IM demonstrated consistent increases in S IgG avidity across time, while the concentrations of IgGs waned before dose 3 was administered as a booster vaccination to adolescents.^39^ These findings support the notion that effective vaccination induces efficient immunity characterized by antibodies that have high avidity and increased effector functions against SARS-CoV-2 rather than large quantities of low-quality immunoglobulins.^40^ In this regard, S IgG avidity maturation in our cohort appears to be markedly boosted by dose 3 of ID CoronaVac, and the kinetics of this antibody response is significantly different from IM.

This study had some limitations. It was not possible to blind the participants because receipt of IM or ID injections was readily differentiable, and the technique of administration for the 2 routes was also noticeably distinct to the clinical staff. It can be viewed that this unblinded, non-randomised study design is a limitation and has the potential for selection bias. However, since the age, sex and ethnic distributions were similar between groups, the immunogenicity comparisons should be valid. Importantly, this practical approach can be directly interpretable for real-life applicability. For example, although the immunogenicity of ID appears greater than IM, ID injections were associated with more local reactions. This known adverse effect can be a reason for some individuals to select IM rather than ID in the real world. Although ARs were predominantly local symptoms and no SAEs occurred, participants who received ID endured weeks of induration and itching at the site of injection, most of whom described as tolerable. These findings were consistent with the recent study on the novel mpox by ID route that was associated with acceptable local reactions, as both CoronaVac and 3^rd^ generation mpox vaccines are incapable of replication.^41^ Larger pharmacovigilance studies over longer periods of time will be required to delineate whether there are higher risks of rarer adverse effects from full-dose, intradermally administered inactivated COVID-19 vaccination. Additionally, the strict social distancing policies that the HK Government mandated during the COVID-19 period kept infection numbers low so that it was not possible to investigate vaccine efficacy in this cohort during this study period, from April 2021 to August 2022 (see Methods).^1, 5, 12^ On the other hand, this provided the opportunity to determine the intradermal vaccine immunogenicity without the confounding effects of existing immunity acquired from past infections.^1, 5, 12^ Moreover, we were able to estimate actual vaccine efficacies using neutralising antibodies.^14, 16, 31^ The comprehensive, validated assays of antibody levels, avidity and binding and T cell responses were a major strength of this study, yet follow-up research on clinical efficacy and real-life effectiveness against COVID-19 from ID would be needed to further support the current findings. The sample size for the ID group was slightly smaller than the initial target due to the unforeseeable wave of Omicron infections that swept across HK during the study period that doses 2 and 3 were administered. This led to exclusion of more enrolled participants than expected, while a few others under quarantine as close contacts could not provide blood samples within the evaluable window, and these can lead to selection bias. Despite the reduced sample size, detection of significant superiority in the ID group was still achieved in many of the major immunogenicity outcomes. The larger sample size from the expanded analysis population confirmed the observed superior immunogenicity. This study focused on adolescents, and thus the immunogenicity of ID CoronaVac will need to be explored for adults and young children as well.

The specific populations that would likely benefit most from this enhanced immunogenicity of ID CoronaVac are the unvaccinated and high priority group that includes immunocompromised patients, young individuals with comorbidities, elderly, pregnant persons and frontline health workers. This aligns with the World Health Organization’s most recent recommendation on 28 March 2023 that this high priority group should receive an additional COVID-19 booster vaccination 6-12 months after the last dose. Many older adults, young children or those with debilitating chronic diseases are hesitant towards receiving the novel monovalent or bivalent mRNA vaccines due to systemic adverse effects, risks of myocarditis and potentially higher association with ischaemic stroke.^14, 42–44^ As this study demonstrated more frequent but tolerable local adverse effects only, it is possible these individuals would be more accepting of ID inactivated vaccines. ID can offer a cost-effective option for areas with limited access to more immunogenic vaccines that have higher financial or storage demands. A previous study revealed enhanced immunogenicity from ID against the hepatitis B virus in dialysis patients that was more striking 52 weeks after ID than IM.^45^ Whether such durability of heightened immunogenicity in healthy individuals is conferred by ID CoronaVac against SARS-CoV-2 and novel VOCs in the future is not yet certain, which we will track in these adolescents for the next 3 years. Importantly, this study serves as the basis for further research on ID using full doses of vaccines, rather than fractional dosing, to optimise protection against COVID-19 and other infectious diseases, particularly for those at risk of vaccine failures.

## METHODS

### Study Design

This registered study is part of the COVID-19 Vaccination in Adolescents and Children (COVAC) (Department of Health, HK, Clinical Trial Certificate 101894; clinicaltrials.gov NCT04800133) that investigates immunobridging for BNT162b2 and CoronaVac in adolescents and children, as previously described.^14, 16, 39^ The current pre-specified interim analysis aims to demonstrate the superiority in immunogenicity of the intradermal (ID) compared to intramuscular (IM) route of administration for CoronaVac in adolescents 11-17 years old and reports on the reactogenicity between ID and IM. The University of Hong Kong (HKU)/HK West Cluster Hospital Authority Institutional Review Board (UW21-157) approved the research procedures, which were in compliance with the October 2013 Declaration of Helsinki principles.

### Participants

Recruitment targeted 11- to 17-year-old adolescents residing in HK who were healthy or in stable condition. Potential participants were recruited using advertisements posted in schools and mass media. The exclusion criteria for this analysis included known history of COVID-19 (by self-reporting at any of the 4 study visits, or baseline S-RBD IgG or ORF8 IgG positivity at any visit), severe allergy, neuropsychiatric conditions, immunocompromised states, transfusion of blood products within 60 days, haemophilia, pregnancy or breastfeeding (see Protocol and Statistical Analysis Plan for details).

### Procedures

Study doctors obtained informed assent from eligible participants and consent from their respective parents or legally acceptable representatives. The skin superficial to the deltoid muscle was cleansed with 70% weight/volume isopropyl alcohol before using the standard 1 mL (KDL Medical, Shanghai, China) or MicronJet600 (NanoPass Technologies, Ness Ziona, Israel) needles for IM or ID, respectively, at a HK Community Vaccination Centre (CVC) (Supplementary Video).^46^ The dosage of CoronaVac was 0.5 mL (equivalent to 600 SU, or 3 µg, of the whole virus antigen of the inactivated SARS-CoV-2 CZ02 strain) for each injection, with a total of 3 separate doses given fo IM or ID. Doses 2 and 3 were given 28-35 days and 84 days after dose 1, respectively. Whole blood was obtained before doses 1 (baseline), 2 (C), 3 (CC) and post dose 3 (CCC) (see Analysis populations in Statistical Analyses of Methods).

#### Safety and reactogenicity data collection

Participants remained at the CVC for observation by the study nurse and doctor for at least 15 mins after each vaccine injection. Participants were required to report pre-specified adverse reactions (ARs) in an online or handwritten diary for the following 7 days, as previously described.^14, 16^ They were encouraged to capture photos of their sites of injection for at least 7 days and until resolution of local reactions, followed by uploading onto our online diary website. Unsolicited adverse events (AEs), such as hospitalisation, life-threatening illnesses, disabilities, deaths, birth defects of offspring and breakthrough COVID-19 are monitored for 3 years. These AEs were reviewed by study physicians, who assessed the probability of causal relationship with the study vaccination.

#### S-RBD IgG, surrogate virus neutralisation assay (sVNT), plaque reduction neutralisation titre (PRNT)

Peripheral blood was collected into clot activator vacutainer tubes, separated into serum and stored at −80° C. Sera were heat-inactivated at 56° C for 30 mins prior to testing. The SARS-CoV-2 S receptor-binding domain (S-RBD) IgG enzyme-linked immunosorbent assay (ELISA) and PRNT have been validated in our previous publications.^14, 16, 39, 47^ The sVNT was carried out according to the manufacturer’s instructions (GenScript Inc, Piscataway, USA) and our previous experiments, which have been validated.^14, 16, 39, 47^

Briefly, S-RBD IgG ELISA plates were coated overnight with 100 ng/well of purified recombinant S-RBD in PBS buffer, and then 100 μL Chonblock Blocking/Sample Dilution (CBSD) ELISA buffer (Chondrex Inc, Redmond, USA) were added. The incubation period of this mixture at room temperature (RT) was 2 hrs. Serum was tested at a dilution of 1:100 in CBSD ELISA buffer, then added to the wells for 2 hrs at 37°C. After washing with PBS that contained 0.2% Tween 20, horseradish peroxidase (HRP)-conjugated goat anti-human IgG (1:5000, Thermo Fisher Scientific) was added for 1 hr at 37°C and then washed 5 times with PBS containing 0.2% Tween 20. HRP substrate (Ncm TMB One, New Cell & Molecular Biotech Co. Ltd, China) of 100 μL was added for 15 mins. This reaction was stopped by 50 μL of 2 M H_2_SO_4_. The optical density (OD) was analysed in a Sunrise absorbance microplate reader (Tecan, Männedorf, Switzerland) at 450 nm wavelength. Each OD reading subtracted the background OD in PBS-coated control wells with the participant’s serum. Values at or above an OD450 of 0.5 were considered positive, while values below were imputed as 0.25.^14, 16, 39^

The sVNT was performed using 10 μL of each serum, positive and negative controls, which were diluted at 1:10 and mixed with an equal volume HRP conjugated to the wild type (WT) SARS-CoV-2 S-RBD (6 ng), and these were incubated for 30 mins at 37°C, then 100 μL of each sample was added to microtitre plate wells coated with angiotensin-converting enzyme-2 (ACE-2) receptor.^14, 16, 39^ This plate was sealed for 15 mins at 37°C, washed with wash-solution, tapped dry, and then 100 μL of 3,3’,5,5’-tetramethylbenzidine (TMB) was added. This mixture was incubated in the dark at RT for 15 mins. The reaction was terminated with 50 μL of Stop Solution and the absorbance was read at 450 nm in a microplate reader. After confirmation that the positive and negative controls provided the recommended OD450 values, the % inhibition of each serum was calculated as 1 - sample OD value/negative control OD value x100%. Inhibition (%) of at least 30%, the limit of quantification (LOQ), was regarded as positive, while values below 30% were imputed as 10%.^14, 16, 39^

The PRNT was performed in duplicate in a biosafety level 3 facility.^14, 47^ Serial dilutions of serum from 1:10 to at least 1:320 were incubated with approximately 30 plaque-forming units of the WT SARS-CoV-2 BetaCoV/Hong Kong/VM20001061/2020 virus in culture plates (Techno Plastic Products AG, Trasadingen, Switzerland) for 1 hr at 37°C.^14–16, 39^ These virus-serum mixtures were added onto Vero E6 cell monolayers and incubated for 1 hr at 37°C in a 5% CO_2_ incubator. The plates were overlaid with 1% agarose in cell culture medium and incubated for 3 days, and then the plates were fixed and stained. Antibody titres were defined as the reciprocal of the highest serum dilution that resulted in the more stringent cut-off of >90% (PRNT90) or >50% (PRNT50) reduction in the number of plaques. Values below the lowest dilution tested, which was 10, were imputed as 5, while those above 320 were imputed as 640.^14, 16, 39^

#### S IgG, avidity and FcγRIIIa-binding

S IgG, avidity and Fc*γ*RIIIa-binding assays were performed as previously described, with the addition of Omicron BA.2.^14, 16, 39, 48^ In brief, plates (Nunc MaxiSorp, Thermofisher Scientific) were coated with 250 ng/mL WT (AcroBiosystems) or Omicron BA.2 (AcroBiosystems) SARS-CoV-2 S protein for IgG and IgG avidity assessments, or 500 ng/mL WT (Sinobiological) or Omicron BA.2 (AcroBiosystems) S for FcγRIIIa-binding detection, or 300 ng/mL ORF8 (Masashi Mori, Ishiwaka University, Japan) at 37°C for 2 hrs. The protein for S IgG was diluted in PBS. The plates were blocked with 1% FBS in PBS for 1 hr, incubated with 1:100 heat-inactivated (HI) serum diluted in 0.05% Tween-20/ 0.1% FBS in PBS for 2 hrs at RT prior to rinsing. For antibody avidity, plates were washed thrice with 8M Urea before incubation for 2 hrs with IgG-HRP (1:5000; G8-185, BD). HRP was revealed by stabilized hydrogen peroxide and tetramethylbenzidine (R&D systems) for 20 mins and stopped with 2N H_2_SO_4_ before analysis with an absorbance microplate reader at 450 nm wavelength (Tecan Life Sciences). For those with a positive S IgG value, the IgG avidity index was calculated by the ratio of the OD450 values after to before washing of the plates, censored at 100%. Fc*γ*RIIIa-binding antibodies were detected after incubation with HI serum at 1:50 dilution for 1 hr at 37°C and then with biotinylated Fc*γ*RIIIa-V158, which was expressed in-house (from Mark Hogarth and Bruce Wines, Burnet Institute, Australia), at 100 ng/mL for 1 hr at 37°C. Streptavidin-HRP (1:10,000, Pierce) was added for detection of S specific Fc*γ*RIIIa-V158-binding antibodies. OD450 values at or above the respective limits of detection (LODs) were considered positive, while values below were imputed as 0.5 of the LOD.^14, 16, 39^

#### T cell responses

Density gradient separation was performed to isolate peripheral blood mononuclear cells (PBMCs) from whole blood, which was frozen in liquid nitrogen.^14, 16, 39^ Subsequently, thawed PBMCs were rested for 2 hrs in RPMI medium supplemented with 10% human AB serum. The cells were stimulated with sterile double-distilled water (ddH2O) or 1 µg/mL overlapping peptide pools representing the WT SARS-CoV-2 S, N (nucleocapsid) and M (membrane) proteins, or Omicron B.1.1.529/BA.1 S mutation pool and WT reference pool, BA.1 N mutation pool and WT N reference pool, BA.1 M mutation pool and WT M reference pool (Miltenyi Biotec, Bergisch Gladbach, Germany) (synthesized by ChinaPeptides Co., Ltd, as previously described) for 16 hrs in 1 µg/mL anti-CD28 and anti-CD49d costimulatory antibodies (clones CD28.2 and 9F10, Biolegend, San Diego, USA).^14, 16, 39^ This mixture was stimulated for 2 hrs, followed by the addition of 10 µg/mL brefeldin A (Sigma, Kawasaki, Japan).^14, 16, 39, 49^ The cells were then washed and subjected to immunostaining with a fixable viability dye (eBioscience, Santa Clara, USA, 1:60) and antibodies against CD3^+^ (HIT3a, 1:60), CD4^+^ (OKT4, 1:60), CD8^+^ (HIT8a, 1:60), IFN-*γ* (B27, 1:15) and IL-2 (MQ1-17H12, 1:15) antibodies (Biolegend, San Diego, USA). Flow cytometry (LSR II with FACSDiva version 8.0, BD Biosciences, Franklin Lakes, USA), analysed by Flowjo version 10 software (BD, Ashland, USA), was used for data acquisition. Antigen-specific T cell results were finalised after subtracting the background (ddH2O) data and presented as the percentage of CD4^+^ or CD8^+^ T cells.^14, 16, 39, 50^ The T cell response against a single peptide pool was considered positive when the frequency of cytokine-expressing cells was higher than 0.005% and the stimulation index was higher than 2, while negative values were imputed as 0.0025%.^14, 16, 39^ Total T cell responses against S, N and M peptide pools were added together, which used a cut-off of 0.01%.^14, 16, 39^

### Outcomes

The primary outcomes in this interim analysis were humoral immunogenicity (S IgG and S-RBD IgG levels, sVNT %inhibition, 90% and 50% PRNT titres, S IgG avidity and Fc*γ*RIIIa-binding) and cellular immunogenicity markers (S, N- and M-specific IFN-*γ*^+^ and IL-2^+^ CD4^+^ and CD8^+^ T cell responses measured by the flow-cytometry-based intracellular cytokine staining assay) 13-42 days after doses 2 and 3 of CoronaVac. The primary reactogenicity outcomes included pre-specified ARs and reported antipyretic use during the 7 days following each vaccine injection.

Omicron humoral and cellular immunogenicity results were secondary outcomes. Regarding safety, the secondary outcomes were unsolicited AEs within 28 days after each vaccine injection and SAEs during the entire study period. The Protocol and Statistical Analysis Plan (Supplementary Information) described other secondary outcomes that were not pertinent to this interim analysis, such as specific assessments for participants with known chronic illnesses.

### Statistical Analyses

#### Power analyses and sample size estimation

For the primary immunogenicity objectives, when comparing the peak geometric mean (GM) immunogenicity outcomes of seroconversion rate or adverse events for CoronaVac ID with that of IM administration in adolescents aged 11-17 years, a sample size of 50 in each group would assure that a two-sided test with *α*=0.05 has 97% power to detect an effect size with a Cohen’s d value=0.78, or a difference of 0.51 after natural logarithmic transformation, between the 2 groups, with a standard deviation (SD) of 0.65 within each group. We aimed to recruit 60 participants for each group of IM and ID administrations to accommodate for potential attrition or protocol deviation. However, performance of assays requiring large blood volumes was omitted for a few younger, small-sized adolescents, who could provide limited amounts of blood. In terms of the proportion of participants with a positive result in immunogenicity outcomes or ARs, 50 adolescents would yield a 95% chance to detect the true value within ±11 percentage points of the measured percentage, assuming a prevalence of 80%. G*Power (Heinrich-Heine-Universität Düsseldorf, Düsseldorf, Germany) and Sampsize (sampsize.sourceforge.net) were used for these power analyses.

#### Analysis populations

The primary analysis of humoral and cellular immunogenicity outcomes was performed in the healthy adolescent participants who received IM or ID injections of CoronaVac on a per-protocol basis. The evaluable analysis population included participants who were generally healthy and have remained uninfected during study visits (based on self-reporting, ORF8 IgG negativity and negative baseline S-RBD IgG), no major protocol deviations, received dose 3 at least 84 days after dose 1, blood sampling within the evaluable window for post-dose 1 (no more than 3 days earlier or later than day 28, and before dose 2), post-dose 2 (within day 13-42 post-dose 2 and before any further doses), within days 13-42 post-dose 3 and had valid results for the relevant analysis and timepoints (see Protocol in Supplementary Information). The expanded analysis population included similar criteria as the evaluable analysis population except the requirement of a valid immunogenicity result for the particular analysis at least 14 days post-dose 1 but before dose 2 and between 7-56 days post-dose 2 (see Protocol in Supplementary Information). The superiority and non-inferiority hypotheses testing for primary immunogenicity outcomes included participants aged 11-17 years in the adolescent groups who received IM or ID injections of CoronaVac for doses 1-3.

#### Statistical tests

Immunogenicity outcome data below the cut-off were imputed with half the cut-off value. GMs were calculated for each immunogenicity outcome, timepoint and group. GM ratios (GMRs) were calculated as exponentiated differences between the means of the natural logarithmic-transformed immunogenicity outcomes between groups. The GMRs were reported with a two-sided 95%CI for testing the superiority hypothesis with the lower bound of the 95%CI for GMR >1. Additionally, confirmation of the superiority results was performed in the expanded analysis population. Simultaneously, we conducted a non-inferiority analysis at the non-inferiority margin of 0.60 for immunogenicity outcomes since it was possible that superiority for a few immunogenicity outcomes would not be satisfied. By convention, results were regarded as inconclusive if both non-inferiority and inferiority were not met. Comparisons of immunogenicity outcomes between groups were performed with unpaired *t* test after natural logarithmic transformation. The proportion of participants with a positive result was reported in percentages, with two-sided 95%CI derived from Clopper-Pearson method. The Fisher exact test was used for comparisons of proportions between groups.

Reactogenicity and safety were assessed in the participants who remained generally healthy, uninfected and contributed any AR or AE data after the 3 doses and before the study database was locked for the current interim analysis in these adolescent groups who received IM or ID CoronaVac that comprised the healthy safety population. For the primary reactogenicity analysis, the proportions of participants reporting each AR at the maximum severity and antipyretic use within 7 days after each vaccine injection were reported in percentages, with the 95%CI derived using the Clopper-Pearson method. ARs of all severity and antipyretic use were compared between vaccine routes of administration by the Fisher exact test. The incidences of AEs by severity and SAEs that were reported by the post-dose 3 study visit (28 days post-dose 3) were presented as counts and events-per-participant by the vaccine route of administration.

Data analyses and graphing were performed using GraphPad Prism (version 9.4.0). Two-sided 95%CIs were presented for all outcomes unless otherwise stated.

#### Vaccine efficacy estimates

Vaccine efficacies were estimated as a secondary objective by correlations with neutralising antibody titres, as described in our previous publication.^14, 16, 31^ In brief, GMTs of PRNT50 in the evaluable analysis populations were divided from that of 102 convalescent sera collected on days 28-59 post-onset of illness in patients aged ≥18 years to calculate the mean neutralising levels. The best fit of the logistic model, generated from the online plot digitizer tool (https://automeris.io/WebPlotDigitizer/, version 4.5), was used to extrapolate a single point VE estimate for each route of vaccine administration.

## Supporting information

Protocol

Statistical Analysis Plan

## Data Availability

The study Protocol and Statistical Analysis Plan are included in the Supplementary Information. To maintain confidentiality for the participants, only deidentified data shall be shared with scientific investigators who submit a justifiable inquiry to. The study is ongoing, and therefore data will be available upon request 1 month after the completion of the study in 2025.

## ACKNOWLEDGEMENTS

We express our appreciation to the staff members at the CVC, laboratory personnel and clinical research team within the Department of Paediatrics and Adolescent Medicine, health care workers and participants who volunteered for this project. We are grateful to Professor Mark Hogarth and Dr Bruce Wines from the Burnet Institute in Australia for the in-house expressed biotinylated Fc*γ*RIIIa-V158. The study was funded by the research grants COVID19F12, COVID19F10 and COVID19F02 awarded to Y.L. Lau by the Health Bureau of the Government of Hong Kong, while Issan Y.S. Tam was partly supported by a donation in memory of Dr. Ton Lung Quong and Reverend Marion Quong. The research work was supported additionally by grants from the Health and Medical Research Fund Commissioned Research on the Novel Coronavirus Disease (COVID-19), Hong Kong SAR (COVID190115) and the Theme-based Research Scheme of the Research Grants Council of the Hong Kong Special Administrative Region, China (T11-712/19-N and T11-705/21-N). The funding sources were not involved in the study design, data collection, laboratory assays, statistical computation, interpretation or final conclusions of this project.

## AUTHOR CONTRIBUTIONS STATEMENT

Y.L. Lau conceptualized the study. Y.L. Lau, W. Tu, M. Peiris, J.S. Rosa Duque and D. Leung designed the study. Y.L. Lau led the acquisition of funding. Y.L. Lau, W. Tu, M. Peiris and S. Valkenburg supervised the project. S.M. Chan, J.S. Rosa Duque, S.M.S. Cheng, D. Leung, X. Mu, X. Wang and J.H.Y. Lam led the study administrative procedures. W.H.S. Wong provided software support. S.M. Chan, J.S. Rosa Duque, Y.L. Lau and W.H.S. Wong contributed to recruitment of participants. Y.L. Lau, J.S. Rosa Duque and S.M. Chan provided clinical assessments and follow-up. J.S. Rosa Duque, Y.L. Lau, D. Leung, S.M. Chan, and J.H.Y. Lam collected safety data. S.M.S. Cheng, M. Peiris, L.C.H. Tsang, S. Chaothai, T.C. Kwan, J.K.C. Li, K.C.K. Chan, L.L.H. Luk and J.C.H. Ho developed and performed S-RBD IgG, sVNT and neutralisation antibody assays. C.A. Cohen and S. Valkenburg developed and performed the S IgG, IgG avidity, S IgG Fc*γ*RIIIa-binding and ORF8 antibody assays. M. Mori provided and developed the specialised ORF8 protein. W. Tu, X. Wang, X. Mu, Y. Zhang, W. Zhang, Y. Chung, M. Wang, I.Y.S Tam, W.Y. Li and A.M.T. Lee developed, performed and supervised the T cell assays. J.S. Rosa Duque, D. Leung, J.H.Y. Lam and T.M. Lau curated and analysed the data. D. Leung, J.S. Rosa Duque, J.H.Y. Lam, S.M.S. Cheng, Y.L. Lau, M. Peiris and T.M. Lau performed the vaccine efficacy extrapolation. J.S. Rosa Duque, D. Leung, Y.L. Lau, J.H.Y. Lam and T.M. Lau visualized the data. J.S. Rosa Duque, D. Leung, J.H.Y. Lam, S.M.S. Cheng, C.A. Cohen, I.Y.S Tam and T.M. Lau validated the data. J.S. Rosa Duque wrote the first draft as supervised by Y.L. Lau, W. Tu, M. Peiris and S.A. Valkenburg, with input from S.M.S. Cheng, C.A. Cohen, D. Leung and X. Wang. All authors contributed to the article and approved the submitted version.

## DATA AVAILABILITY

The study’s Protocol and Statistical Analysis Plan are included in the Supplementary Information. To maintain confidentiality for the participants, only deidentified data shall be shared with scientific investigators who submit a justifiable inquiry to. The study is ongoing, and therefore data will be available upon request 1 month after the completion of the study in 2025.

## REFERENCES

1. Tso, W.W., et al. Severity of SARS-CoV-2 Omicron BA. 2 infection in unvaccinated hospitalized children: comparison to influenza and parainfluenza infections. Emerg Microbes Infect (2022).

2. McCrindle, B.W., et al. SARS-CoV-2 variants and multisystem inflammatory syndrome in children. N Engl J Med (2023).

3. Al-Aly, Z., Bowe, B. & Xie, Y. Long COVID after breakthrough SARS-CoV-2 infection. Nat Med (2022).

4. Xiao, Y., Yip, P.S., Pathak, J. & Mann, J.J. Association of social determinants of health and vaccinations with child mental health during the COVID-19 pandemic in the US. JAMA Psychiatry 79, 610–621 (2022).

5. Rosa Duque, J.S., et al. COVID-19 vaccines versus pediatric hospitalization. Cell Rep Med 4, 100936 (2023).

6. Polack, F.P., et al. Safety and efficacy of the BNT162b2 mRNA COVID-19 vaccine. N Engl J Med 383, 2603–2615 (2020).

7. Palacios, R., et al. Efficacy and safety of a COVID-19 inactivated vaccine in healthcare professionals in Brazil: the PROFISCOV study. SSRN http://dx.doi.org/10.2139/ssrn.3822780 (2021).

8. Tanriover, M.D., et al. Efficacy and safety of an inactivated whole-virion SARS-CoV-2 vaccine (CoronaVac): interim results of a double-blind, randomised, placebo-controlled, phase 3 trial in Turkey. The Lancet 398, 213–222 (2021).

9. Frenck, R.W., Jr., et al. Safety, immunogenicity, and efficacy of the BNT162b2 COVID-19 vaccine in adolescents. N Engl J Med 385, 239–250 (2021).

10. Han, B., et al. Safety, tolerability, and immunogenicity of an inactivated SARS-CoV-2 vaccine (CoronaVac) in healthy children and adolescents: a double-blind, randomised, controlled, phase 1/2 clinical trial. The Lancet Infectious Diseases 21, 1645–1653 (2021).

11. Jara, A., et al. Effectiveness of an inactivated SARS-CoV-2 vaccine in children and adolescents: a large-scale observational study. SSRN http://dx.doi.org/10.2139/ssrn.4035405 (2022).

12. Leung, D., et al. Effectiveness of BNT162b2 and CoronaVac in children and adolescents against SARS-CoV-2 infection during Omicron BA.2 wave in Hong Kong. Commun Med (Lond) 3, 3 (2023).

13. Mallapaty, S., et al. How COVID vaccines shaped 2021 in eight powerful charts. Nature 600, 580–583 (2021).

14. Rosa Duque, J.S., et al. Immunogenicity and reactogenicity of SARS-CoV-2 vaccines BNT162b2 and CoronaVac in healthy adolescents. Nat Commun 13, 4798 (2022).

15. Cheng, S.M.S., et al. Neutralizing antibodies against the SARS-CoV-2 Omicron variant BA.1 following homologous and heterologous CoronaVac or BNT162b2 vaccination. Nat Med 28, 486–489 (2022).

16. Leung, D., et al. Immunogenicity against wild-type and Omicron SARS-CoV-2 after a third dose of inactivated COVID-19 vaccine in healthy adolescents. Frontiers in Immunology 14(2023).

17. Egunsola, O., et al. Immunogenicity and safety of reduced-dose intradermal vs intramuscular influenza vaccines: a systematic review and meta-analysis. JAMA Netw Open 4, e2035693 (2021).

18. Haniffa, M., et al. Human tissues contain CD141hi cross-presenting dendritic cells with functional homology to mouse CD103+ nonlymphoid dendritic cells. Immunity 37, 60–73 (2012).

19. Kashem, S.W., Haniffa, M. & Kaplan, D.H. Antigen-presenting cells in the skin. Annu Rev Immunol 35, 469–499 (2017).

20. St John, A.L., Rathore, A.P.S. & Ginhoux, F. New perspectives on the origins and heterogeneity of mast cells. Nat Rev Immunol (2022).

21. Palucka, K., Banchereau, J. & Mellman, I. Designing vaccines based on biology of human dendritic cell subsets. Immunity 33, 464–478 (2010).

22. Snider, C.J., et al. Immunogenicity of full and fractional dose of inactivated poliovirus vaccine for use in routine immunisation and outbreak response: an open-label, randomised controlled trial. The Lancet 393, 2624–2634 (2019).

23. Egemen, A., et al. Low-dose intradermal versus intramuscular administration of recombinant hepatitis B vaccine: a comparison of immunogenicity in infants and preschool children. Vaccine 16, 1511–1515 (1998).

24. Nelson, E.A., et al. A pilot randomized study to assess immunogenicity, reactogenicity, safety and tolerability of two human papillomavirus vaccines administered intramuscularly and intradermally to females aged 18-26 years. Vaccine 31, 3452–3460 (2013).

25. Chiu, S.S., Peiris, J.S., Chan, K.H., Wong, W.H. & Lau, Y.L. Immunogenicity and safety of intradermal influenza immunization at a reduced dose in healthy children. Pediatrics 119, 1076–1082 (2007).

26. Chiu, S.S., Chan, K.H., Tu, W., Lau, Y.L. & Peiris, J.S. Immunogenicity and safety of intradermal versus intramuscular route of influenza immunization in infants less than 6 months of age: a randomized controlled trial. Vaccine 27, 4834–4839 (2009).

27. Arnou, R., et al. Intradermal influenza vaccine for older adults: a randomized controlled multicenter phase III study. Vaccine 27, 7304–7312 (2009).

28. Holland, D., et al. Intradermal influenza vaccine administered using a new microinjection system produces superior immunogenicity in elderly adults: a randomized controlled trial. J Infect Dis 198, 650–658 (2008).

29. Pinpathomrat, N., et al. Immunogenicity and safety of an intradermal ChAdOx1 nCoV-19 boost in a healthy population. NPJ Vaccines 7, 52 (2022).

30. Chalermphanchai, N., et al. Safety, tolerability, and antibody response after intradermal vaccination of PFE-BNT in adults who have completed two-doses of Verocell (inactivated vaccine). Vaccine X 10, 100148 (2022).

31. Khoury, D.S., et al. Neutralizing antibody levels are highly predictive of immune protection from symptomatic SARS-CoV-2 infection. Nat Med 27, 1205–1211 (2021).

32. Guo, L., et al. SARS-CoV-2-specific antibody and T-cell responses 1 year after infection in people recovered from COVID-19: a longitudinal cohort study. Lancet Microbe 3, e348–e356 (2022).

33. Wilkinson, T.M., et al. Preexisting influenza-specific CD4+ T cells correlate with disease protection against influenza challenge in humans. Nat Med 18, 274–280 (2012).

34. Hayward, A.C., et al. Natural T cell-mediated protection against seasonal and pandemic influenza. Results of the Flu Watch Cohort Study. Am J Respir Crit Care Med 191, 1422–1431 (2015).

35. Sridhar, S., et al. Cellular immune correlates of protection against symptomatic pandemic influenza. Nat Med 19, 1305–1312 (2013).

36. Tsang, T.K., et al. Investigation of CD4 and CD8 T cell-mediated protection against influenza A virus in a cohort study. BMC Med 20, 230 (2022).

37. Chen, J., et al. DNA vaccines expressing the envelope and membrane proteins provide partial protection against SARS-CoV-2 in mice. Front Immunol 13, 827605 (2022).

38. Nakagama, Y., et al. Antibody avidity maturation, following recovery from infection or the booster vaccination, grants breadth in SARS-CoV-2 neutralizing capacity. J Infect Dis (2022).

39. Mu, X., et al. Antibody and T cell responses against wild-type and Omicron SARS-CoV-2 after third-dose BNT162b2 in adolescents. Signal Transduct Target Ther 7, 397 (2022).

40. Gagne, M., et al. Protection from SARS-CoV-2 Delta one year after mRNA-1273 vaccination in rhesus macaques coincides with anamnestic antibody response in the lung. Cell 185, 113–130.e115 (2022).

41. Frey, S.E., Goll, J.B. & Beigel, J.H. Erythema and induration after Mpox (JYNNEOS) vaccination revisited. N Engl J Med (2023).

42. Chua, G.T., et al. Epidemiology of acute myocarditis/pericarditis in Hong Kong adolescents following Comirnaty vaccination. Clin Infect Dis 75, 673–681 (2022).

43. Hause, A.M., et al. Safety Monitoring of Bivalent COVID-19 mRNA Vaccine Booster Doses Among Persons Aged ≥12 Years - United States, August 31-October 23, 2022. MMWR Morb Mortal Wkly Rep 71, 1401–1406 (2022).

44. Yu, M.K.L., et al. Hesitancy, reactogenicity and immunogenicity of the mRNA and whole-virus inactivated Covid-19 vaccines in pediatric neuromuscular diseases. Hum Vaccin Immunother in press (2023).

45. Hung, I.F., et al. A double-blind, randomized phase 2 controlled trial of intradermal hepatitis b vaccination with a topical toll-like receptor 7 agonist imiquimod, in patients on dialysis. Clin Infect Dis 73, e304–e311 (2021).

46. Beals, C.R., et al. Immune response and reactogenicity of intradermal administration versus subcutaneous administration of varicella-zoster virus vaccine: an exploratory, randomised, partly blinded trial. The Lancet Infectious Diseases 16, 915–922 (2016).

47. Perera, R.A., et al. Serological assays for severe acute respiratory syndrome coronavirus 2 (SARS-CoV-2), March 2020. Euro Surveill 25(2020).

48. Mok, C.K.P., et al. Comparison of the immunogenicity of BNT162b2 and CoronaVac COVID-19 vaccines in Hong Kong. Respirology 4, 301–310 (2021).

49. Sattler, A., et al. Impaired humoral and cellular immunity after SARS-CoV-2 BNT162b2 (tozinameran) prime-boost vaccination in kidney transplant recipients. J Clin Invest 131(2021).

50. Mateus, J., et al. Low-dose mRNA-1273 COVID-19 vaccine generates durable memory enhanced by cross-reactive T cells. Science 374, eabj9853 (2021).

