## Supplementary Tables and Figures for "Superior antibody and membrane protein-specific T cell responses to CoronaVac by intradermal versus intramuscular routes in adolescents"

| **Supp. Table 1. Characteristics of the study participants in the healthy safety population.** | |
|---|---|
| **Intradermal** | |
| **Numbers of participants** | 59 |
| **Male sex** | 30 (50.8%) |
| **Han Chinese** | 59 (100%) |
| **Age (years)** | 13.7 (1.53) |
| **Intramuscular** | |
| **Number of participants** | 119 |
| **Male sex** | 65 (54.6%) |
| **Han Chinese** | 119 (100%) |
| **Age (years)** | 14.3 (17.60) |
| Values are either counts (%) or means (standard deviations). | |

**Supp. Table 2. Humoral immunogenicity outcomes against wild type SARS-CoV-2 post-dose 2 and post-dose 3 of CoronaVac in the expanded analysis population.**

| | **Intramuscular** | | **Intradermal** | |
|---|---|---|---|---|
| | **2 doses** | **3 doses** | **2 doses** | **3 doses** |
| **S IgG on ELISA** | | | | |
| N | 116 | 77 | 47 | 37 |
| GM OD450 value (95% CI) | 0.536 (0.493-0.582) | 0.872 (0.785-0.968) | 0.634 (0.562-0.716) | 1.05 (0.983-1.12) |
| % positive (>/=LOD at 0.3) | 94.0% | 97.4% | 95.7%, *P*>0.9999 | 100%, *P*>0.9999 |
| **S-RBD IgG on ELISA** | | | | |
| N | 119 | 82 | 59 | 43 |
| GM OD450 value (95% CI) | 1.20 (1.10-1.31) | 1.69 (1.60-1.78) | 2.15 (2.03-2.28) | 2.43 (2.31-2.56) |
| % positive (>/=LOD at 0.5) | 96.6% | 100% | 100%, *P*=0.303 | 100%, *P*>0.9999 |
| **S-RBD ACE2-blocking antibody on sVNT** | | | | |
| N | 119 | 82 | 59 | 43 |
| GM % inhibition (95% CI) | 71.2 (66.7-76.0) | 84.0 (81.0-87.1) | 78.2 (74.5-82.1) | 90.7 (87.5-94.0) |
| % positive (>/=LOQ at 30%) | 96.6% | 100% | 100%, *P*=0.303 | 100%, *P*>0.9999 |
| **Neutralising antibody on PRNT** | | | | |
| N | 119 | 82 | 59 | 43 |
| GM PRNT90 (95% CI) | 9.83 (8.67-11.1) | 18.2 (14.5-22.9) | 10.9 (8.39-14.1) | 37.5 (26.8-52.4) |
| % positive (>/=LOD at 10) | 65.6% | 80.5% | 62.7%, *P*=0.741 | 97.7%, *P*=0.0061 |
| GM PRNT50 (95% CI) | 26.8 (23.0-31.1) | 54.2 (44.3-66.5) | 30.5 (24.1-38.6) | 105 (78.9-140) |
| % positive (>/=LOD at 10) | 96.6% | 100% | 98.3%, *P*>0.9999 | 100%, *P*>0.9999 |
| **S IgG avidity on ELISA** | | | | |
| N | 109 | 75 | 45 | 37 |
| GM avidity index (95% CI) | 20.5 (19.1-22.1) | 35.6 (32.5-39.1) | 6.95 (5.03-9.60) | 52.6 (47.7-58.1) |
| **S IgG FcγRIIIa-binding on ELISA** | | | | |
| N | 116 | 77 | 47 | 37 |
| GM OD450 value (95% CI) | 0.749 (0.649-0.864) | 1.30 (1.14-1.49) | 1.10 (0.955-1.27) | 1.78 (1.70-1.87) |
| % positive (>/=LOD at 0.28) | 87.1% | 97.4% | 95.7%, *P*=0.156 | 100%, *P*>0.9999 |

GM, geometric mean; OD, optical density; LOD, limit of detection; LOQ, limit of quantification; CI, confidence interval; S, spike protein; ELISA, enzyme-linked immunosorbent assay; RBD, receptor-binding domain; ACE-2, angiotensin-converting enzyme-2; sVNT, surrogate virus neutralisation test; PRNT, plaque reduction neutralisation titre; PRNT90, 90% plaque reduction neutralisation titre; PRNT50, 50% plaque reduction neutralisation titre; FcγRIIIa, Fc gamma receptor III-a. *P*-values compare the proportion of positive responses between intramuscular and intradermal administration by Fisher's exact test.

| **Supp. Table 3. Cellular immunogenicity outcomes against wild type SARS-CoV-2 S, N and M peptide pools post-dose 2 and post-dose 3 of CoronaVac in the expanded analysis population.** | | | | |
|---|---|---|---|---|
| | **Intramuscular** | | **Intradermal** | |
| | **2 doses** | **3 doses** | **2 doses** | **3 doses** |
| **Total S, N, M-specific T cell responses on flow cytometry** | | | | |
| N | 60 | 69 | 48 | 41 |
| **GM** % IFN-$\gamma^+$CD4$^+$ T cells (95% CI) | 0.0581% (0.0405-0.08335%) | 0.0682% (0.0445-0.104%) | 0.107% (0.0628-0.183%) | 0.107% (0.0560-0.204%) |
| % positive (>/=cut-off at 0.0075%) | 83.3% | 73.9% | 81.3%, *p*=0.804 | 78.1%, *p*=0.656 |
| **GM** % IL-2$^+$CD4$^+$ T cells (95% CI) | 0.0395% (0.0301-0.0517%) | 0.0788% (0.0552-0.112%) | 0.112% (0.0680-0.183%) | 0.142% (0.0778-0.258%) |
| % positive (>/=cut-off at 0.0075%) | 83.3% | 81.2% | 79.2%, *p*=0.624 | 80.5%, *p*>0.9999 |
| **GM** % IFN-$\gamma^+$CD8$^+$ T cells (95% CI) | 0.0501% (0.0328-0.0766%) | 0.0642% (0.0384-0.107%) | 0.0589% (0.0333-0.104%) | 0.0635% (0.0327-0.124%) |
| % positive (>/=cut-off at 0.0075%) | 65.0% | 59.4% | 62.5%, *P*=0.842 | 63.4%, *p*=0.692 |
| **GM** % IL-2$^+$CD8$^+$ T cells (95% CI) | 0.0173% (0.0138-0.0216%) | 0.0387% (0.0262-0.0570%) | 0.0498% (0.0309-0.803%) | 0.0670% (0.0353-0.127%) |
| % positive (>/=cut-off at 0.0075%) | 58.3% | 60.9% | 68.8%, *p*=0.318 | 65.9%, *p*=0.685 |
| **S-specific T cell responses on flow cytometry** | | | | |
| N | 60 | 70 | 48 | 42 |
| **GM** % IFN-$\gamma^+$CD4$^+$ T cells (95% CI) | 0.0233% (0.0149-0.0362%) | 0.0153% (0.00958-0.0245%) | 0.0220% (0.0120-0.0405%) | 0.0266% (0.0138-0.0510%) |
| % positive (>/=cut-off at 0.005%) | 70.0% | 54.3% | 58.3%, *P*=0.229 | 69.1%, *P*=0.164 |
| **GM** % IL-2$^+$CD4$^+$ T cells (95% CI) | 0.0147% (0.0106-0.0204%) | 0.0177% (0.0114-0.0274%) | 0.0205% (0.0119-0.0353%) | 0.0320% (0.0172-0.0594%) |
| % positive (>/=cut-off at 0.005%) | 73.3% | 60.0% | 60.4%, *P*=0.214 | 69.1%, *P*= 0.419 |
| **GM** % IFN-$\gamma^+$CD8$^+$ T cells (95% CI) | 0.0142% (0.00861-0.0235%) | 0.0169% (0.00990-0.0289%) | 0.0138% (0.00772-0.0247%) | 0.0105% (0.00559-0.0197%) |
| % positive (>/=cut-off at 0.005%) | 48.3% | 48.6% | 45.8%, *P*=0.848 | 40.5%, *P*=0.439 |
| **GM** % IL-2$^+$CD8$^+$ T cells (95% CI) | 0.00625% (0.00476-0.00822%) | 0.00969 (0.00634-0.0148%) | 0.0121% (0.00727-0.0202%) | 0.0101% (0.00546-0.0185%) |
| % positive (>/=cut-off at 0.005%) | 48.3% | 45.7% | 52.1%, *P*=0.847 | 41.5%, *P*=0.696 |
| **N-specific T cell responses on flow cytometry** | | | | |
| N | 60 | 69 | 48 | 41 |
| **GM** % IFN-$\gamma^+$CD4$^+$ T cells (95% CI) | 0.0113% (0.00752-0.0170%) | 0.0132% (0.00816-0.0215%) | 0.0222% (0.0114-0.0433%) | 0.0221% (0.0111-0.0441%) |
| % positive (>/=cut-off at 0.005%) | 55.0% | 50.7% | 54.2%, *P*>0.9999 | 58.5%, *P*=0.438 |
| **GM** % IL-2$^+$CD4$^+$ T cells (95% CI) | 0.00126% (0.00899-0.0176%) | 0.0190% (0.0123-0.0294%) | 0.0281% (0.0149-0.0531%) | 0.0281% (0.0142-0.0553%) |
| % positive (>/=cut-off at 0.005%) | 66.7% | 60.9% | 60.4%, *P*=0.549 | 63.4%, *P*=0.841 |
| **GM** % IFN-$\gamma^+$CD8$^+$ T cells (95% CI) | 0.00767% (0.00485-0.0121%) | 0.0132% (0.00760-0.0230%) | 0.0163% (0.00842-0.0314%) | 0.0121% (0.00597-0.0246%) |
| % positive (>/=cut-off at 0.005%) | 31.7% | 39.1% | 43.8%, *P*=0.232 | 39.0%, *P*>0.9999 |
| **GM** % IL-2$^+$CD8$^+$ T cells (95% CI) | 0.00419% (0.00330-0.00532%) | 0.00893% (0.00583-0.0137%) | 0.0135% (0.00754-0.0241%) | 0.0136% (0.00715-0.0257%) |
| % positive (>/=cut-off at 0.005%) | 28.3% | 37.7% | 47.9%, *P*=0.0457 | 48.8%, *P*=0.318 |
| **M-specific T cell responses on flow cytometry** | | | | |
| N | 60 | 70 | 48 | 41 |
| **GM** % IFN-$\gamma^+$CD4$^+$ T cells (95% CI) | 0.00706% (0.00483-0.0103%) | 0.00628% (0.00416-0.00947%) | 0.00827% (0.00479-0.0143%) | 0.0106% (0.00503-0.0221%) |
| % positive (>/=cut-off at 0.005%) | 36.7% | 25.7% | 35.4%, *P*>0.9999 | 34.2%, *P*=0.389 |
| **GM** % IL-2$^+$CD4$^+$ T cells (95% CI) | 0.00571% (0.00449-0.00728%) | 0.00623% (0.00427-0.00909%) | 0.0107% (0.00617-0.0187%) | 0.0138% (0.00667-0.0287%) |
| % positive (>/=cut-off at 0.005%) | 46.7% | 27.1% | 41.7%, *P*=0.698 | 46.3%, *P*=0.0613 |
| **GM** % IFN-$\gamma^+$CD8$^+$ T cells (95% CI) | 0.00588% (0.00390-0.00886%) | 0.00453% (0.00302-0.00678%) | 0.00661% (0.00399-0.0110%) | 0.0116% (0.00589-0.0230%) |
| % positive (>/=cut-off at 0.005%) | 25.0% | 11.4% | 29.2%, *P*=0.667 | 41.5%, *P*=0.0004 |
| **GM** % IL-2$^+$CD8$^+$ T cells (95% CI) | 0.00383% (0.00309-0.00475%) | 0.00414% (0.00308-0.00556%) | 0.00521% (0.00348-0.00778%) | 0.0115% (0.00578-0.0229%) |
| % positive (>/=cut-off at 0.005%) | 23.3% | 15.7% | 27.1%, *P*=0.662 | 46.3%, *P*=0.0008 |

GM, geometric mean; CI, confidence interval; S, spike; N, nucleocapsid protein; M, membrane protein; IFN-$\gamma$, interferon-gamma; IL-2, interleukin-2. *P*-values compare the proportion of positive responses between intramuscular and intradermal administration by Fisher's exact test.

| **Supp. Table 4. Unsolicited adverse events within 28 days of vaccination in the healthy safety population.** | | | |
|---|---|---|---|
| | **Intramuscular (*N*=123)** | **Intradermal (*N*=59)** | **Overall (*N*=182)** |
| **Summary of adverse events and severe adverse events** | | | |
| **Any adverse event** | 8 (0.065) | 5 (0.085) | 13 (0.071) |
| **Grade 1** | 8 (0.065) | 4 (0.068) | 12 (0.066) |
| **Grade 2** | 0 (0.000) | 0 (0.000) | 0 (0.000) |
| **Grade 3** | 0 (0.000) | 1 (0.017) | 1 (0.005) |
| **Severe** | 0 (0.000) | 0 (0.000) | 0 (0.000) |
| Data are number of events (events per participant). N, total number of participants in the healthy safety population. | | | |

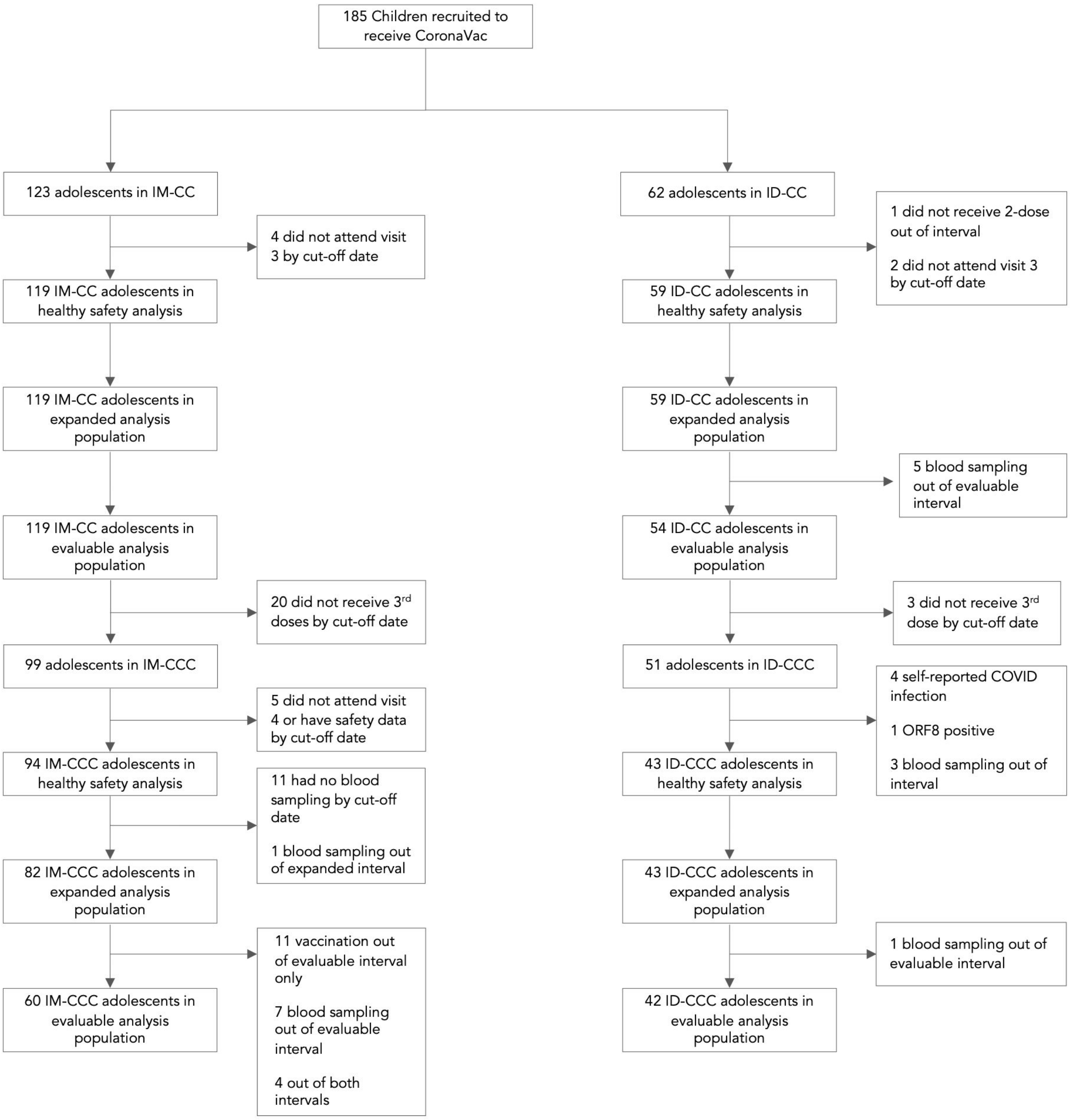


**Supp. Fig. 1. Study flow diagram.** The evaluable analysis population comprised of uninfected participants without major protocol deviations and valid results. For confirmation of results, an expanded analysis population with more relaxed time windows between doses and blood sample were included and analysed additionally. IM-CC and IM-CCC, 2 and 3 doses of vaccine administered intramuscularly, respectively; ID-CC and ID-CCC, 2 and 3 doses of vaccine administered intradermally, respectively; ORF8 IgG (a serological marker of past natural SARS-CoV-2 infection).

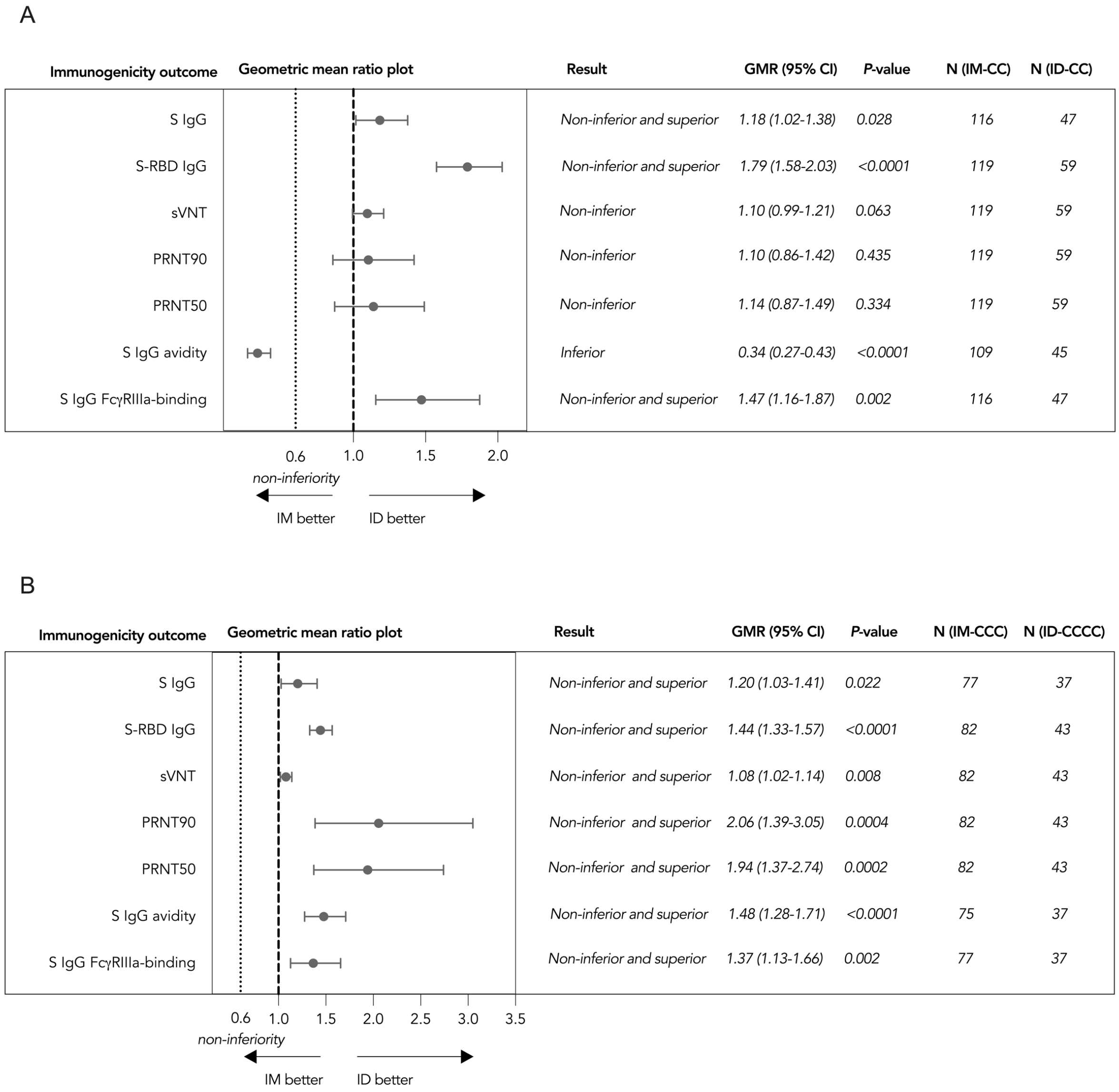


**Supp. Fig. 2. Superiority and non-inferiority hypotheses testing of humoral immunogenicity against wild type SARS-CoV-2 post-dose 2 and post-dose 3 of vaccination the in expanded analysis population.** Adolescents receiving (A) 2 doses of CoronaVac administered intramuscularly or intradermally and (B) 3 doses of CoronaVac administered intramuscularly or intradermally were tested for humoral immunogenicity outcomes in the expanded analysis population for confirmation of the findings from the evaluable analysis population. Dots and error bars show GMR estimates and two-sided 95% CI respectively. GMR, geometric mean ratio; CI, confidence interval; ID, intradermal injection; IM, intramuscular injection; S, spike protein; RBD, receptor-binding domain; sVNT, surrogate virus neutralisation test; PRNT, plaque reduction neutralisation titres; FcγRIIIa, Fcγ receptor IIIa.

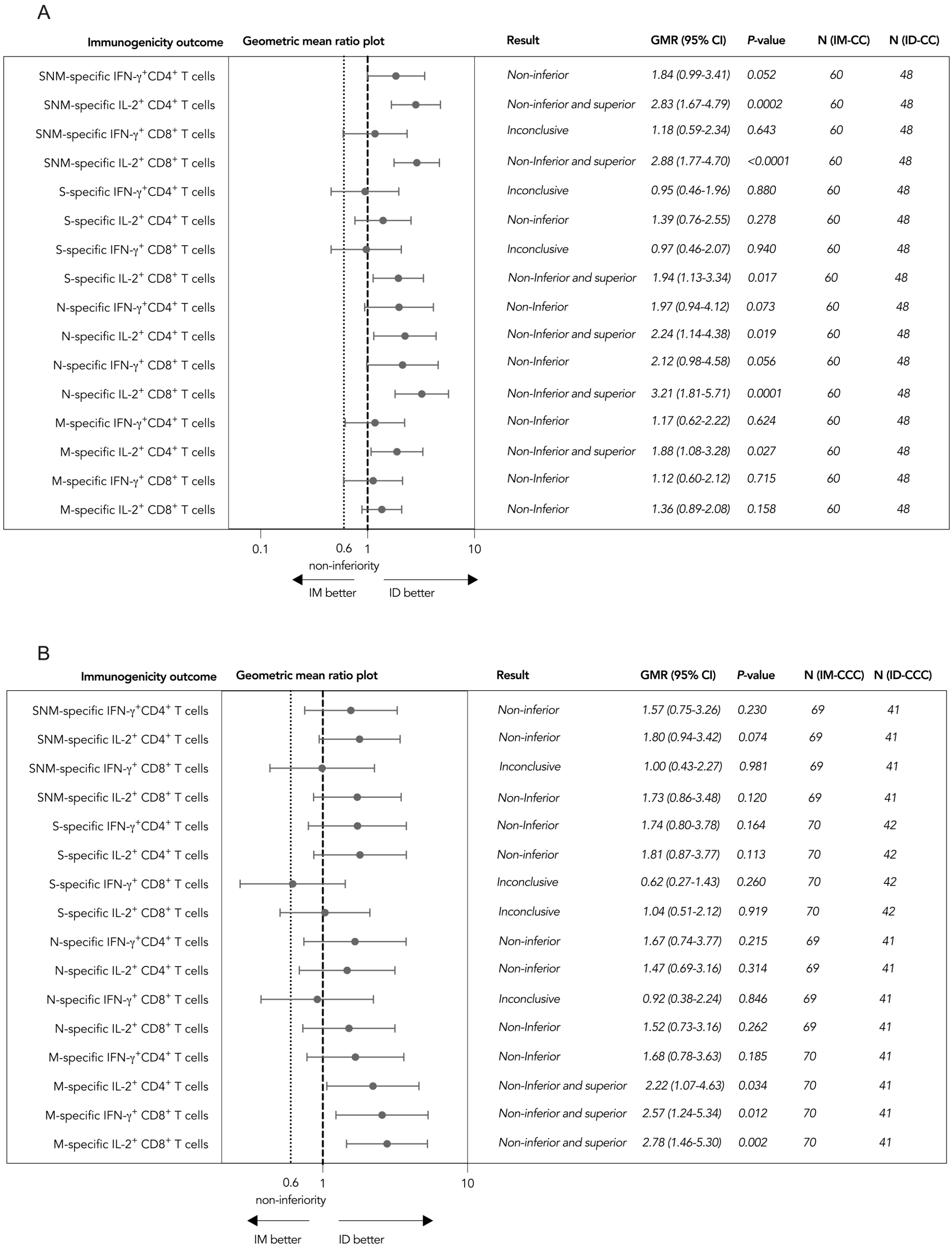


**Supp. Fig. 3. Superiority and non-inferiority hypotheses testing of cellular immunogenicity against wild type SARS-CoV-2 post-dose 2 and post-dose 3 of vaccination in the expanded analysis population.** Adolescents receiving (A) 2 doses of

CoronaVac administered intramuscularly or intradermally and (B) 3 doses of CoronaVac administered intramuscularly or intradermally were tested for T cell responses by flow-cytometry-based intracellular cytokine staining assays specific to S, N and M post-dose 2 or post-dose 3 in the expanded analysis population for confirmation of the findings from the evaluable analysis population. The results of SNM-specific T cell responses were calculated from the sum of responses of the individual S, N and M peptide pools. Dots and error bars show GMR estimates and two-sided 95% CI respectively. GMR, geometric mean ratio; CI, confidence interval; ID, intradermal injection; IM, intramuscular injection; S, spike protein; N, nucleocapsid protein; M, membrane protein; IFN-γ, interferon-γ; IL-2, interleukin-2.

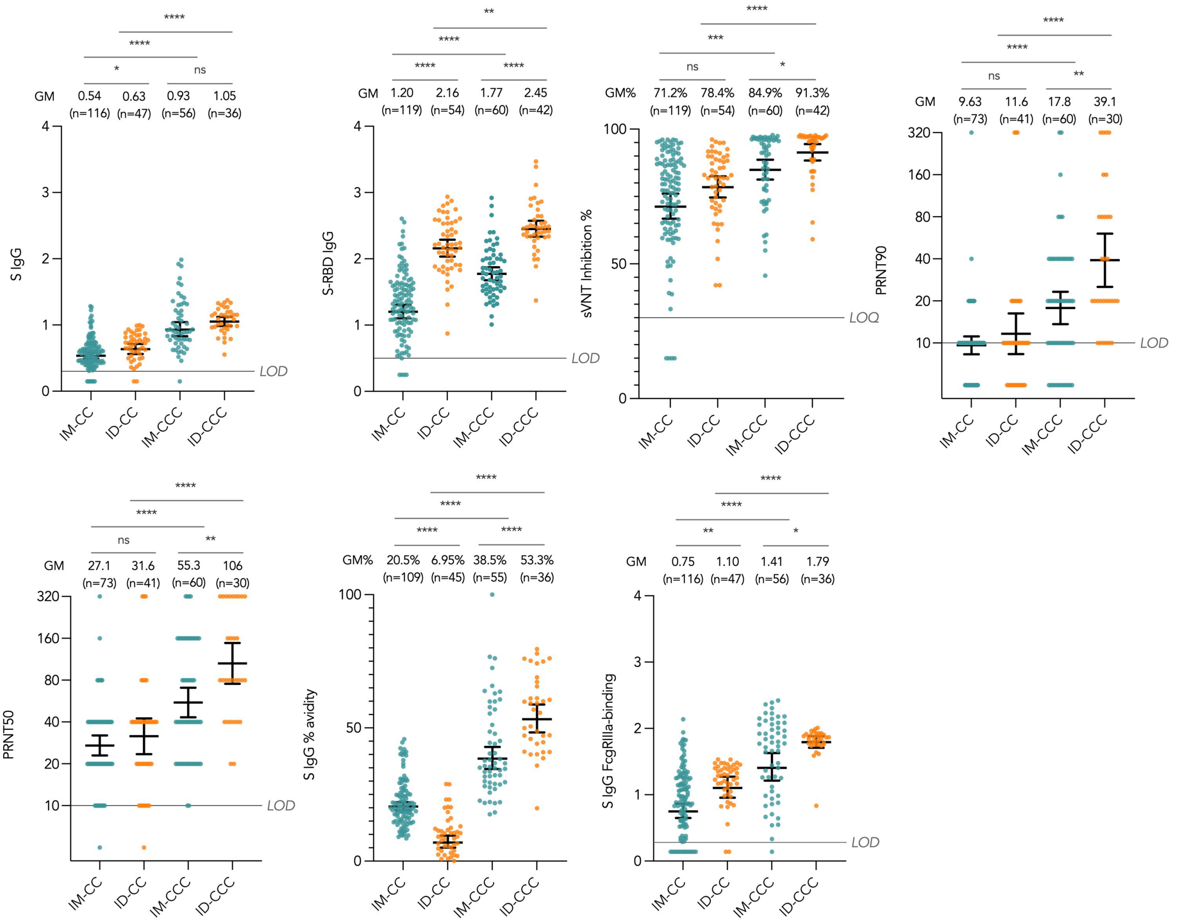


**Supp. Fig. 4. Changes in antibody responses post-dose 2 and post-dose 3 of vaccination in the evaluable analysis population.** Humoral immunogenicity outcomes were compared between CoronaVac administered intramuscularly (IM) or intradermally (ID) post-dose 2 and post-dose 3. Data labels and centre lines show GM estimates, and error bars show 95% CI. *P*-values were derived from two-tailed unpaired *t* test after natural logarithmic transformation. GM geometric mean, CI confidence interval; IM-CC and IM-CCC, 2 and 3 doses of vaccine administered intramuscularly, respectively; ID-CC and ID-CCC, 2 and 3 doses of vaccine administered intradermally, respectively; S, spike protein; RBD, receptor-binding domain; sVNT, surrogate virus neutralisation test; PRNT, plaque reduction neutralisation titres; FcγRIIIa, Fcγ receptor IIIa. *, $P<0.05$; **, $P<0.01$; ***, $P<0.001$; ****, $P<0.0001$; ns, no significant difference.

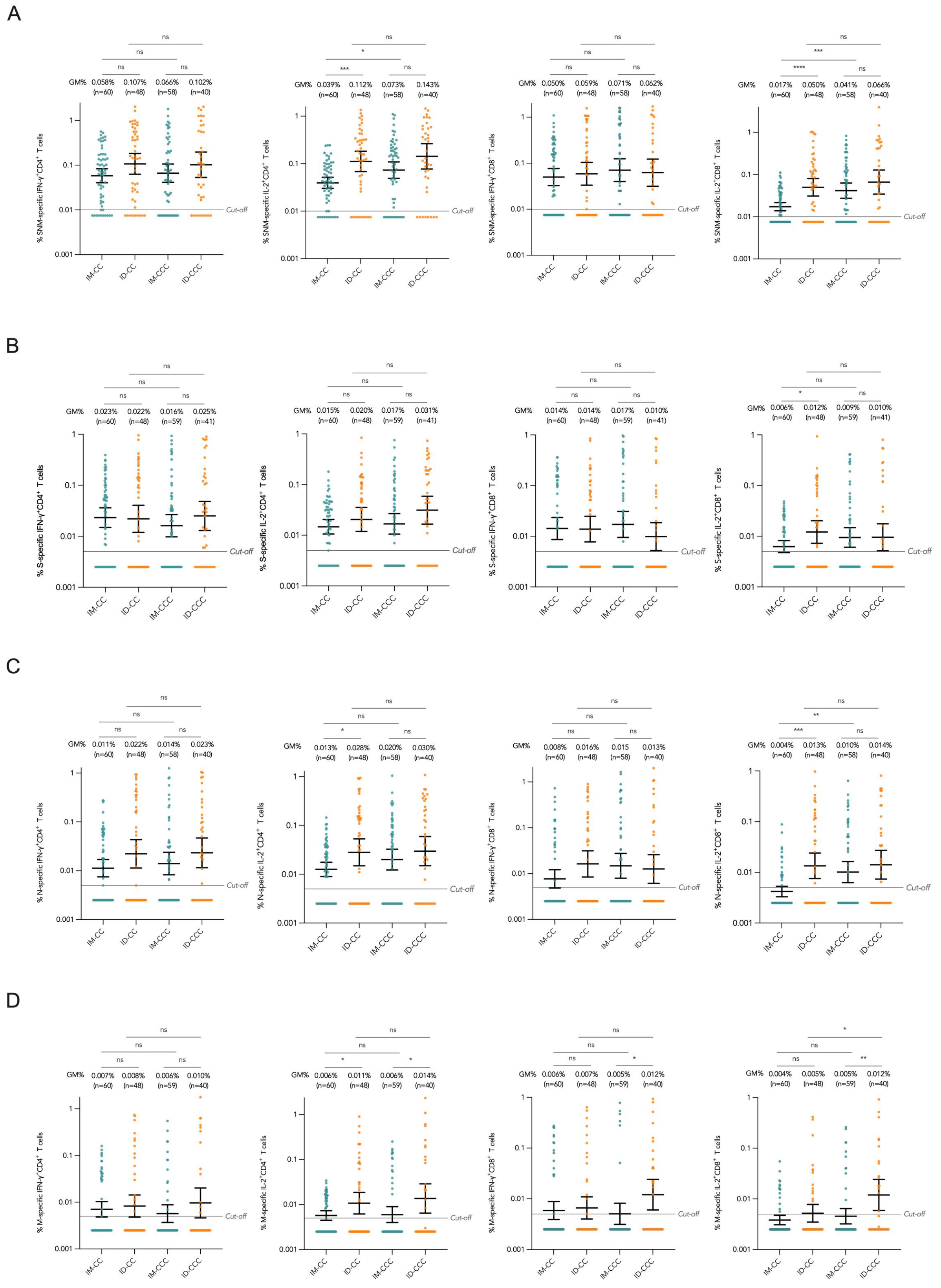


**Supp. Fig. 5. Changes in T cell responses post-dose 2 and post-dose 3 of vaccination in the evaluable analysis population.** Cellular immunogenicity outcomes were compared between CoronaVac administered intramuscularly (IM) or intradermally (ID) post-dose 2 and post-dose 3. Data labels and centre lines show GM estimates, and error bars show 95% CI. *P*-values were derived from two-tailed unpaired *t* test after natural logarithmic transformation. GM geometric mean, CI confidence interval; IM-CC and IM-CCC, 2 and 3 doses of vaccine administered intramuscularly, respectively; ID-CC and ID-CCC, 2 and 3 doses of vaccine administered intradermally, respectively; S, spike protein; N, nucleocapsid protein; M, membrane protein; IFN-γ, interferon-γ; IL-2, interleukin-2. *, $P<0.05$; **, $P<0.01$; ***, $P<0.001$; ****, $P<0.0001$; ns, no significant difference.

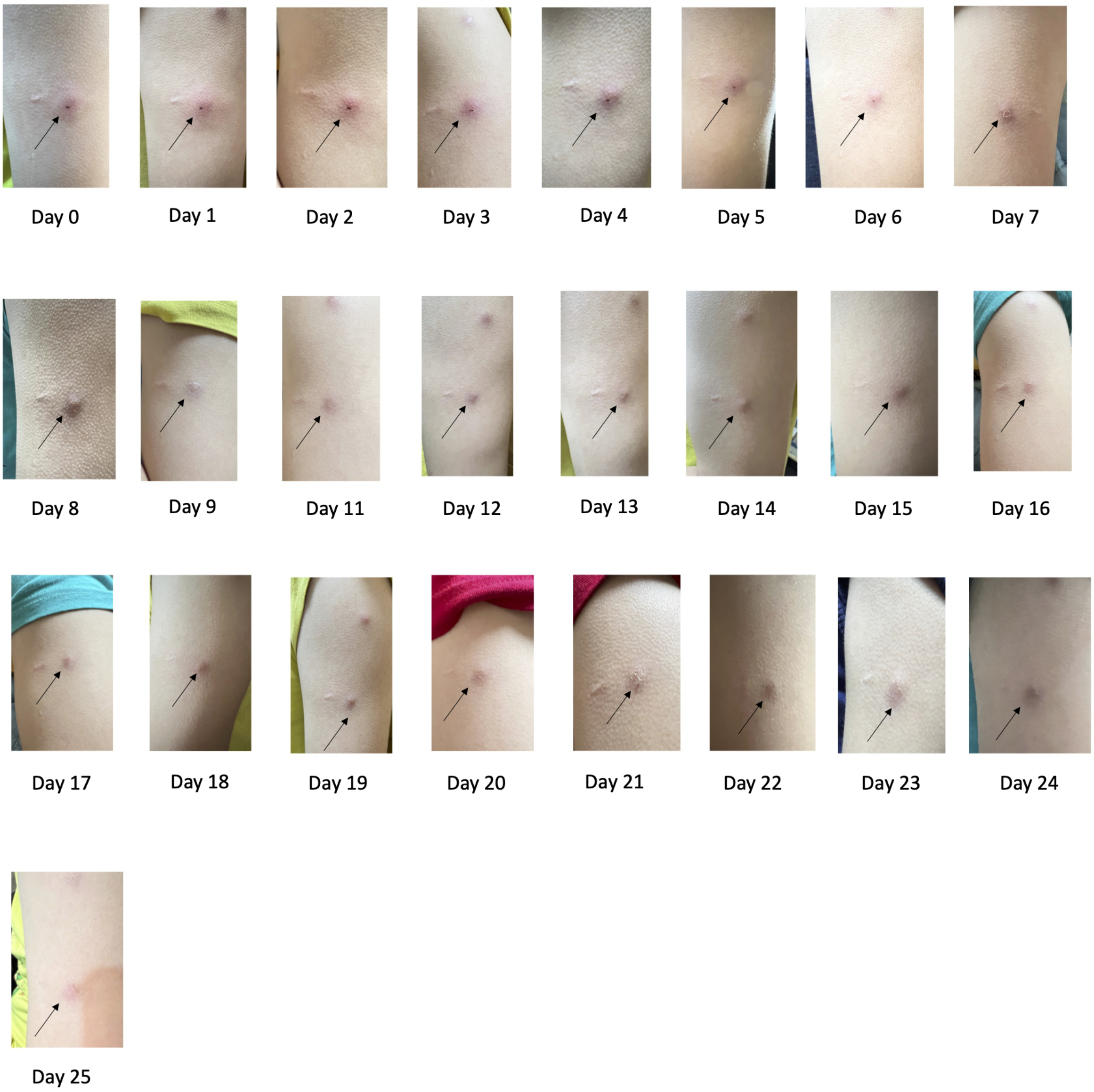


**Supp. Fig. 6. The evolution of cutaneous manifestations at the site of inoculation after dose 2 of intradermal CoronaVac.** The photos from an adolescent participant are representative of the typical injection site manifestations days 0 to 25 after dose 2 of CoronaVac administered intradermally.
