## Supplementary material for "Superior antibody and membrane protein-specific T cell responses to CoronaVac by intradermal versus intramuscular routes in adolescents": Protocol

### Study protocol

**Protocol title: To compare the reactogenicity and immunogenicity of the recommended COVID-19 vaccines in young adolescents and children in Hong Kong**

Protocol Identifier COVAC01

Version No. 7.2

Date July 11, 2022


Professor Yu Lung LAU

1/F, New Clinical Building, Queen Mary Hospital, Pokfulam, HK.


COVID-19 Vaccination in Adolescents and Children
兒童及青少年新冠病毒疫苗研究

HKU Med LKS Faculty of Medicine Department of Paediatrics & Adolescent Medicine
香港大學兒童及青少年科學系

C2i Centre for Immunology & Infection
免疫與感染研究中心

### Table of Contents


***Administrative information*** ***4***

**Title** **4**

**Trial registration** **4**

**Protocol version** **4**

**Funding** **9**

**Roles and responsibilities** **9**

***Declaration by principal investigator*** ***12***

***Summary*** ***13***

**Synopsis** **13**

**Schematic** **14**

**Schedule** **16**

***Introduction*** ***18***

**Background and rationale** **18**

**Objectives** **25**

**Trial design** **27**

***Methods: Participants, interventions, and outcomes*** ***28***

**Study setting** **28**

**Eligibility criteria** **28**

**Interventions** **32**

**Outcomes** **40**

**Participant timeline** **42**

**Sample size** **43**

**Recruitment** **44**

***Methods: Assignment of interventions (for controlled trials)*** ***45***

**Allocation: Sequence generation** **45**

**Allocation concealment mechanism** **45**

**Implementation** **45**

**Blinding (masking)** **45**

***Methods: Data collection, management, and analysis*** ***46***

**Data collection methods** **46**

**Data management** **57**

**Statistical methods** **57**

***Methods: Monitoring*** ***58***

**Data monitoring** **58**
**Harms** **59**
**Auditing** **59**
***Ethics and dissemination*** ***61***
**Research ethics approval** **61**
**Protocol amendments** **61**
**Consent or assent** **62**
**Confidentiality** **62**
**Declaration of interests** **62**
**Access to data** **63**
**Ancillary and post-trial care** **63**
**Dissemination policy** **63**

### Administrative information

#### Title

*1 Descriptive title identifying the study design, population, interventions, and, if applicable, trial acronym*

##### Complete Title

To compare the reactogenicity and immunogenicity of the recommended COVID-19 Vaccines in young Adolescents and Children in Hong Kong (COVAC)

##### Short Title

COVID-19 Vaccination in Adolescents and Children

##### Trial acronym

COVAC

#### Trial registration

*2a Trial identifier and registry name. If not yet registered, name of intended registry*

##### Trial registry

clinicaltrials.gov

##### Trial identifier

NCT04800133

*2b All items from the World Health Organization Trial Registration Data Set*

Are included in this protocol.

#### Protocol version

*3 Date and version identifier*

##### Protocol and version identifier

COVAC01

Version 7.2 (July 11, 2022)

#### Version history

This is the latest revised version (version 7.2) as of July 11 2022. Summary of key changes including rationale are appended here.

New changes in version 7.2 dated July 11 2022

| Section | Change | Rationale |
|---|---|---|
| Outcomes | Added IgG allotypes | To explore correlates of vaccine responses |
| Data storage | Added transferring of pseudonymized samples to overseas collaborators | For additional immune testing not available in Hong Kong |

New changes in version 7.1 dated May 6 2022

| Section | Change | Rationale |
|---|---|---|
| Intervention | CS-B: dose 2 offered as booster, optional | Updated vaccine regimen due to Omicron wave in Hong Kong |

New changes in version 7.0 dated March 1 2022

| Section | Change | Rationale |
|---|---|---|
| Intervention | CA-B: dose 3 offered, optional | Updated vaccine regimen due to Omicron wave in Hong Kong |
| Intervention | CA-C/I: dose 3 offered, optional | Updated with preliminary data |
| Intervention | CE: dose 4 offered, optional | Updated vaccine regimen due to Omicron wave in Hong Kong |
| Eligibility | CE-C: age limit lower to 0 months | High risk due to very young age and immunocompromise during the Omicron wave; no safety concerns observed in healthy young children data from phase 2 trial |

New changes in version 6.1 dated Feb 1 2022

| Section | Change | Rationale |
|---|---|---|
| Study setting | Sun Yat Sen Memorial Park Sports Centre Community Vaccination Centre added | New study site |

New changes in version 6.0 dated Jan 10 2022

| Section | Change | Rationale |
|---|---|---|
| Eligibility criteria/ intervention | Arm CE-B/p added (3 doses of BNT162b2 at one-third of adult dosage for paediatric patients aged 5-11) | To study immune responses in younger children with immune compromise/ paediatric illnesses |

New changes in version 5.0 dated Oct 4 2021

| Section | Change | Rationale |
|---|---|---|
| Schematic, schedule | Timing for post-third dose visit is updated | To allow flexibility for third dose timing |
| Eligibility criteria/ intervention | Age limit lowered to 3 years for CoronaVac arms | To study immune responses in younger children |
| Intervention | Arms CA-C/I and CE-C/I added (intradermal injection of 2 doses of CoronaVac) | To compare intradermal and intramuscular injection |

New changes in version 4.0 dated July 7 2021

| Section | Change | Rationale |
|---|---|---|
| Schematic, schedule | Timing for third dose from 6 months earliest to 2 months post dose 1 earliest | To allow flexibility for third dose timing |
| Objectives | To compare children and parents and unrelated adults instead of children and parents only | Parents not always available |
| Eligibility criteria | Allow unrelated adults to be recruited | Parents not always available |
| Interventions | Arm CA-C/3 and CE-C/3: timing of third dose from day 161-175 to day 56 or after | To allow flexibility for third dose timing |
| Outcomes | Add 2 weeks post-dose 3 as timepoint | To assess response to dose 3 |

| | | |
|---|---|---|
| Definition of analysis population | Deleted sections on modified intention to treat analysis and missing data | To refer to statistical analysis plan at interim analysis |

New changes in version 3.0 dated May 20 2021

| Section | Change | Rationale |
|---|---|---|
| Funding | Added private donations and funders have no role in design, execution and release of this study | Inclusion of private donations for additional tests |
| Summary | Deleted study arm A (AZD1222 by Oxford/AstraZeneca) | No AZD1222 will be provided in Hong Kong |
| Summary, Schematic, Introduction, Objectives | Added recruitment of adolescents and parents who have been previously diagnosed with COVID-19 and patients with immune, blood and severe paediatric illnesses in our study | To include such special patient groups |
| Schedule | Table summarizing events taking place after optional dose 3 | Optional dose 3 |
| Schedule | Revised study windows | Flexibility |
| Introduction | Added preliminary results from BioNTech and SinoVac; added information related to heparin-induced-like thrombocytopenic thrombosis for AZD1222 | To update with new data |
| Objectives | Specified non-inferiority criterion | Clarified non-inferiority framework |
| Methods | Revised age from 11-16 years to 11 years or above | Revise eligibility as CoronaVac only available to those aged 18 years or above in Hong Kong |
| Methods | Revised eligibility to include parents or relatives of students | Revise eligibility as parents not always available |
| Methods | Added criteria for patients with previous COVID-19, patients with severe paediatric/immune conditions | To include these special patient groups |
| Interventions | Deleted arms CA-A and CE-A | Not provide AZD1222 |
| Interventions | Added CA-C/3 and CE-C/3, three-dose | Offer third dose of SinoVac CoronaVac |
| Interventions | Added Arm CS-B/1, to give one dose of BNT162b2 to those previously infected | To recruit these patients |

| | | |
|---|---|---|
| Interventions | Added Arms CS-C/1 and /2, to give one or two doses of SinoVac CoronaVac to those previously infected | To recruit these patients |
| Interventions | Added intervention modification for participants upon request | To allow modification |
| Outcomes | Added percentage of occurrence, types, duration and severity of long COVID-19 abnormalities as an outcome | To capture long COVID changes with vaccination |
| Sample size | For subgroups CE and CS | To justify sample size |
| Pre-existing history assessment | For subgroups CE and CS | Baseline data for CE and CS |
| Statistical methods | For subgroups CE and CS | To describe statistical management for CE and CS |

New changes in version 2.0 dated Apr 19, 2021

| Page and section | Change | Rationale |
|---|---|---|
| Cover page | Revised study logo | To include HKU Centre for Immunology and Infection, the affiliation of CoA Prof Malik Peiris |
| Trial registration | Trial identifier from ‘pending’ to ‘NCT04800133’ | Successful trial registration on clinicaltrials.gov |
| Trial registration | Updated trial identifier and IRB number | Successful trial registration and ethics approval |
| Eligibility | Added ‘receipt of coronavirus vaccines in the past’ as exclusion criteria | To exclude any participants already vaccinated |
| Interventions Arm C | Changed unit from 3ug/0.5ml to 600SU/ml | To align with product insert as requested by Department of Health |
| Blood-based assessment | Added 1 EDTA tube for each participant; and blood drawn amount becomes uniform for two parents | To allow complete blood count; to allow full immune tests for both parents |
| Statistical methods | Added description of statistical analyses on secondary outcomes | To describe statistical analyses on secondary outcomes |
| Informed Consent | Updated information sheet | As requested by Department of Health and to reflect protocol updates |

| | | |
|---|---|---|
| Biological Specimens | Added 1 EDTA tube for each participant; and blood drawn amount becomes uniform for two parents | To allow complete blood count; to allow full immune tests for both parents |

#### Funding

##### *4 Sources and types of financial, material, and other support*

This study is funded by the Food and Health Bureau, Hong Kong Special Administrative Region and private donations. Funders have no role in the design, execution and release of this study.

#### Roles and responsibilities

##### *5a Names, affiliations, and roles of protocol contributors*

**Coordinating author**

Yu Lung Lau, principal investigator

**Authors**

Malik Peiris, investigator

Wenwei Tu, investigator

Wing Hang Leung, investigator

Patrick Ip, Investigator

Pamela Pui-Wah Lee, Investigator

Jaime Rosa Duque, Investigator

Gilbert T Chua, Investigator

Winnie Wan-Yee Tso, Investigator

Kai N. Cheong, Investigator

Wilfred Hing Sang Wong, Investigator

Daniel Leung, Investigator

##### *5b Name and contact information for the trial sponsor*

**Sponsor**

The University of Hong Kong

#### Principal investigator

Professor Yu Lung Lau

Office: Room 106, 1/F, New Clinical Building, 102 Pokfulam Road, Queen Mary Hospital, HK.

##### *5c Role of study sponsor and funders, if any, in study design; collection, management, analysis, and interpretation of data; writing of the report; and the decision to submit the report for publication, including whether they will have ultimate authority over any of these activities*

The study sponsor(s) and the funder(s) have no role in the design, performance, interpretation, reporting or any other parts of the study, nor any authority over these activities. The principal investigator Yu Lung Lau conducts and oversees all aspects of the study.

##### *5d Composition, roles, and responsibilities of the steering committee, endpoint adjudication committee, data management team, and other individuals or groups overseeing the trial, if applicable (see Item 21a for data monitoring committee)*

#### Steering and endpoint adjudication committee

The steering committee is responsible for the oversight of the study. It also serves as the endpoint adjudication committee, responsible for examination and approval of outcome measures as requested by the trial staff. Members include:

Yu Lung Lau, Principal Investigator

Malik Peiris, Investigator

Wenwei Tu, Investigator

Wing Hang Leung, Investigator

Patrick Ip, Investigator

Pamela Pui-Wah Lee, Investigator

Jaime Rosa Duque, Investigator

Gilbert T Chua, Investigator

Winnie Wan-Yee Tso, Investigator

Kai N. Cheong, Investigator

Wilfred Hing Sang Wong, Investigator

Daniel Leung, Investigator

#### Data management team

The data management team is responsible for data management, including ensuring data confidentiality and transparency in the performance of this study. Members include:

Wilfred Wong, Senior IT Manager

Sau Man Chan, Research Nurse in Charge

### Declaration by principal investigator

I have read and understood the latest version of the protocol for the study dated July 11 2022, and agree to supervise all aspects of the trial and ensure its proper conduct according to this current protocol, the International Council for Harmonisation Good Clinical Practice guidance, and all applicable laws and institutional regulations. I agree to implement changes to the protocol only with institutional ethics approval or to mitigate an unforeseen imminent risk to participants.

digitally signed

____________________________________

Yu Lung Lau, Principal Investigator

Dated July 11 2022

### Summary

#### Synopsis

There are now 160 million cases of COVID-19 since the onset of the pandemic with over 3 million deaths globally. For children, prolonged school closure and isolation have generated more mental health deterioration and self-reported adverse experiences. Moreover, it is now understood that children and adolescents under 20 years of age are more likely to infect others than adults older than 60 years. Therefore, establishment and maintenance of immunity in children and adolescents are key steps that will be necessary to decrease household transmission and protect older persons.

This study plans to generate timely data on reactogenicity and immunogenicity of COVID-19 candidates in Hong Kong adolescents and children, as data from paediatric trials are not yet expected to be available until the commencement of the next school year. The primary objectives of this study include:

1. Describe the frequencies of reactogenicity within the 7 days after vaccine injection;

2. Compare such frequencies among the vaccines.

3. Short-term and long-term (up to 36 months post-vaccination) immune responses including spike protein IgG and related markers, neutralizing antibody titres and B and T cell immune responses

4. Differences between children and adults, such that the geometric mean values of immunological parameters are statistically non-inferior (0.6 of adult value), and differences between vaccines

COVID-19 vaccines are planned to be purchased or have begun being distributed to the public in Hong Kong, and will be the study vaccines in our nonrandomized comparator trial. Their efficacy has been studied in phase 3 clinical trials, and data suggest they offer more than 50% protection against COVID-19 in adults.

1. Study Arm B: BNT162b2 by BioNTech/Fosun
2. Study Arm C: CoronaVac by SinoVac

Participants aged 3-17 years and adult controls will be recruited for this study, which takes place over 37 months. As shown in the schematic, 6 basic visits over 37 months will be scheduled. CoronaVac may be given intramuscularly or intradermally, as opted for by participants, and their immunogenicity will be compared. In addition to healthy adolescents and children, adolescents (and their parents) who have been previously diagnosed with COVID-19 will be recruited, and given 1 or 2 shots as it is known that reinfection, albeit uncommon, may occur in COVID survivors. Paediatric patients with immunological, rheumatological and blood disorders, as well as other severe paediatric illnesses will also be included in our trial as they may be at increased risk of severe COVID and poor immune control of virus allows evolution and possible emergence of new variants. Patients with acute lymphoblastic leukemia who developed an allergy to peg-asparaginase will receive their dose 1 of BNT162b2 in a gold-standard graded challenge or without modification as BNT162b2 contains polyethylene glycol (PEG).

Vaccination will be given in the first 2-3 visits for most participants, scheduled according to the latest scientific recommendation, and blood taking at all visits. Nasopharyngeal swab will be collected at visits 1, 3 and 4 for most participants, but refusal of the swab is not an exclusion criterion. Adverse reactions will be solicited for 7 days after each dose. Adverse events will be monitored 28 days after each dose, and severe adverse events, Covid-19 and pregnancy will be monitored throughout the 37 months of the study period from each participant via an electronic diary card.

Results will be reported as soon as possible to the government, the public and the academic community via release of preprints and articles in academic journals. This trial is approved by the Institutional Review Board of The University of Hong Kong/Hospital Authority Hong Kong West Cluster (UW21-157), and the Department of Health, Hong Kong Special Administrative Region Government (Clinical Trial Certificate 101894). This trial is registered at clinicaltrials.gov (NCT04800133).

#### Schematic

##### Healthy adolescents and children

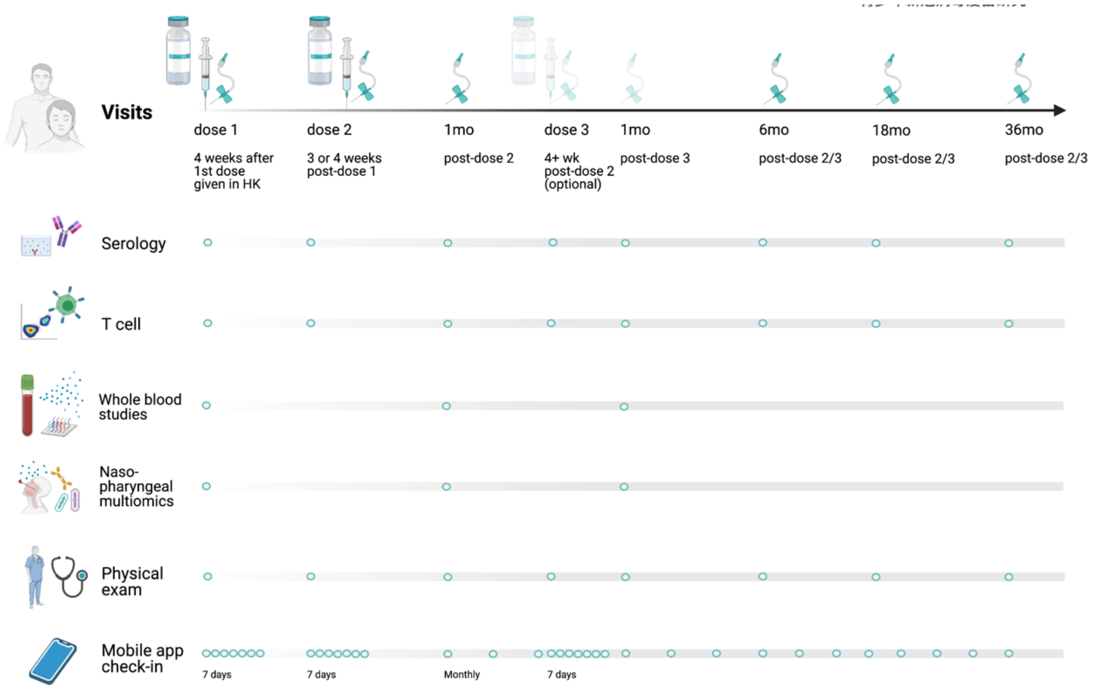

#### COVID survivors

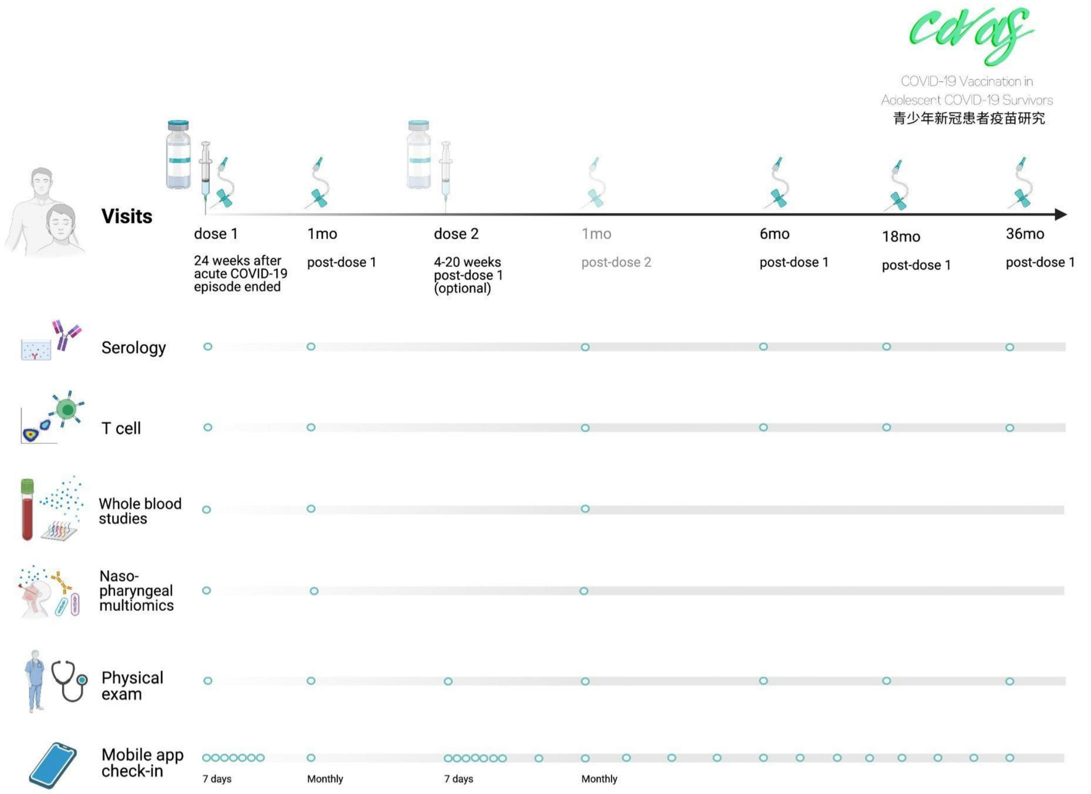


#### Patients with immunological, rheumatological and hematological diseases

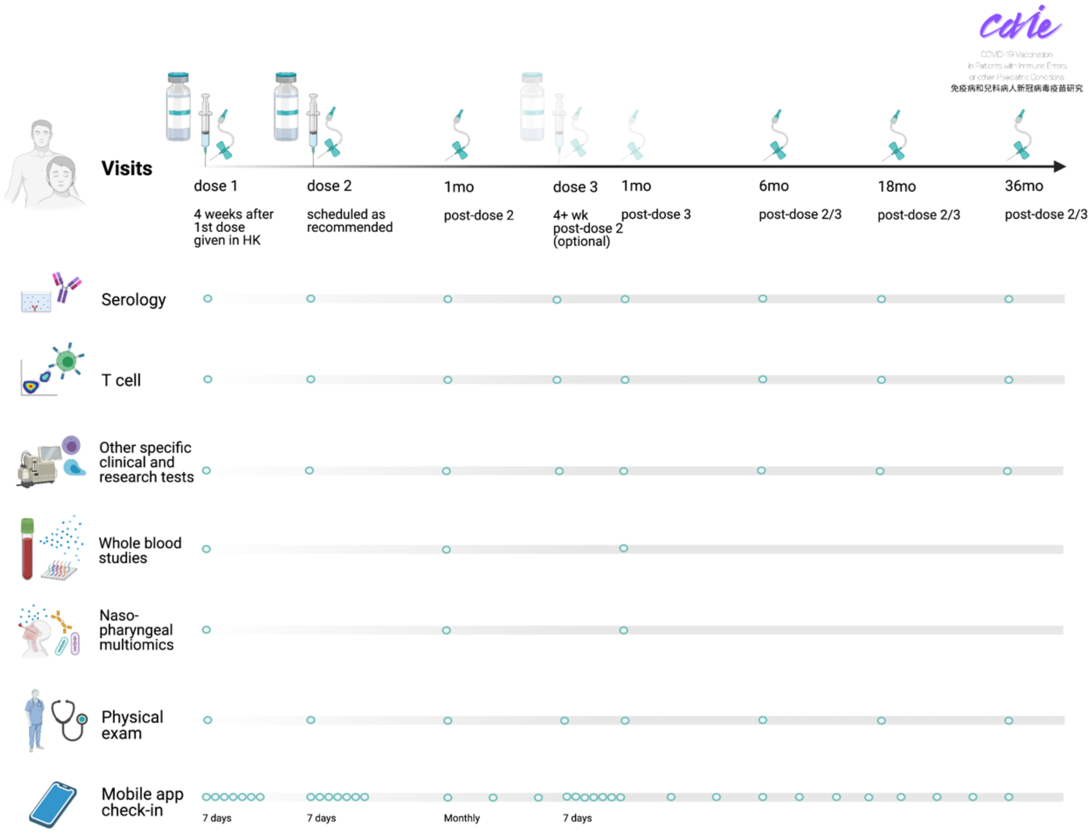

### Schedule

| | Before dose 1 | **Visit 1**<br>Dose 1 (day 0) | 7 days post dose 1<br>(day 1-6) | 21-28 days post dose 1<br>(day 1-20/27) |
|---|---|---|---|---|
| Participant registration and eligibility screen | X | | | |
| Eligibility, history, vital signs, physical exam | | X | | |
| Bloodtaking | | X | | |
| Nasopharyngeal swab* | | X * | | |
| Vaccination | | X | | |
| Solicitation of AR | | X | X | |
| Solicitation of AE | | X | X | X |
| Solicitation of SAE, SE | | X | X | X |

| | **Visit 2**<br>Dose 2 (day 21/28$) | 7 days post dose 2<br>(day 22-27/29-34) | 28 days post dose 2<br>(day 22-48/29-55) | **Visit 3**<br>1 mo+ post dose 2# |
|---|---|---|---|---|
| Eligibility, history, vital signs, physical exam | X | | | X (If any AE reported) |
| Bloodtaking | X | | | X |
| Nasopharyngeal swab* | | | | X * |
| Vaccination | X | | | |
| Solicitation of AR | X | X | | |
| Solicitation of AE | X | X | X | |
| Solicitation of SAE, SE | X | X | X | X |

| | Dose 3<br>(day 49+@) | 6 days post dose 3<br>(day 50+) | 14 days post dose 3<br>(day 63+) | 27 days post dose 3<br>(day 50-) |
|---|---|---|---|---|
| Eligibility, history, vital signs, physical exam | X | | | |
| Bloodtaking | X | | X | |
| Nasopharyngeal swab* | | | X | |
| Vaccination | X | | | |
| Solicitation of AR | X | X | | |
| Solicitation of AE | X | X | X | X |
| Solicitation of SAE, SE | X | X | X | X |

| | Dose 4<br>(day 133+@) | 6 days post dose 4<br>(day 134+) | 14 days post dose 4<br>(day 147+) | 27 days post dose 4<br>(day 134-) |
|---|---|---|---|---|

| | | | | |
|---|---|---|---|---|
| Eligibility, history, vital signs, physical exam | X | | | |
| Bloodtaking | X | | X | |
| Nasopharyngeal swab* | | | X | |
| Vaccination | X | | | |
| Solicitation of AR | X | X | | |
| Solicitation of AE | X | X | X | X |
| Solicitation of SAE, SE | X | X | X | X |

| | **Visit 4** 6 mo post dose 2/3/4^ | **Visit 5** 18 mo post dose 2/3/4^ | **Visit 6** 36 mo post dose 2/3/4^ | After 36 mo post dose 2/3/4 |
|---|---|---|---|---|
| Eligibility, history, vital signs, physical exam | X (If any AE reported) | X (If any AE reported) | X (If any AE reported) | |
| Bloodtaking | X | X | X | |
| Nasopharyngeal swab* | X * | | | |
| Vaccination | | | | |
| Solicitation of AR | | | | |
| Solicitation of AE | | | | |
| Solicitation of SAE, SE | X | X | X | |

Abbreviations: AR adverse reactions, AE adverse events, SAE severe adverse events, SE special events. Note: * Nasopharyngeal swab is opt-in for participants. $ Dose 2 is preferably given fewer than 7 days later but not before. Visit 2 bloods is preferably drawn up to 7 days before. # Visit 3 can be scheduled preferably not more than 14 days earlier or 14 days later. @ An optional dose 3 of CoronaVac or BNT162b2 for some patients is scheduled 4 or more weeks after the 2$^{nd}$ dose. Optional dose 4 is scheduled 12 or more weeks after the 3$^{rd}$ dose. ^ Visits 4-6 may be scheduled 42 days earlier or later

### Introduction

#### Background and rationale

*6a Description of research question and justification for undertaking the trial, including summary of relevant studies (published and unpublished) examining benefits and harms for each intervention*

##### General background

There are now 160 million cases of COVID-19 since the onset of the pandemic with over 3 million deaths globally (1). Hong Kong (HK) has had over 10,000 cases, with an overall case fatality rate of 1.9% (2). Most deaths occurred in those over 60 years old, while those younger than 20 years old have had mostly mild disease or were asymptomatic (3). In addition to the physical health impact as evident by the severe morbidity and mortality, there has been enormous disruption of everyone's livelihoods due to non-pharmaceutical interventions, such as social distancing policies and practices (4). For children, prolonged school closure and isolation have generated more mental health deterioration and self-reported adverse experiences, while the precise ensuing long-term consequences has yet to be determined (5). There have also been delays in patients accessing health care, resulting in late diagnoses of children cancers and other serious diseases, such as disseminated tuberculosis in early infancy (personal observation, YL Lau). Moreover, it is now understood that children and adolescents under 20 years of age are more likely to infect others than adults older than 60 years, with an odds ratio of 1.58 according to one study (6). Therefore, establishment and maintenance of immunity in children and adolescents are key steps that will be necessary to decrease household transmission and protect older persons (7). Accumulating release of data from ongoing phase 3 trials and national programs supports this notion that vaccine efficacy is important for blocking transmission, albeit likely this phenomenon is less effective compared to symptomatic diseases. Based on these observations, whole population vaccination is now the standard aim for most countries and regions that have the infrastructure to deliver this goal, such as Israel. This intervention is expected to aid our society's transition from the pandemic to endemic phase as soon as possible, so that our community can regain near-normal social and economic activities (8). With this goal in mind, Israel has already delivered 3.08 million first vaccine doses and 1.79 million second vaccine doses for its 9 million population by the end of January 2021, including children between 10-19 years old. The United Kingdom has vaccinated over 15 million of the 68 total million population after 10 weeks of the national vaccination program as of mid-February 2021.

In HK, commencement of a COVID-19 vaccination programme is expected to begin in March 2021 (9). The initial phase would target members of the community who are at the highest risk of contracting COVID-19 or death, such as healthcare workers, the elderly and those with chronic medical conditions. At the time of writing, the HK Government has announced the planned procurement and distribution of 3 COVID-19 vaccines (10):

- The BioNTech vaccine BNT162b2 (Comirnaty) consists of a small segment of messenger RNA (mRNA) encoding the spike protein from the SARS-CoV2 virus (11). After administration, this nucleic acid material translates into SARS-CoV2 spike protein antigen that ultimately leads to development of immunity (11). Although this is based on a novel technological concept, the safety profile from clinical trials and post-marketing surveillance appears to be satisfactory, with rare allergic events postulated to be due to the pegylated portion of the lipid nanoparticle carrier and adjuvant (11). The immunization regimen requires 2 injections, separated by 21 days. Of all the vaccines, the BNT162b2 has reported the highest efficacy rate at preventing symptomatic infections, although there has been no study design with a direct head-to-head comparison to other vaccines. Moreover, the vaccine efficacy to prevent asymptomatic infections is not known.

- Sinovac Biotech's vaccine CoronaVac has employed the more traditional vaccinology methodology by using the inactivated whole SARS-CoV-2 virus as the antigen (12). The immunization regimen requires 2 injections, with 14 days in between for emergency use but 28 days for routine use which could generate higher immunogenicity. This vaccine appears to be promising based on results from phase 1 and 2 trials, and a phase 3 study in Brazil reported efficacy of more than 50% in preventing infection from very mild to severe cases. The safety profile seems more favourable than BNT162b2.
- AZD1222 (Vaxevria) by Oxford-AstraZeneca is a viral vector vaccine using an engineered chimpanzee adenovirus and a phase 3 study reported efficacy of between 62-90% in preventing any infection. (13). Interestingly, lower initial dosing was associated with a more robust response. Due to possible confounding effects, more investigation regarding this opposite correlation is needed. Two injections are given 28 days apart, although trials comparing this schedule with a single administration, delayed second dose and heterologous vaccination with other vaccines are currently under exploration.

Regardless of the vaccine used, all COVID-19 vaccines approved for emergency use as of now have no published data for people younger than 16 years old before the start of this trial. Therefore, paediatric studies are being advocated and pursued actively. As of early February 2021, the BNT162b2 phase III trial has recruited 2,000 children aged 12-16 years, and preliminary data will be submitted in March 2021 to various regulatory bodies (personal communication, Eleni Lagkadinou, BioNTech, Germany), preliminary results of which suggests favorable safety and up to 100% efficacy. Dose reduction to ⅓ of adult dose was trialled for children aged 5-11 years for BNT162b2 and press release from Pfizer/BioNTech showed comparable immunogenicity with adolescents. As mRNA vaccines are associated with myocarditis, especially in adolescents, the use of 2-dose BNT162b2 is not clearly known to cause more benefit than risk in paediatric populations. Oxford-AstraZeneca was planning to test AZD1222 in 240 British children aged 6 and above beginning late February 2021 (14), yet the trial was suspended due to concerns of heparin-induced-like thrombocytopenic thrombosis in young adults. Sinovac Biotech has tested CoronaVac in children aged 3 to 17 years old with results to be submitted to regulatory bodies in May 2021(15, 16), and results suggest the full dose in adolescents aged 12 or above had similar but slightly higher immunogenicity than in adults.

As paediatric data for these vaccines may not be available in time from companies for the HK Government to decide on immunization plans for children in order to achieve the goal of whole population vaccination within 12 months, with at least 70 to 80% of our population vaccinated for establishing herd immunity, we therefore propose to generate our own paediatric data for the 3 chosen vaccines at the beginning of our adult vaccination program (11-13). This project is powered to provide convincing data to our public that these vaccines are safe and immunogenic for children, which will pave the way for whole population vaccination. Local academic research not funded by pharmaceutical companies on these vaccines may also reduce public perception of bias and hesitancy towards these new vaccines. This is important as evident by a recent local survey from the University of Hong Kong (HKU) that showed less than half of respondents intend to receive vaccines against COVID-19 (17). In children and adolescents, we will also trial intradermal regimen for CoronaVac to test the hypothesis that intradermal CoronaVac will result in superior immunogenicity than intramuscular route, as it has been shown for inactivated flu vaccines previously Moreover, our study will also provide novel immunogenicity and reactogenicity data on the BNT162b2 and AZD1222 vaccines focused on East Asian adults and children for the first time, as well as head-to-head comparison of these 3 COVID-19 vaccines which are based on entirely different technology platforms.

Apart from healthy adolescents and children, our study also aims to study the reactogenicity and immunogenicity of the vaccines in COVID-19 survivors, and patients with paediatric conditions. Cases of reinfection have occurred, the first case worldwide having been reported in Hong Kong (18). In general, prior COVID-19 is estimated to confer 84% protection against any subsequent infections (19).

Numerous studies have demonstrated that a single dose of mRNA vaccine in previously infected adults is adequate to raise antibodies and T cell response to a level approximate to two doses of mRNA vaccine, with the second dose potentially reducing antibody or cellular responses (20, 21). Moreover, there is no data for the number of doses of inactivated vaccine required for COVID-19 survivors. We aim to demonstrate that a single dose of mRNA vaccine, and possibly of inactivated vaccine, are also adequate for previously infected children. Moreover, as we understand many COVID-19 survivors endure long COVID, including local infected children with brain fog or psychiatric conditions, there is emerging scientific discussion and data that vaccination may reduce long COVID.

Patients with paediatric conditions, such as inborn errors of immunity, childhood blood cancers, paediatric lupus, and Down syndrome are at a much higher risk of severe COVID-19 or multisystem inflammatory syndrome than healthy counterparts (22-25). Inborn errors of type I interferon immunity and autoantibodies against type I interferon have been demonstrated to cause no less than 3% and 20% of severe COVID-19 respectively (26, 27 and unpublished data). Mutations in the SOCS1 gene, a key regulator of interferon responses, have also been implicated in multisystem inflammatory syndrome (28). It is now thought that genetic mutations and their autoimmune phenocopies account for most cases of severe COVID-19, and that common lifestyle diseases such as hypertension or diabetes have only a small effect on COVID-19 severity. The study of safety and immunogenicity of COVID-19 vaccines in patients with severe paediatric conditions are therefore extremely essential to protect those most vulnerable to COVID-19. Moreover, immunocompromised patients allow a higher magnitude and longer duration of viral replication, increasing the likelihood of generating new virus variants (29, 30). Effective immunization of these patients and its study would be essential to prevent the emergence of new variants of concern, which may have higher transmissibility or immune escape, bearing public health importance. Among these patients, there are likely to be non-responders known as primary vaccine failure, to whom we will offer a 3rd dose of the same vaccine via intramuscular or intradermal route, as intradermal route has been shown to elicit higher immunogenicity for other vaccines before including by our group (31).

A unique group of patients under our study would be patients with acute lymphoblastic leukemia (ALL), who may be treated with pegylated asparaginase with PEG of molecular weight 345-410 Daltons, and around 1 in 4 develop allergy to polyethylene glycol (PEG) (32, 33). As BNT162b2 also contains PEG with a molecular weight of 2000 Daltons, albeit of a different molecular weight, we seek to understand whether there is cross-reactivity, and how BNT162b2 could be given safely to ALL survivors. We will recruit patients with non-severe known PEG allergy, and perform a 2 or 3-step graded challenge with BNT162b2.


1. Dong E, Du H, Gardner L. An interactive web-based dashboard to track COVID-19 in real time. Lancet Infect Dis. 2020;20(5):533-4.
2. HKSARG Centre for Health Protection. Latest Situation of Coronavirus Disease (COVID-19) in Hong Kong.
3. Dorjee K, Kim H, Bonomo E, Dolma R. Prevalence and predictors of death and severe disease in patients hospitalized due to COVID-19: A comprehensive systematic review and meta-analysis of 77 studies and 38,000 patients. PLoS One. 2020;15(12):e0243191.
4. Zimet GD, Silverman RD, Fortenberry JD. Coronavirus Disease 2019 and Vaccination of Children and Adolescents: Prospects and Challenges. J Pediatr. 2020.
5. Panda PK, Gupta J, Chowdhury SR, Kumar R, Meena AK, Madaan P, et al. Psychological and behavioral impact of lockdown and quarantine measures for COVID-19 pandemic on children, adolescents and caregivers: a systematic review and meta-analysis. J Trop Pediatr. 2020.
6. Li F, Li YY, Liu MJ, Fang LQ, Dean NE, Wong GWK, et al. Household transmission of SARS-CoV-2 and risk factors for susceptibility and infectivity in Wuhan: a retrospective observational study. Lancet Infect Dis. 2021.
7. Klass P, Ratner AJ. Vaccinating Children against Covid-19 - The Lessons of Measles. N Engl J Med. 2021.
8. Chodick G, Tene L, Patalon T, Gazit S, Tov AB, Cohen D, et al. The effectiveness of the first dose of BNT162b2 vaccine in reducing SARS-CoV-2 infection 13-24 days after immunization: real-world evidence. medRxiv. 2021:2021.01.27.21250612.
9. COVID-19 vaccine to arrive soon [Available from: https://www.news.gov.hk/eng/2021/02/20210202/20210202_101618_818.html.

10. HKSARG. Government announces latest development of COVID-19 vaccine procurement [Available from: https://www.info.gov.hk/gia/general/202012/12/P2020121200031.htm.
11. Polack FP, Thomas SJ, Kitchin N, Absalon J, Gurtman A, Lockhart S, et al. Safety and Efficacy of the BNT162b2 mRNA Covid-19 Vaccine. N Engl J Med. 2020;383(27):2603-15.
12. Zhang Y, Zeng G, Pan H, Li C, Hu Y, Chu K, et al. Safety, tolerability, and immunogenicity of an inactivated SARS-CoV-2 vaccine in healthy adults aged 18-59 years: a randomised, double-blind, placebo-controlled, phase 1/2 clinical trial. Lancet Infect Dis. 2021;21(2):181-92.
13. Voysey M, Clemens SAC, Madhi SA, Weckx LY, Folegatti PM, Aley PK, et al. Safety and efficacy of the ChAdOx1 nCoV-19 vaccine (AZD1222) against SARS-CoV-2: an interim analysis of four randomised controlled trials in Brazil, South Africa, and the UK. Lancet. 2021;397(10269):99-111.
14. BBC News. Covid: Oxford-AstraZeneca vaccine to be tested on children BBC News2021 [Available from: https://www.bbc.com/news/uk-56052673.
15. Global Times. Sinovac's vaccine proven safe and effective in several countries [Available from: https://www.globaltimes.cn/page/202101/1212727.shtml.
16. Sinovac Research and Development Co. L. NCT04551547 Safety and immunogenicity study of inactivated vaccine for prevention of COVID-19: clinicaltrials.gov; [Available from: https://clinicaltrials.gov/ct2/show/NCT04551547.
17. HKUMed. HKUMed updates on real-time population data on vaccine confidence [Available from: http://www.med.hku.hk/en/news/press/20210128-real-time-population-data-on-vaccine-confidence.
18. Kelvin Kai-Wang To, Ivan Fan-Ngai Hung, Jonathan Daniel Ip, et al, Coronavirus Disease 2019 (COVID-19) Re-infection by a Phylogenetically Distinct Severe Acute Respiratory Syndrome Coronavirus 2 Strain Confirmed by Whole Genome Sequencing, Clinical Infectious Diseases, 2020;, ciaa1275, https://doi.org/10.1093/cid/ciaa1275
19. Victoria Jane Hall, Sarah Foulkes, Andre Charlett, et al. SARS-CoV-2 infection rates of antibody-positive compared with antibody-negative health-care workers in England: a large, multicentre, prospective cohort study (SIREN). Lancet 2021; 397: 1459–69. April 9, 2021 https://doi.org/10.1016/ S0140-6736(21)00675-9
20. Catherine J. Reynolds, Corinna Pade, Joseph M. Gibbons, et al. Prior SARS-CoV-2 infection rescues B and T cell responses to variants after first vaccine dose. Science 2021. 10.1126/science.abh1282 (2021).
21. Carmen Camara, Daniel Lozano-Ojalvo, Eduardo Lopez-Granados, et al. Differential effects of the second SARS-CoV-2 mRNA vaccine dose on T cell immunity in naïve and COVID-19 recovered individuals. bioRxiv 2021. doi: https://doi.org/10.1101/2021.03.22.436441.
22. Isabelle Meyts, Giorgia Bucciol, Isabella Quinti, et al. Coronavirus disease 2019 in patients with inborn errors of immunity: An international study. J Allergy Clin Immunol 2021;147:520- 31.
23. Wood WA, Neuberg DS, Thompson JC, Tallman MS, Sekeres MA, Sehn LH, Anderson KC, Goldberg AD, Pennell NA, Niemeyer CM, Tucker E, Hewitt K, Plovnick RM, Hicks LK. Outcomes of patients with hematologic malignancies and COVID-19: a report from the ASH Research Collaborative Data Hub. Blood Adv. 2020 Dec 8;4(23):5966-5975. doi: 10.1182/bloodadvances.2020003170.
24. Strangfeld A, Schäfer M, Gianfrancesco MA, et al Factors associated with COVID-19-related death in people with rheumatic diseases: results from the COVID-19 Global Rheumatology Alliance physician-reported registry. Annals of the Rheumatic Diseases Published Online First: 27 January 2021. doi: 10.1136/annrheumdis-2020-219498
25. Louise Malle, Cynthia Gao, Chin Hur, et al. Individuals with Down syndrome hospitalized with COVID-19 have more severe disease. Genetics in Medicine (2021) 23:576–580; https://doi.org/10.1038/s41436- 020-01004-w
26. QIAN ZHANG, PAUL BASTARD, ZHIYONG LIU, et al. Inborn errors of type I IFN immunity in patients with life-threatening COVID-19. Science 2021. DOI: 10.1126/science.abd4570.
27. PAUL BASTARD, LINDSEY B. ROSEN, QIAN ZHANG. Autoantibodies against type I IFNs in patients with life-threatening COVID-19. Science 2021. DOI: 10.1126/science.abd4585
28. Lee PY, Platt CD, Weeks S, et al. Immune dysregulation and multisystem inflammatory syndrome in children (MIS-C) in individuals with haploinsufficiency of SOCS1. J Allergy Clin Immunol. 2020 Nov;146(5):1194-1200.e1. doi: 10.1016/j.jaci.2020.07.033. Epub 2020 Aug 25. PMID: 32853638; PMCID: PMC7445138.
29. Bina Choi, Manish C. Choudhary, James Regan, et al. Persistence and Evolution of SARS-CoV-2 in an Immunocompromised Host. N Engl J Med 2021 383;23. DOI: 10.1056/NEJMc2031364.
30. Katarina Braun, Gage Moreno, Cassia Wagner, et al. Limited within-host diversity and tight transmission bottlenecks limit SARS-CoV-2 evolution in acutely infected individuals. bioRxiv 2021.04.30.440988; doi: https://doi.org/10.1101/2021.04.30.440988.
31. Susan S. Chiu, J.S. Malik Peiris, Kwok H. Chan, Wilfred Hing Sang Wong, Yu Lung Lau. Immunogenicity and Safety of Intradermal Influenza Immunization at a Reduced Dose in Healthy Children Pediatrics Jun 2007, 119 (6) 1076-1082; DOI: 10.1542/peds.2006-3176

32. Turner PJ, Ansotegui IJ, Campbell DE, et al. COVID-19 vaccine-associated anaphylaxis: A statement of the World Allergy Organization Anaphylaxis Committee. World Allergy Organ J. 2021;14(2):100517. doi:10.1016/j.waojou.2021.100517
33. The Hong Kong College of Paediatricians and the Hong Kong Society for Paediatric Immunology, Allergy and Infectious Disease. 2021. Joint Position Statement on BioNTech Vaccination in Adolescents with Allergic Diseases.

#### Phase 3 data for the study drugs and benefit-risk assessment

Preliminary efficacy data in phase 3 trials are summarised below.

It is not recommended to directly compare vaccine efficacy estimates for various vaccines as the trial design (e.g. location, participant characteristics) and definition of cases for each vaccine efficacy estimate are different for each trial. In addition to the data below, vaccine efficacy trials on CoronaVac are also underway in Chile, Turkey and Indonesia, yet the full data have not been disclosed.

| **Vaccine** | **AZD1222** | **BNT162b2** | **CoronaVac (Brazil)** |
|---|---|---|---|
| **Number of participants analyzed in study vs placebo arms** | 5807 vs 5829 | 21669 vs 21686 | 4953 vs 4870 |
| **Vaccine efficacy against... (95% CI) and number of cases(/number of participants) in study vs placebo arms** | 55.7% (41.1,66.7)<br>37 vs 112<br>any infections, including asymptomatic<br><br>67.1% (52.3, 77.3)<br>68 vs 153<br>any symptomatic infections<br><br>0 vs 10<br>hospitalization | 94.8% (89.8, 97.6)<br>any symptomatic infections<br><br>1 vs 3<br>severe | 50.7% (35.6, 62.1)<br>85 vs 168<br>any symptomatic infections<br><br>83.7% (58.0, 93.6)<br>5 vs 30<br>symptomatic infections needing treatment<br><br>100% (56, 100)<br>0 vs 10<br>hospitalization |

CoronaVac and BNT162b2 have been trialled successfully in the paediatric population. Pfizer and BioNTech have announced favorable safety and 100% efficacy of the BNT162b2 vaccine in 2260 adolescents aged 12-15 years. SinoVac has also finished a phase ½ trial of CoronaVac in more than 500 children aged 3-17 years, finding favorable safety and higher antibody levels than adult recipients.

| **Vaccine** | **AZD1222** | **BNT162b2** | **CoronaVac** |
|---|---|---|---|
| **Phase** | N/A | Phase 2/3 | Phase 1/2 |
| **Number of participants analyzed in study vs placebo arms** | N/A | 1131 vs 1129<br>12-15y | 72 vs 36<br>12-17y<br>at adult dose |
| Antibody level | N/A | 1239.5 geometric mean neutralizing | 146 geometric mean neutralizing titre |

| | | | |
|---|---|---|---|
| | | titre | |
| Vaccine efficacy against … (95% CI) and number of cases (/participants) in study vs placebo arms | N/A | 100%<br>0 vs 18<br>any symptomatic infections | N/A |

Known reactions to the 3 vaccines are summarised below.

| Rate | AZD1222 | BNT162b2 | CoronaVac |
|---|---|---|---|
| >1/10 | tenderness, pain, warmth, itching or bruising where the injection is given<br>generally feeling unwell<br>feeling tired (fatigue)<br>chills or feeling feverish<br>headache<br>feeling sick (nausea)<br>joint pain or muscle ache | pain and swelling at injection site<br>fatigue<br>headache<br>myalgia<br>arthralgia<br>chills and feverish | pain at injection site<br>headache<br>fatigue |
| <=1/10 | swelling, redness or a lump at the injection site<br>fever<br>being sick (vomiting) or diarrhoea<br>flu-like symptoms, such as high temperature, sore throat, runny nose, cough and chills | redness at injection site<br>nausea | swelling, pruritus, erythema, induration at injection site<br>myalgia<br>nausea<br>diarrhea<br>arthralgia<br>cough<br>chills<br>pruritus<br>loss of appetite<br>rhinorrhea<br>sore throat<br>nasal congestion<br>abdominal pain |
| <=1/100 | feeling dizzy<br>decreased appetite<br>abdominal pain<br>enlarged lymph nodes<br>excessive sweating, itchy skin or rash | swollen lymph node<br>malaise<br>painful limbs<br>insomnia<br>itch at injection site | burn at injection site<br>vomit<br>hypersensitivity<br>abnormal skin and mucosa<br>fever<br>tremor<br>flushing<br>edema<br>dizziness<br>drowsiness |
| <=1/1000 | nil | Bell's palsy | muscle spasms<br>eyelid edema<br>nasal congestion<br>abdominal distension<br>constipation |

| | | | |
|---|---|---|---|
| | | | hyposmia<br>ocular congestion<br>hot flashes<br>hiccup<br>conjunctival congestion |
| Not known | severe allergic reaction (anaphylaxis) | severe allergic reaction (anaphylaxis) | |

In phase 3 data so far available, all 3 vaccines demonstrate a reduction of COVID-19 by more than 50%, and both BNT162b2 and CoronaVac showed no serious safety concern and no evidence of vaccine-mediated severe infection. Therefore, we anticipate the benefits of trialling the vaccines in adolescent participants far outweigh the harm of doing so, and find that there may be inadequate clinical equipoise to necessitate this immunogenicity trial to be controlled with a placebo arm.

Reference

Voysey M, Clemens SAC, Madhi SA, Weckx LY, Folegatti PM, Aley PK, et al. Safety and efficacy of the ChAdOx1 nCoV-19 vaccine (AZD1222) against SARS-CoV-2: an interim analysis of four randomised controlled trials in Brazil, South Africa, and the UK. Lancet. 2021;397(10269):99-111.

Polack FP, Thomas SJ, Kitchin N, Absalon J, Gurtman A, Lockhart S, et al. Safety and Efficacy of the BNT162b2 mRNA Covid-19 Vaccine. N Engl J Med. 2020;383(27):2603-15.

Zhang Y, Zeng G, Pan H, Li C, Hu Y, Chu K, et al. Safety, tolerability, and immunogenicity of an inactivated SARS-CoV-2 vaccine in healthy adults aged 18-59 years: a randomised, double-blind, placebo-controlled, phase 1/2 clinical trial. Lancet Infect Dis. 2021;21(2):181-92.

#### Post-marketing data for the study drugs and benefit-risk assessment

Mass vaccination with the BioNTech/Pfizer mRNA vaccine is rapidly underway in Israel which allows post-marketing observational study of the real-world effectiveness of the vaccine. In a cohort with 596,618 vaccinated individuals, documented SARS-CoV2 infection is found to be reduced by 92% 7 days after a second dose compared to a control cohort of the same size matched for a large range of baseline characteristics over the same period. This study confirms the findings of very high efficacy in the Pfizer trials. Similar data are also available and published by multiple sources, demonstrating high effectiveness of the BNT162b2 vaccine.

In a preprint reporting a national prospective cohort study of 5.4 million people from Scotland, first dose of BNT162b2 and AZD1222 vaccines alone are found to result in 85% and 94% reduction in hospitalizations from COVID-19.

A test-negative case-control study in Manaus, Brazil conducted by American and Brazilian researchers looked into the effectiveness of a single dose of CoronaVac in a setting of high pre-existing seroprevalence and current P.1 variant transmission. At least one dose of CoronaVac is found to reduce symptomatic COVID-19 by 49.6%.Since the widespread use of AZD1222 under emergency use authorization worldwide, cases of unusual thrombotic events associated with thrombocytopenia have been reported. The European Medicines Agency has confirmed a link between the vaccine and a rare thrombotic event, i.e. cerebral venous sinus thrombosis (CVST). Such cases are thought to be immune-mediated, with platelet-activating antibodies against PF4 detected in 11 patients in Germany and Austria with thrombosis, such as CVST, splanchnic vein thrombosis, pulmonary embolism, or thrombocytopenia beginning 5 to 16 days after vaccination.

Such conditions mimic heparin-induced thrombocytopenic thrombosis, and those affected are commonly middle aged. The UK has since then recommended individuals under age of 30 to receive an alternative vaccine. An originally ongoing paediatric trial of AZD1222 by AstraZeneca/Oxford has been temporarily halted as such, despite no safety concerns arising from trial participants. In Hong Kong, delivery of AZD1222 doses ordered has been cancelled by the government.

mRNA vaccines have been determined to associate with myopericarditis, especially in young recipients. Local data suggests a rate of 1 in 2000 for boys aged 11-17 (unpublished). As the benefit of a second dose is unclear locally, the government has postponed giving second dose for people aged below 18 receiving BNT162b2. The mechanism of the disease is unclear, and participants have been given relevant information in this study.

Reference: Dagan, N., Barda, N., et al. BNT162b2 mRNA Covid-19 vaccine in a Nationwide Mass Vaccination Setting. N Engl J Med 2021. DOI: 10.1056/NEJMoa2101765.

Vasileiou E, et al. Effectiveness of first dose of COVID-19 vaccines against hospital admissions in Scotland: national prospective cohort study of 5.4 million people. SSRN 2021.

Hitchings M.D.T, et al. Effectiveness of CoronaVac in the setting of high SARS-CoV2 P.1 variant transmission in Brazil: A test-negative case-control study. medRxiv 2021.

European Medicines Agency Pharmacovigilance Risk Assessment Committee. Signal assessment report on embolic and thrombotic events with COVID-19 vaccine (ChAdOx1-S [recombinant]) - COVID-19 vaccine AstraZeneca (Other viral vaccines). 2021.

Greinacher A, et al. Thrombotic Thrombocytopenia after ChAdOx1 nCov-19 vaccination. N Engl J Med 2021.

Schultz N.H., et al. Thrombosis and Thrombocytopenia after ChAdOx1 nCoV-19 Vaccination. N Engl J Med 2021.

##### *6b Explanation for choice of comparators*

Participants in one study arm will be compared to the other two study arms. There is no placebo control arm.

#### Objectives

##### *7 Specific objectives or hypotheses*

##### Aims

1. To measure and track immune responses such as anti-SARS-CoV-2-spike-protein IgG levels and related markers, neutralizing antibody titres and B and T cell recall memory responses for 3 years after vaccine dose 2/3 in children and parents/adults
2. To assess proactively the reactogenicity daily for 7 days post vaccine doses 1 and 2 (and 3), and document adverse events throughout the 3-year study period
3. To compare the immunogenicity across the vaccines used—BNT162b2 and CoronaVac —for both children and parents or unrelated adults to establish immunobridging
4. To compare the reactogenicity and immunogenicity between each vaccine
5. To explore any correlates of inducing serological and cellular response and memory from vaccination
6. To explore the psychosocial impacts of COVID-19 vaccination on adolescents and children

7. To assess the reactogenicity and immunogenicity of the vaccines in adolescents and children with pre-existing conditions, such as prior COVID-19 and paediatric immunocompromising illnesses
8. To compare the immunogenicity of intramuscular and intradermal CoronaVac immunization in children and adolescents

#### Objectives

*Primary objectives*

1. Frequencies of reactogenicity within the 7 days after vaccine injection

2. Compare frequencies of reactions among the vaccines, including across doses, routes and types

3. Short-term and long-term (up to 36 months post-vaccination) longitudinal differences in spike protein IgG and related markers, neutralizing antibody titres and B and T cell immune responses

4. Compare immunogenicity outcomes between children and adults, such that the geometric mean values of immunological parameters are statistically non-inferior (0.6 of adult value), and differences between vaccines

*Secondary objectives*

5. Describe all adverse events and serious adverse events during the study period

6. Explore any parameters or markers predictive and correlative of immune responses to vaccination.

7. Short-term and long-term (up to 36 months post-vaccination) longitudinal differences in non-spike protein antibody titres and related markers for subjects receiving CoronaVac;

8. Short-term and long-term (up to 36 months post-vaccination) longitudinal differences in type I and II T cell immune response

9. Differences in short-term and long-term (up to 36 months post-vaccination) longitudinal differences in spike protein IgG and related markers, neutralizing antibody titres and cellular immune responses against emerging viral variants between vaccines, and between children and adults;

10. Vaccine breakthroughs as documented by N and ORF8 antibodies in subjects receiving BNT162b2, or ORF8 in subjects receiving CoronaVac, with estimation of potential immunological correlates of protection against infection.

11. Explore psychosocial impacts of COVID-19 vaccination in adolescents and children

12. Impact of pre-existing conditions on safety, reactogenicity and immunogenicity of vaccines

13. Impact of vaccination on adolescents with previous COVID-19 and long COVID

14. Short-term and long-term (up to 36 months post-vaccination) longitudinal differences in SARS-CoV2 whole blood immune responses

15. Short-term and long-term (up to 36 months post-vaccination) longitudinal differences in SARS-CoV2 nasopharyngeal immune responses

16. Estimate vaccine efficacy based on established immune correlates of protection

17. Compare immunogenicity of intradermal and intramuscular CoronaVac immunization

#### Trial design

*8 Description of trial design including type of trial (eg, parallel group, crossover, factorial, single group), allocation ratio, and framework (eg, superiority, equivalence, noninferiority, exploratory)*

##### Type of trial

Parallel

##### Allocation ratio

1:1 (in terms of number of participants by vaccine type)

##### Framework

Non-inferiority (and exploratory)

### Methods: Participants, interventions, and outcomes

#### Study setting

*9 Description of study settings (eg, community clinic, academic hospital) and list of countries where data will be collected. Reference to where list of study sites can be obtained*

##### Study setting

Vaccination site
Ap Lei Chau Vaccination Center, Ap Lei Chau, Hong Kong. (Community vaccine clinic)

Gleneagles Community Vaccination Center, Wong Chuk Hang, Hong Kong. (Community vaccine clinic)

Sun Yat Sen Memorial Park Sports Center Community Vaccination Center, Sai Ying Pun, Hong Kong. (Community vaccine clinic)

Follow-up
Queen Mary Hospital, Pokfulam, Hong Kong. (Academic hospital)

#### Eligibility criteria

*10 Inclusion and exclusion criteria for participants. If applicable, eligibility criteria for study centres and individuals who will perform the interventions (eg, surgeons, psychotherapists)*

##### Criteria for healthy participants (subgroup CA)

Inclusion criteria for participants

1. informed consent for adult participants or from the parents or a legally acceptable representative for an underage participant and assent for an underage participant aged 7 years or above
2. for students, aged 3-17 years (or at least 11 years for BNT162b2), inclusive, at time of vaccination or (biological) parents or relatives of students enrolled in the trial or healthy unrelated adults
3. ability to adhere to the follow-up schedules
4. willingness to report reactogenicity daily for 7 days post each dose proactively
5. willingness to receive that vaccine available for that particular recruitment period (as student-parent pair, if applicable)
6. good past health, including pre-existing clinically stable disease

Exclusion criteria for participants

7. known history of COVID-19
8. receipt of coronavirus vaccines in the past (apart from COVID-19 vaccination, if to receive further dosing or perform blood taking only)
9. history of severe allergy, such as angioedema, bronchospasm and/or hypotension to food, drug, vaccine, or unknown trigger

10. history of Steven-Johnson syndrome (SJS), toxic epidermal necrolysis (TEN), drug rash with eosinophilia and systemic syndrome (DRESS)/drug-induced hypersensitivity syndrome (DIHS)
11. any significant medical or psychiatric condition including laboratory abnormalities or suicidal ideation and behaviour within past 12 months that may increase risk of vaccination
12. history of severe neurological conditions, e.g. transverse myelitis, Guillain-Barre Syndrome (GBS), demyelinating diseases
13. severe immunocompromise, including present or planned use of cytotoxic agents or systemic immunosuppressives, known primary or secondary immunodeficiency diseases. Short-term systemic steroids use (<14 days) for an acute condition precludes the participant from receiving study vaccines until 28 days after the termination of said therapy
14. receipt of blood or plasma products or immunoglobulin within 60 days prior to vaccination to conclusion of the study
15. any haematological diseases (that preclude intramuscular injection)
16. reported pregnancy or breastfeeding

Criteria for temporarily delaying enrollment/intervention administration of participants

17. Any acute illness or symptoms within 48 hours before vaccination that may suggest a potential COVID-19 illness, including new or increased respiratory symptoms, chills, new or increased myalgia, new loss of smell or taste, new gastrointestinal distress, fever (>= 37.5 degrees Celsius) and any other signs as judged by investigators
18. Short-term systemic steroids use (<14 days) for an acute condition precludes the participant from receiving study vaccines until 28 days after the termination of said therapy

**Criteria for joining the observational immunogenicity (and safety) aspect of this study only with vaccination already performed outside of this study (CA parents or reference controls)**

Inclusion

1. informed consent for adult participants or from the parents or a legally acceptable representative for an underage participant and assent for an underage participant
2. ability to adhere to the follow-up schedules for every visit subsequent to enrollment in the study
3. have completed/soon to complete two doses of BNT162b2 with the second dose given within 21-28 days after the first outside of this trial; or have completed/soon to complete two doses of CoronaVac with the second dose given within 28-35 days after the first outside of this trial

Exclusion

1. known history of COVID-19
2. history of severe allergy, such as angioedema, bronchospasm and/or hypotension to food, drug, vaccine, or unknown trigger
3. history of Steven-Johnson syndrome (SJS), toxic epidermal necrolysis (TEN), drug rash with eosinophilia and systemic syndrome (DRESS)/drug-induced hypersensitivity syndrome (DIHS)
4. severe immunocompromise, including present or planned use of cytotoxic agents or systemic immunosuppressives, known primary or secondary immunodeficiency diseases
5. receipt of blood or plasma products or immunoglobulin within 60 days prior to vaccination to conclusion of the study
6. short-term systemic steroids use (<14 days) for an acute condition within 28 days before vaccination

#### Criteria for patients with previous COVID-19 (subgroup CS)

Inclusion

1. informed consent for adult participants or from the parents or a legally acceptable representative for an underage participant and assent from an underage participant aged 7 years or above
2. age 3 years or above at the time of dose 1 (or at least 11 years for BNT162b2)
3. ability to adhere to follow-up schedule for every visit subsequent to enrollment in this study
4. willingness to report reactogenicity daily for 7 days post dose proactively
5. willingness to receive that vaccine available for that particular recruitment period (as student-parent pair, if applicable)
6. willingness to participate as a child-parent pair or a family unit, in which all participants have had a history of microbiologically diagnosed COVID-19, if possible
7. good past health, including pre-existing clinically stable disease

Exclusion

8. receipt of coronavirus vaccines in the past (apart from COVID-19 vaccination, if to receive further dosing or perform blood taking only)
9. history of severe allergy, such as angioedema, bronchospasm and/or hypotension to food, drug, vaccine, or unknown trigger
10. history of Steven-Johnson syndrome (SJS), toxic epidermal necrolysis (TEN), drug rash with eosinophilia and systemic syndrome (DRESS)/drug-induced hypersensitivity syndrome (DIHS)
11. any significant medical or psychiatric condition including laboratory abnormalities or suicidal ideation and behaviour within past 12 months that may increase risk of vaccination
12. history of severe neurological conditions, e.g. transverse myelitis, Guillain-Barre Syndrome (GBS), demyelinating diseases
13. severe immunocompromise, including present or planned use of cytotoxic agents or systemic immunosuppressives, known primary or secondary immunodeficiency diseases
14. receipt of blood or plasma products or immunoglobulin during the study
15. any haematological diseases (that preclude intramuscular injection)
16. reported pregnancy or breastfeeding

Delay vaccination

17. hospitalization or acute COVID-19-related episode within 6 months prior to vaccination
18. short-term steroid or immunomodulating drug use for fewer or equal to 14 days within 28 days prior to vaccination
19. receipt of blood or plasma products or immunoglobulin within 60 days prior to vaccination
20. any acute illness or symptoms within 48 hours prior to vaccination that may suggest a potential COVID-19 illness, including new or increased respiratory symptoms, chills, new or increased myalgia, new loss of smell or taste, new gastrointestinal distress, fever (>= 37.5 degrees Celsius) and any other signs as judged by investigators

#### Criteria for patients with severe immune compromise or paediatric conditions

Inclusion

1. informed consent for adult participants or from the parents or a legally acceptable representative for an underage participant and assent from an underage participant aged 7 years or above
2. At least 5 years for BNT162b2
3. ability to adhere to follow-up schedule for every visit subsequent to enrollment in this study
4. willingness to report reactogenicity daily for 7 days post dose proactively
5. willingness to receive that vaccine available for that particular recruitment period
6. a history of severe paediatric conditions, e.g. inborn errors of immunity, paediatric solid or blood tumor patients, immune dysregulation disorders, various congenital diseases

Exclusion

7. receipt of coronavirus vaccines in the past (apart from COVID-19 vaccination, if to receive further dosing or perform blood taking only)
8. any unstable significant medical or psychiatric condition including laboratory abnormalities or suicidal ideation and behaviour that may increase risk of vaccination
9. reported pregnancy or breastfeeding

Delay vaccination

10. recent unstable disease, including hospitalization due to acute disease onset or exacerbation within 90 days prior to vaccination
11. recent medication change within 90 days prior to vaccination
12. recent surgery within past 12 months
13. any acute illness or symptoms within 48 hours prior to vaccination that may suggest a potential COVID-19 illness, including new or increased respiratory symptoms, chills, new or increased myalgia, new loss of smell or taste, new gastrointestinal distress, fever (>= 37.5 degrees Celsius) and any other signs as judged by investigators

#### Interventions

*11a Interventions for each group with sufficient detail to allow replication, including how and when they will be administered*

##### Preparation, dispensing and administration of vaccines

Dispensing

Vaccines will be prepared and dispensed by a qualified and experienced medical staff member at the community vaccination clinic.

Vaccination

Vaccines will be given intramuscularly, unless otherwise specified, to the deltoid muscle (of the nondominant arm preferably) by clinical staff experienced with vaccination. A separate study investigator or trial staff will monitor the vaccination, and the date and time of vaccination will be documented.

Scheduling of 2nd and any subsequent booster doses may be modified according to the latest local government recommendations, based on the latest scientific evidence.

Participants should not premedicate solely to prevent adverse reactions from the vaccine, e.g. taking paracetamol before the onset of fever.

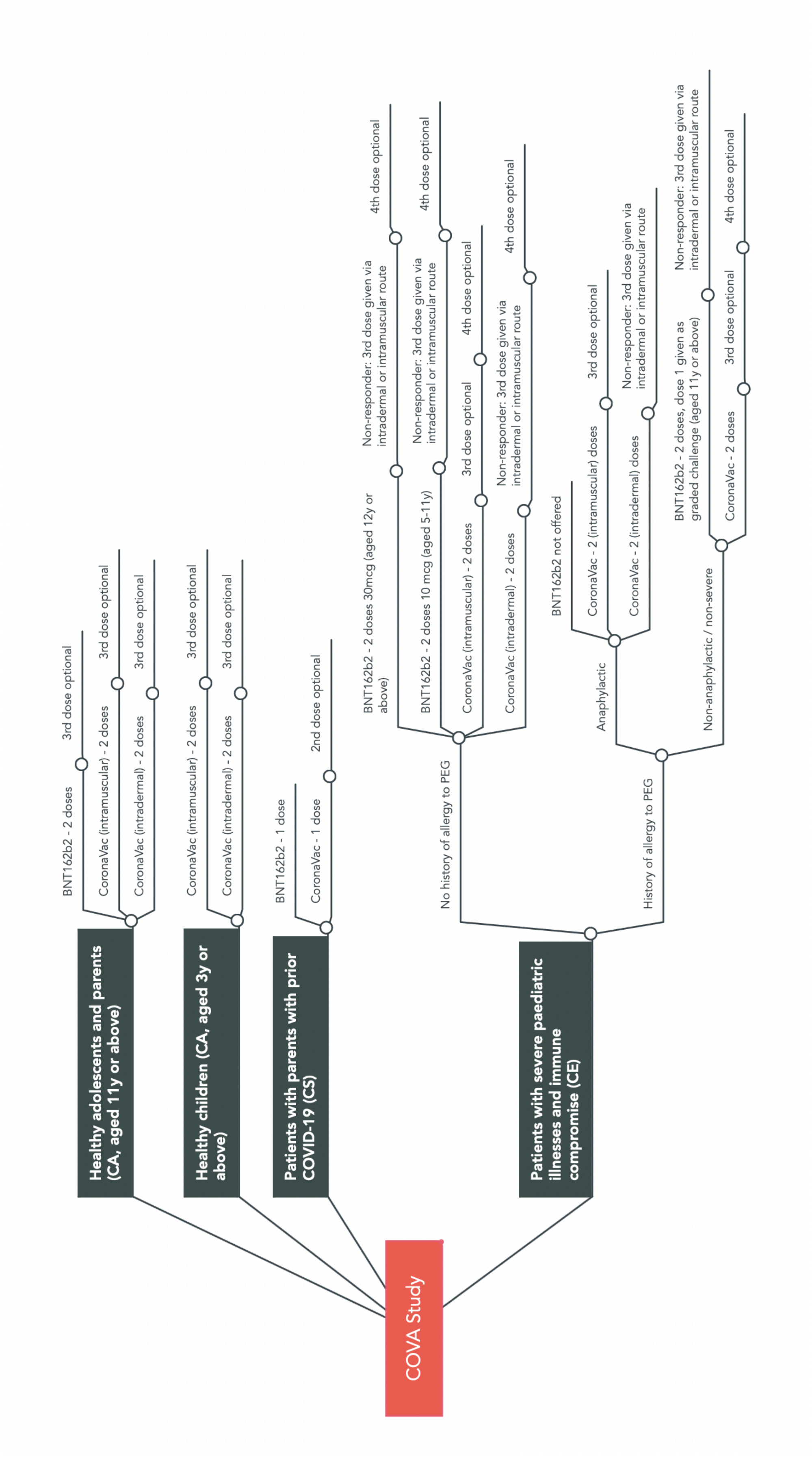
COVA Study
Healthy adolescents and parents (CA, aged 11y or above)
BNT162b2 - 2 doses
3rd dose optional
CoronaVac (intramuscular) - 2 doses
3rd dose optional
CoronaVac (intradermal) - 2 doses
3rd dose optional
Healthy children (CA, aged 3y or above)
CoronaVac (intramuscular) - 2 doses
3rd dose optional
CoronaVac (intradermal) - 2 doses
3rd dose optional
Patients with parents with prior COVID-19 (CS)
BNT162b2 - 1 dose
CoronaVac - 1 dose
2nd dose optional
Patients with severe paediatric illnesses and immune compromise (CE)
No history of allergy to PEG
BNT162b2 - 2 doses 30mcg (aged 12y or above)
Non-responder: 3rd dose given via intradermal or intramuscular route
4th dose optional
BNT162b2 - 2 doses 10 mcg (aged 5-11y)
Non-responder: 3rd dose given via intradermal or intramuscular route
4th dose optional
CoronaVac (intramuscular) - 2 doses
3rd dose optional
4th dose optional
CoronaVac (intradermal) - 2 doses
Non-responder: 3rd dose given via intradermal or intramuscular route
4th dose optional
History of allergy to PEG
Anaphylactic
BNT162b2 not offered
CoronaVac - 2 (intramuscular) doses
3rd dose optional
CoronaVac - 2 (intradermal) doses
Non-responder: 3rd dose given via intradermal or intramuscular route
Non-anaphylactic / non-severe
BNT162b2 - 2 doses, dose 1 given as graded challenge (aged 11y or above)
Non-responder: 3rd dose given via intradermal or intramuscular route
CoronaVac - 2 doses
3rd dose optional
4th dose optional

##### Arm CA-B

Study article
BioNTech-Fosun COVID-19 vaccine (also known as tozinameran, BNT162b2)

Description
modified messenger RNA vaccine

Route and dosing
2 doses of 30ug/0.3ml per dose given by intramuscular injection given 21 (or up to 28) days apart; dose 3 optional and at least 84 days after second dose

Age
11 years or above at time of dose 1

##### Arm CE-B/g (Patients with possible history of allergy to polyethylene glycol or first dose of BNT162b2)

Study article
BioNTech-Fosun COVID-19 vaccine (also known as tozinameran, BNT162b2)

Description
modified messenger RNA vaccine

Route and dosing
First dose given in a graded challenge of full-strength 0.03ml, 0.12ml and 0.15ml with 60 minute observation apart and 60 minute observation at the end by intramuscular injection, or full strength 0.03ml and 0.27ml with 60 minutes after each injection; followed up by 1 dose of 30ug/0.3ml given by intramuscular injection given 21 (or up to 28) days later. For those with allergic reaction to dose 1 BNT162b2, dose 2 may be given by graded challenge. Dose 3 optional and at least 84 days after second dose

Age
11 years or above at time of dose 1

#### Arm CE-B/4 (Patients with immune compromise aged 11 years or above)

Study article
BioNTech-Fosun COVID-19 vaccine (also known as tozinameran, BNT162b2)

Description
modified messenger RNA vaccine

Route and dosing
4 doses of 30ug/0.3ml given by intramuscular injection on day 0, day 21-28, day 49-175 and day 133-343; 3rd dose may be given by intradermal injection if opted by patient

Age
11 years or above at time of dose 1

#### Arm CE-B/p (Patients with immune compromise aged 5-11 years)

Study article
BioNTech-Fosun COVID-19 vaccine (also known as tozinameran, BNT162b2)

Description
modified messenger RNA vaccine

Route and dosing
4 doses of 10ug/0.1ml given by intramuscular injection on day 0, day 21-28, day 49-175 and day 133-343

Age
5-11 years at time of dose 1

#### Arms CA-C and CE-C

Study article
CoronaVac by SinoVac

Description
inactivated virus vaccine

Route and dosing
2 doses of 600SU/0.5ml per dose given by intramuscular injection given 28 (or up to 35) days apart

#### Arms CA-C/3 and CE-C/3 (Any CoronaVac recipients not previously infected who opt to receive third dose)

Study article
CoronaVac by SinoVac (and BioNTech-Fosun COVID-19 vaccine, also known as tozinameran, BNT162b2)

Description
inactivated virus vaccine (and modified messenger RNA vaccine)

Route and dosing
3 doses of 600SU/0.5ml per dose given by intramuscular injection given on day 0, day 28-35 and day 56 or after; 3rd dose may be given by intradermal injection in CE group if opted by immunocompromised patient; alternative vaccine (BNT162b2, 30ug/0.3ml given by intramuscular injection) may be given as dose 3 upon request by participant

#### Arm CA-C/I

Study article
CoronaVac by SinoVac

Description
inactivated virus vaccine

Route and dosing
2 or 3 doses of 600SU/0.5ml per dose given by intradermal injection (Mantoux technique or MicronJet intradermal injector) given on days 0, 28 and 56+

#### Arm CE-C/I

Study article
CoronaVac by SinoVac

Description
inactivated virus vaccine

Route and dosing
3 or 4 doses of 600SU/0.5ml per dose given by intradermal injection (Mantoux technique or MicronJet intradermal injector) given on days 0, 28, 56+; 4th dose to be given 84-168 days after 3rd dose

##### Arm CS-B/1 (COVID survivors opting for one dose of BNT162b2)

Study article
BioNTech-Fosun COVID-19 vaccine (also known as tozinameran, BNT162b2)

Description
modified messenger RNA vaccine

Route and dosing
1 dose of 30ug/0.3ml per dose given by intramuscular injection

##### Arm CS-B/2 (COVID survivors opting for 2 doses of BNT162b2)

Study article
BioNTech-Fosun COVID-19 vaccine (also known as tozinameran, BNT162b2)

Description
modified messenger RNA vaccine

Route and dosing
2 dose of 30ug/0.3ml per dose given by intramuscular injection at least 180 days apart

##### Arm CS-C/1 (COVID survivors opting for one dose of CoronaVac)

Study article
CoronaVac by SinoVac

Description
inactivated virus vaccine

Route and dosing
1 dose of 600SU/0.5ml per dose given by intramuscular injection

**Arm CS-C/2 (COVID survivors opting for 2 doses of CoronaVac)**

Study article
CoronaVac by SinoVac

Description
inactivated virus vaccine

Route and dosing
2 doses of 600SU/0.5ml per dose given by intramuscular injection given 28 to 147 days apart

#### *11b Criteria for discontinuing or modifying allocated interventions for a given trial participant (eg, drug dose change in response to harms, participant request, or improving/worsening disease)*

##### Criteria for discontinuing intervention for a trial participant

Investigators responsible for clinical oversight may withhold further dosing for a trial participant if any of the following criteria are met. The exact reason will be documented in the study database.

- Becomes ineligible for the study according to exclusion criteria
- Experiences a severe adverse event considered to be related to intervention given
- Experiences an adverse event, which may or may not be related to intervention given, and prompts investigators to withhold further dosing
- Experiences a clinically abnormal vital sign reading or physical examination finding, or any general condition, which may or may not be related to intervention given, and prompts investigators to withhold further dosing
- Tests positive serology for SARS-CoV2 prior to vaccination
- Participant request to discontinue intervention

##### Criteria for modifying intervention for a trial participant

Further doses of vaccine may be modified given the respective conditions are fulfilled:

1. Upon participant request and agreement by investigators, subsequent dose of the same vaccine is refused unless alternative vaccine is given
2. At the judgement of the investigator that the participant is medically unsuitable to receive a second dose of the same vaccine, such as because of anaphylaxis to dose 1, with the

agreement by participants or their parents/legally acceptable representatives as needed, the alternative vaccine may be given as dose 2

3. At the judgement of the investigator that the participant is medically unsuitable to receive a second dose of the same vaccine, such as because of anaphylaxis to dose 1, with the agreement by participants or their parents/legally acceptable representatives as needed, the same vaccine may be given by graded administration

##### *11c Strategies to improve adherence to intervention protocols, and any procedures for monitoring adherence (eg, drug tablet return, laboratory tests)*

A separate study investigator or trial staff will monitor the vaccination, and the date and time of vaccination will be documented.

##### *11d Relevant concomitant care and interventions that are permitted or prohibited during the trial*

Prophylactic use of medications, such as antipyretic or pain medicine, intended to prevent the onset of adverse reactions to vaccines is prohibited. Use of medications to treat adverse reactions is permitted.

Item 10 details exclusion criteria, as well as criteria for deferring intervention, e.g. recent or planned receipt of non-study vaccines and recent or planned short-term systemic steroid use.

Concomitant care and interventions will be documented at the time of/prior to vaccinations, with adverse events in 28 days following vaccination, and with severe adverse events throughout the study period.

#### Outcomes

*12 Primary, secondary, and other outcomes, including the specific measurement variable (eg, systolic blood pressure), analysis metric (eg, change from baseline, final value, time to event), method of aggregation (eg, median, proportion), and time point for each outcome. Explanation of the clinical relevance of chosen efficacy and harm outcomes is strongly recommended*

##### Primary outcomes

1a. Percentage of types and severity of adverse reactions occurring within 7 days post-doses 1 and 2 (and 3)

2a. Difference in percentages of types and severity of adverse reactions occurring within 7 days post-doses 1 and 2 (and 3)

3a. Geometric mean OD450 value of SARS-CoV2 S and S RBD-specific binding antibody and related markers as determined by Enzyme-linked Immunosorbent Assay at 1 month post-dose 1, and 1, 6, 18, and 36 months post-vaccination (and 2 weeks post-dose 3)

3b. Geometric mean inhibition level of SARS-CoV2 surrogate virus neutralization test at 1 month post-dose 1, and 1, 6, 18, and 36 months post-vaccination (and 2 weeks post-dose 3)

3c. Geometric mean titre of SARS-CoV2 neutralizing antibodies as determined by plaque reduction neutralization assay at 1 month post-dose 1, and 1, 6, 18, and 36 months post-vaccination (and 2 weeks post-dose 3)

3d. Geometric mean frequency of Th1 and Tc1 cells specific to SARS-CoV2 S (and N and M) protein at 1 month post-dose 1, and 1, 6, 18, and 36 months post-vaccination (and 2 weeks post-dose 3)

3e. Geometric mean OD450 value of SARS-CoV2 S-specific FcgRIIIa-binding antibody as determined by Enzyme-linked Immunosorbent Assay at 1 month post-dose 1, and 1, 6, 18, and 36 months post-vaccination (and 2 weeks post-dose 3)

3f. Geometric mean OD450 value of SARS-CoV2 S-specific binding antibody avidity index as determined by Enzyme-linked Immunosorbent Assay at 1 month post-dose 1, and 1, 6, 18, and 36 months post-vaccination (and 2 weeks post-dose 3)

3g. Geometric mean frequency of SARS-CoV2 S -specific B cells as determined by flow cytometry at 1 month post-dose 1, and 1, 6, 18, and 36 months post-vaccination (and 2 weeks post-dose 3)

4a. Geometric mean ratios of primary immunogenicity outcomes between adults and children

4b. Geometric mean ratios of primary immunogenicity outcomes between vaccines and routes

#### Secondary outcomes

5a. Percentage of types and severity of adverse events and severe adverse events throughout study period

6a. Mean count and percentage of white blood cell subtypes in peripheral blood of participants at time of dose 1

6b. Whole blood cytokine and transcriptomic profiles upon ex vivo stimulation by TLR3, 7-9 ligands of participants at time of dose 1

6c. Nasopharyngeal metagenomic, cytokine, immunoglobulin, IgG allotypes and other proteogenomic profiles at time of dose 1

6d. Demographic, anthropometric and other baseline characteristics of participants

7a. Geometric mean OD450 value of SARS-CoV2 N (and N-CTD)-specific binding antibody as determined by Enzyme-linked Immunosorbent Assay at 1 month post-dose 1, and 1, 6, 18, and 36 months post-vaccination (and 2 weeks post-dose 3)

7b. Geometric mean OD450 value of SARS-CoV2 N-specific FcgRIIIa-binding antibody as determined by Enzyme-linked Immunosorbent Assay at 1 month post-dose 1, and 1, 6, 18, and 36 months post-vaccination (and 2 weeks post-dose 3)

7c. Geometric mean OD450 value of SARS-CoV2 N-specific binding antibody avidity index as determined by Enzyme-linked Immunosorbent Assay at 1 month post-dose 1, and 1, 6, 18, and 36 months post-vaccination (and 2 weeks post-dose 3)

7d. Geometric mean ratios of immunogenicity outcomes between children and adults

8a. Geometric mean frequency of Th2 and Tc2 cells specific to SARS-CoV2 peptide pools at 1 month post-dose 1, and 1, 6, 18, and 36 months post-vaccination (and 2 weeks post-dose 3)

8b. Geometric mean ratio of Th1/Tc1 and Th2/Tc2 cells specific to SARS-CoV2 peptide pools at 1 month post-dose 1, and 1, 6, 18, and 36 months post-vaccination(and 2 weeks post-dose 3)

8c. Geometric mean ratios of such counts/ratios in adults versus adolescent/children vaccinees of same vaccine at various timepoints

8d. Geometric mean ratios of such counts/ratios in vaccinees of different vaccine

9a. Geometric mean OD450 value of SARS-CoV2 variant S RBD-specific binding antibody as determined by Enzyme-linked Immunosorbent Assay at 1 month post-dose 1, and 1, 6, 18, and 36 months post-vaccination (and 2 weeks post-dose 3)

9b. Geometric mean inhibition level of SARS-CoV2 surrogate variant virus neutralization test at 1 month post-dose 1, and 1, 6, 18, and 36 months post-vaccination (and 2 weeks post-dose 3)

9c. Geometric mean titre of SARS-CoV2 neutralizing antibodies as determined by variant plaque reduction neutralization assay at 1 month post-dose 1, and 1, 6, 18, and 36 months post-vaccination (and 2 weeks post-dose 3)

9d. Geometric mean frequency of Th1 and Tc1 cells specific to SARS-CoV2 variant S protein and other peptide pools at 1 month post-dose 1, and 1, 6, 18, and 36 months post-vaccination (and 2 weeks post-dose 3)

10a. Incidence of COVID-19 in participants throughout study period as self-reported or as determined by Luciferase Immunoprecipitation Systems assay

11a. Psychosocial assessments at time of dose 1 and 6, 18 and 36 months post-vaccination

12a. Geometric mean ratios of immunogenicity outcomes between paediatric patients and healthy children

13a. Percentage of types, duration and severity of long COVID-19 abnormalities at time of dose 1 and 1, 6, 18 and 36 months post vaccination

14a. Whole blood cytokine and transcriptomic profiles upon ex vivo stimulation by SARS-CoV2 S protein of participants at 1 month post-dose 2 (and 2 weeks post-dose 3)

15a. Nasopharyngeal immunoglobulin and other immune responses 1 month and 6 months post-vaccination (and 2 weeks post-dose 3)

16. Predicted vaccine efficacy based on mean neutralizing antibody titers

17. Geometric mean ratio of immunogenicity outcomes between intradermal and intramuscular CoronaVac recipients

#### Participant timeline

*13 Time schedule of enrolment, interventions (including any run-ins and washouts), assessments, and visits for participants. A schematic diagram is highly recommended*

##### Enrollment

Promotion, assessment for eligibility and enrollment will be started 4 weeks after the vaccine is first available to the public in Hong Kong. Enrollment for the 3 arms will therefore commence at different times.

- Study arm C (CoronaVac): March 22, 2021 - February 2022
- Study arm B (BioNTech): March 22 – February 2022

##### Intervention

- Study arm C (CoronaVac): June 2021 – May 2022
- Study arm B (BioNTech): May 2021 - May 2022

##### Assessment

Assessment will begin together with intervention, and will last 36 months after the second/last dose of the vaccine is given for each participant.

- Study arm C (CoronaVac): June, 2021 - May 2025 anticipated
- Study arm B (BioNTech): May 2021 - May 2025 anticipated

##### Overall schematic

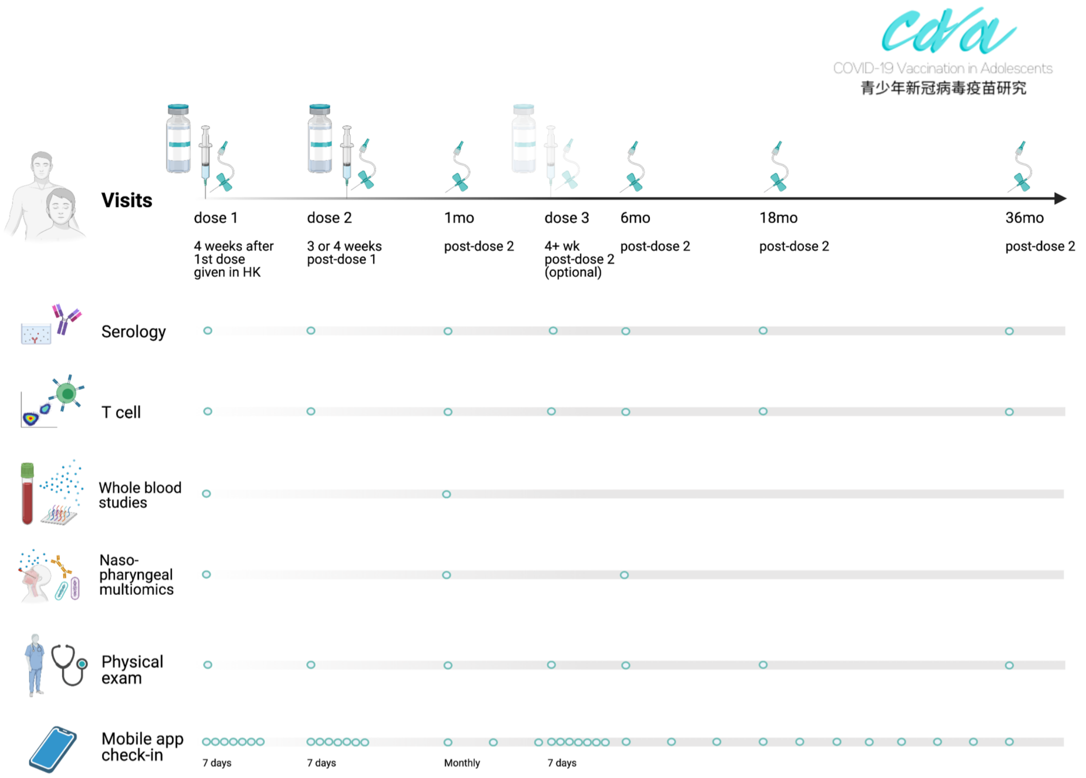


#### Sample size

*14 Estimated number of participants needed to achieve study objectives and how it was determined, including clinical and statistical assumptions supporting any sample size calculations*

##### Subgroup CA (healthy adolescents, children and adults)

When comparing the peak GM immunogenicity outcomes (e.g. PRNT) of children with that of parents, or between vaccine types, a sample size of 61 in each group would assure that a two-sided test with alpha=0.05 has 99% power to detect an effect size with a Cohen's d value=0.78, assuming 1.67-fold response in adolescents on the linear scale, or a difference of 0.51 after natural log transformation, compared to adults and a standard deviation (SD) of 0.65 on the natural log scale within each group. For assays with higher technical requirements, 66 evaluable adolescents and 16 evaluable adults tested would achieve 80% power to detect the same non-inferiority margin with the same alpha and SD. For the proportion of participants with a positive result in immunogenicity outcomes or ARs, 110 adolescents would yield a 95% chance to detect the true value within 7.5% of the measured percentage, assuming a prevalence of 80%. Recruitment of 120 participants were targeted per vaccine regimen and age group to accommodate for attrition.

**Subgroup CE (patients with inborn errors of immunity, immunological or hematological disorders, and other paediatric conditions)**
As paediatric conditions are diverse and individually rare, although collectively affecting a percentage of the population, a target sample size of 100 patients, 50 for each vaccine arm, with different conditions is planned. These patients will be studied both collectively, comparing CE and CA subgroups for the various outcomes in this trial, and individually as ‘n of 1 trials’.

### Recruitment

#### *15 Strategies for achieving adequate participant enrolment to reach target sample size*

Recruitment will be performed via 3 routes:

- Promotion to local secondary and primary schools
- Promotion to previous trial participants of studies by the principal investigator
- Promotion to the general public via mass media and social media

### Methods: Assignment of interventions (for controlled trials)

#### Allocation: Sequence generation

*16a Method of generating the allocation sequence (eg, computer-generated random numbers), and list of any factors for stratification. To reduce predictability of a random sequence, details of any planned restriction (eg, blocking) should be provided in a separate document that is unavailable to those who enrol participants or assign interventions*

Not applicable. This is a nonrandomized trial.

#### Allocation concealment mechanism

*16b Mechanism of implementing the allocation sequence (eg, central telephone; sequentially numbered, opaque, sealed envelopes), describing any steps to conceal the sequence until interventions are assigned*

Not applicable. This is a nonrandomized trial.

#### Implementation

*16c Who will generate the allocation sequence, who will enrol participants, and who will assign participants to interventions*

Not applicable. This is a nonrandomized trial.

#### Blinding (masking)

*17a Who will be blinded after assignment to interventions (eg, trial participants, care providers, outcome assessors, data analysts), and how*

Not applicable. This is a nonrandomized trial.

*17b If blinded, circumstances under which unblinding is permissible, and procedure for revealing a participant's allocated intervention during the trial*

Not applicable. This is a nonrandomized trial.

### Methods: Data collection, management, and analysis

#### Data collection methods

*18a Plans for assessment and collection of outcome, baseline, and other trial data, including any related processes to promote data quality (eg, duplicate measurements, training of assessors) and a description of study instruments (eg, questionnaires, laboratory tests) along with their reliability and validity, if known. Reference to where data collection forms can be found, if not in the protocol*

Study procedures for each scheduled visit are described in the Appendices.

##### First-tier eligibility screening (electronic, self-entered by participants)

Informed consent and assent will be taken electronically or in written form from trial participants or their parents/legally acceptable representatives for underage participants, prior to eligibility screening and any other study procedures.

First-tier eligibility screening will be conducted in an online system for interested potential participants. Key information collected will include:

1. Consent
2. (For 7-17 years) assent
3. Full English and Chinese name
4. Date of birth
5. Sex
6. (For students) School and grade
7. Race
8. Language preferred for communication (English/Chinese)
9. Phone number (WhatsApp or Signal-enabled), email address, home address
10. Known COVID-19
11. Chronic diseases
12. Allergy history
13. Recent (past 28 days), regular or anticipated use of medications and vaccines, including name, purpose and duration
14. Pregnancy and breastfeeding
15. Acute illness or symptoms experienced recently, including new or increased respiratory symptoms, new or increased myalgia, new loss of smell or taste, new gastrointestinal distress, or others as declared by the potential participants
16. Family members joining the study
17. Vaccine intended
18. Ability to adhere to follow-up schedule, including planned residence in Hong Kong for the coming 3 years
19. Willingness to report reactions daily 7 days post-doses

A phone interview will be conducted by investigators for confirmation if the potential participant fails any of the screening questions. Visit 1 will be scheduled with participants who pass first-tier screening, with their family member(s).

Screen failures, reason for exclusion and data already collected will be recorded. Minimal information to be collected on screen failure includes demographic information, screen failure details, eligibility criteria and any severe adverse events.

#### Second-tier eligibility screening (by investigators)

Second-tier eligibility screening will be conducted in person at the first scheduled visit. Participants under the age of 18 years must be accompanied by a parent or legally acceptable representative (LAR). Written consent and assent will be obtained prior to second-tier screening. Communication will be in the language preferred by the participants.

Screening questions

1. confirm printed information including name, date of birth, sex, school, grade, race, language preferred, and contact information
2. able to adhere to follow-up schedules for the coming 3 years
3. willing to report reactions daily 7 days post-doses
4. no known history of COVID-19
5. past receipt of coronavirus vaccines
6. history of severe allergy, such as angioedema, bronchospasm and/or hypotension to food, drug, vaccine, or unknown trigger
7. history of Steven-Johnson syndrome (SJS), toxic epidermal necrolysis (TEN), drug rash with eosinophilia and systemic syndrome (DRESS)/drug-induced hypersensitivity syndrome (DIHS)
8. history of severe neurological conditions, e.g. transverse myelitis, Guillain-Barre Syndrome, demyelinating diseases
9. any significant medical disease that require regular medication or care
10. severe immunocompromise, including present or planned use of cytotoxic agents or systemic immunosuppressives, known primary or secondary immunodeficiency diseases
11. receipt of blood or plasma products or immunoglobulin within 60 days prior to vaccination to conclusion of the study
12. any haematological diseases (that preclude intramuscular injection)
13. reported pregnancy or breastfeeding
14. any acute illness or symptoms within 48 hours before vaccination that may suggest a potential COVID-19 illness, including new or increased respiratory symptoms, chills, new or increased myalgia, new loss of smell or taste, new gastrointestinal distress, and any other signs as judged by investigators
15. short-term systemic steroids use (<14 days) for an acute condition precludes the participant from receiving study vaccines until 28 days after the termination of said therapy

Physical examination

1. Height and weight
2. Temperature
3. General examination
4. Targeted organ system examination as directed by medical history

Screen failures, reason for exclusion and data already collected will be recorded. Minimal information to be collected on screen failure includes demographic information, screen failure details, eligibility criteria and any severe adverse events.

#### Pre-dose 2/3 screening

1. confirm adverse reactions and adverse events reported post-dose 1/2
2. any new arising medical history, pregnancy/breastfeeding or medication/vaccine use
3. any acute illness or symptoms within 48 hours before vaccination that may suggest a potential COVID-19 illness, including new or increased respiratory symptoms, chills, new or increased myalgia, new loss of smell or taste, new gastrointestinal distress, and any other signs as judged by investigators
4. short-term systemic steroids use (<14 days) for an acute condition precludes the participant from receiving study vaccines until 28 days after the termination of said therapy
5. temperature
6. targeted organ system examination as directed by medical history

Screen failures, reason for exclusion and data already collected will be recorded. Minimal information to be collected on screen failure includes demographic information, screen failure details, eligibility criteria and any severe adverse events.

#### Pre-existing history assessments

Previous medical history, including chronic illnesses, allergy history, and prior COVID-19 will be documented.

For chronic illnesses and allergies, the type of conditions, stability and disease course and concomitant medications will be recorded. For patients with inborn errors of immunity or other paediatric conditions, their latest medical charts will also be retrieved and studied.

For patients with prior COVID, we capture the type, severity and duration of signs and symptoms

1. Method and results of microbiological diagnosis, and evidence of reinfection if applicable
2. Results of clinical laboratory tests
3. Clinical history documentations
4. Concomitant care and intervention received
5. Duration of hospitalization and outcome

Such information will also be captured when there are persistent clinical abnormalities after the resolution of the acute episode. Investigators will then determine the overall clinical severity of each COVID-19 episode, according to the following criteria.

1. Asymptomatic: patient free of symptoms throughout the episode
2. Mild/Moderate: patient symptomatic, but never admitted to intensive care
3. Severe: patient admitted to intensive care unit due to clinical deterioration during the episode

After resolution of the initial acute COVID-19 episode, the following information will be solicited from participants monthly until the resolution of all symptoms from the episode, or as volunteered by the participants. They include

1. Persistence of clinical or laboratory abnormalities and related clinical details, suggestive of long COVID-19
2. Recurrence of symptoms and diagnosis of reinfection and related clinical details
3. Onset of multisystem inflammatory syndrome in children or adults, as diagnosed according to latest accepted guidelines available at the time, and related clinical details

#### Reactogenicity assessments

Adverse reactions will be solicited from participants on the day and 6 days after each dose on a web-based electronic diary card, with daily notifications/messages sent to participants. Participants are requested to fill out the electronic diary card at the same time of the day each day, preferably before bed.

List of adverse reactions and related information to submit include:

1. Pain at injection site 注射部位疼痛
2. Pruritus at injection site 注射部位瘙癢
3. Swelling, erythema and induration at injection site: diameter (cm) and photo 注射部位腫脹，瘙癢，紅斑和硬結：直徑（cm）和照片
4. Headache 頭痛
5. Fatigue 疲勞
6. Myalgia 肌肉痛
7. Nausea 噁心
8. Diarrhea 腹瀉
9. Vomiting 嘔吐
10. Arthralgia 關節痛
11. Cough 咳嗽
12. Chills 寒意/發冷
13. Fever: temperature (degrees Celsius) 發燒
14. Decreased appetite 食慾不振
15. Rhinorrhea 漏鼻涕
16. Nasal congestion 鼻塞
17. Sore throat 喉痛
18. Abdominal pain 腹痛

For listed reactions that are present, participants have to fill in related information as specified for the reaction, or determine their severity according to the following scale:

1. Mild: tolerable, not affecting daily activities
2. Moderate: performance of some daily activities affected
3. Severe: performance of some daily activities prevented

Such submitted information will be reviewed by trial staff prior to the next visit. Erratic inputs may be clarified by trial staff over phone calls to participants. Investigators will review the submitted inputs with participants at the next visit.

The following primary outcome can be assessed.

- 1a. Percentage of types and severity of adverse reactions occurring within 7 days post-doses 1 and 2 (and 3)

- 2a. Difference in percentages of types and severity of adverse reactions occurring within 7 days post-doses 1 and 2 (and 3)

#### Safety assessments (adverse events, COVID-19 and pregnancy)

Adverse events

Adverse events other than listed adverse reactions 28 days (or until their next dose, whichever is earlier) will be solicited from participants before the next visit. Their severity (on scale 1-3), duration and concomitant care and interventions will be documented. Those requiring medical attendance will be classified as medically attended adverse events. Their relevance to the study vaccines will be determined by investigators at the next visit, and confirmed by the study endpoint committee. Adverse events do not include those related to COVID-19 infection or sequelae. In the subgroup CE with inborn errors of immunity, immunological or hematological conditions, or other paediatric conditions, clinical follow-up visits may be scheduled with their respective clinicians within 28 days after each dose. Any new or worsened clinical or laboratory abnormalities, as tested by their clinicians, will also be solicited.

Severe adverse events

Severe adverse events include those adverse events involving any of the following conditions.

1. Hospitalization
2. Life-threatening
3. Death
4. Disability or permanent damage, including birth defect of participants' offspring

Severe adverse events are solicited from participants monthly through the electronic diary system. Detailed medical history will be requested from the participants promptly once reported and recorded. Additional visits may be scheduled for participants experiencing a severe adverse event. Their relevance to the study vaccines will be determined by investigators at the next visit, and confirmed by the study endpoint committee. Severe adverse events do not include those related to new onset COVID-19 infection or sequelae.

Special event: COVID-19

If a participant becomes diagnosed with COVID-19 breakthrough infection, either flagged by investigators on the basis of positive serology not explained by vaccine and with further clinical confirmation, or reported by participants on electronic diary card or via other routes, the following information will be solicited by the investigators, and reported to the Steering Committee promptly.

6. Type, severity and duration of signs and symptoms
7. Method and results of microbiological diagnosis, and evidence of reinfection if applicable
8. Results of clinical laboratory tests
9. Clinical history documentations
10. Concomitant care and intervention received
11. Duration of hospitalization and outcome

Such information will also be captured when there are persistent clinical abnormalities after the resolution of the acute episode. Investigators will then determine the overall clinical severity of each COVID-19 episode, according to the following criteria.

4. Asymptomatic: patient free of symptoms throughout the episode
5. Mild/Moderate: patient symptomatic, but never admitted to intensive care

6. Severe: patient admitted to intensive care unit due to clinical deterioration during the episode
7. Death: patient died during the episode

After resolution of the initial acute COVID-19 episode, the following information will be solicited from participants monthly until the resolution of all symptoms from the episode, or as volunteered by the participants. They include

4. Persistence of clinical or laboratory abnormalities and related clinical details, suggestive of long COVID-19
5. Recurrence of symptoms and diagnosis of reinfection and related clinical details
6. Onset of multisystem inflammatory syndrome in children or adults, as diagnosed according to latest accepted guidelines available at the time, and related clinical details

Special event: pregnancy

In female participants, pregnancy and breastfeeding practices will be requested monthly. Pregnancy that arise after vaccination during the period of the study will be documented at each visit for the following:

1. Gestational age
2. Expected due date
3. Results of prenatal examinations
4. (After the conclusion of pregnancy) History of the delivery
5. (After the conclusion of pregnancy) Outcome of pregnancy

The following secondary outcomes will also be assessed.

- 5a. Percentage of types and severity of adverse events and severe adverse events throughout study period
- 10a. Incidence of COVID-19 in participants throughout study period as self-reported or as determined by Luciferase Immunoprecipitation Systems assay

#### Blood-based assessments (immunogenicity and other immune tests)

Sample collection

Blood samples will be collected at each visit, and prior to vaccination for the visits involving vaccination. The following volumes of blood will be drawn in the following tubes in this order. Total volume of blood will not exceed 25 ml for paediatric participants at each visit.

1. 6 ml in clotted blood (red) tube
2. 16 ml in lithium heparin non-gel (green) tubes
3. 1 ml in EDTA (lavender) tube

Total volume of blood will not exceed 35 ml for adult participants at each visit. Written and oral consent will be additionally obtained by the phlebotomist to obtain the higher volume of blood for an adult participant.

4. 6-12 ml in clotted blood (red) tube
5. 16-20 ml in lithium heparin non-gel (green) tubes

6. 1 ml in EDTA (lavender) tube

Participant identifier and English name will be documented on the blood tubes. Time of blood taking will be documented.

Blood samples will be stored at 4 degrees Celsius or room temperature at the site of bloodtaking, and transported to the laboratory within the same day for processing.

Assessments to perform
Protocols for laboratory assessments of blood-based samples are available in the appendices.

1. Serology: Enzyme-linked Immunosorbent Assay (ELISA) against SARS-CoV2 S and N proteins; plaque reduction microneutralization assays (PRNA) against SARS-CoV2 and related coronaviruses or variants; Luciferase Immunoprecipitation System (LIPS) assay against multiple SARS-CoV2 proteins; surrogate neutralization tests against SARS-CoV2 and related coronaviruses or variants
2. T cell: flow cytometric analysis of T cells responsive to SARS-CoV2 peptide pools stimulation
3. Whole blood stimulation: transcriptomic and cytokine analyses of whole blood samples stimulated by TLR agonists and SARS-CoV2 S protein

Outcomes assessed
The following primary outcomes will be assessed.

3a. Geometric mean OD450 value of SARS-CoV2 S and S RBD-specific binding antibody and related markers as determined by Enzyme-linked Immunosorbent Assay at 1 month post-dose 1, and 1, 6, 18, and 36 months post-vaccination (and 2 weeks post-dose 3)

3b. Geometric mean inhibition level of SARS-CoV2 surrogate virus neutralization test at 1 month post-dose 1, and 1, 6, 18, and 36 months post-vaccination (and 2 weeks post-dose 3)

3c. Geometric mean titre of SARS-CoV2 neutralizing antibodies as determined by plaque reduction neutralization assay at 1 month post-dose 1, and 1, 6, 18, and 36 months post-vaccination (and 2 weeks post-dose 3)

3d. Geometric mean frequency of Th1 and Tc1 cells specific to SARS-CoV2 S (and N and M for CC) protein at 1 month post-dose 1, and 1, 6, 18, and 36 months post-vaccination (and 2 weeks post-dose 3)

3e. Geometric mean OD450 value of SARS-CoV2 S-specific FcgRIIIa-binding antibody as determined by Enzyme-linked Immunosorbent Assay at 1 month post-dose 1, and 1, 6, 18, and 36 months post-vaccination (and 2 weeks post-dose 3)

3f. Geometric mean OD450 value of SARS-CoV2 S-specific binding antibody avidity index as determined by Enzyme-linked Immunosorbent Assay at 1 month post-dose 1, and 1, 6, 18, and 36 months post-vaccination (and 2 weeks post-dose 3)

3g. Geometric mean frequency of SARS-CoV2 S -specific B cells as determined by flow cytometry at 1 month post-dose 1, and 1, 6, 18, and 36 months post-vaccination (and 2 weeks post-dose 3)

4a. Geometric mean ratios of primary immunogenicity outcomes between adults and children

4b. Geometric mean ratios of primary immunogenicity outcomes between vaccines

The following secondary outcomes will be assessed.

6a. Mean count and percentage of white blood cell subtypes in peripheral blood of participants at time of dose 1

6b. Whole blood cytokine and transcriptomic profiles upon ex vivo stimulation by TLR3, 7-9 ligands of participants at time of dose 1

6c. Nasopharyngeal metagenomic, cytokine, immunoglobulin, IgG allotypes and other proteogenomic profiles at time of dose 1

6d. Demographic, anthropometric and other baseline characteristics of participants

7a. Geometric mean OD450 value of SARS-CoV2 N (and N-CTD)-specific binding antibody as determined by Enzyme-linked Immunosorbent Assay at 1 month post-dose 1, and 1, 6, 18, and 36 months post-vaccination (and 2 weeks post-dose 3)

7b. Geometric mean OD450 value of SARS-CoV2 N-specific FcgRIIIa-binding antibody as determined by Enzyme-linked Immunosorbent Assay at 1 month post-dose 1, and 1, 6, 18, and 36 months post-vaccination (and 2 weeks post-dose 3)

7c. Geometric mean OD450 value of SARS-CoV2 N-specific binding antibody avidity index as determined by Enzyme-linked Immunosorbent Assay at 1 month post-dose 1, and 1, 6, 18, and 36 months post-vaccination (and 2 weeks post-dose 3)

7d. Geometric mean ratios of immunogenicity outcomes between children and adults

8a. Geometric mean frequency of Th2 and Tc2 cells specific to SARS-CoV2 peptide pools at 1 month post-dose 1, and 1, 6, 18, and 36 months post-vaccination (and 2 weeks post-dose 3)

8b. Geometric mean ratio of Th1/Tc1 and Th2/Tc2 cells specific to SARS-CoV2 peptide pools at 1 month post-dose 1, and 1, 6, 18, and 36 months post-vaccination(and 2 weeks post-dose 3)

8c. Geometric mean ratios of such counts/ratios in adults versus adolescent vaccinees of same vaccine at various timepoints

8d. Geometric mean ratios of such counts/ratios in vaccinees of different vaccine

9a. Geometric mean OD450 value of SARS-CoV2 variant S RBD-specific binding antibody and related markers as determined by Enzyme-linked Immunosorbent Assay at 1 month post-dose 1, and 1, 6, 18, and 36 months post-vaccination (and 2 weeks post-dose 3)

9b. Geometric mean inhibition level of SARS-CoV2 surrogate variant virus neutralization test at 1 month post-dose 1, and 1, 6, 18, and 36 months post-vaccination (and 2 weeks post-dose 3)

9c. Geometric mean titre of SARS-CoV2 neutralizing antibodies as determined by variant plaque reduction neutralization assay at 1 month post-dose 1, and 1, 6, 18, and 36 months post-vaccination (and 2 weeks post-dose 3)

9d. Geometric mean frequency of Th1 and Tc1 cells specific to SARS-CoV2 variant S protein and other peptide pools at 1 month post-dose 1, and 1, 6, 18, and 36 months post-vaccination (and 2 weeks post-dose 3)

10a. Incidence of COVID-19 in participants throughout study period as self-reported or as determined by Luciferase Immunoprecipitation Systems assay

12a. Geometric mean ratios of immunogenicity outcomes between paediatric patients and healthy children

14a. Whole blood cytokine and transcriptomic profiles upon ex vivo stimulation by SARS-CoV2 S protein of participants at 1 month post-dose 2 (and 2 weeks post-dose 3)

Geometric mean ratio of immunogenicity outcomes between intradermal and intramusuclar CoronaVac immunization

#### Nasopharyngeal assessments

Sample collection
A nasopharyngeal swab will be taken in consenting participants at visits 1 and 3 and after. Refusal to participate in nasopharyngeal assessment is not an exclusion criterion. Written and oral consent will be taken from participants prior to sampling.

This procedure will be performed by healthcare professionals in full personal protective equipment experienced in nasopharyngeal swabs in a well ventilated room that is sterilized after use.

Assessments to perform
Nasopharyngeal metagenomic, cytokine, immunoglobulin and other proteogenomic analyses

Outcomes assessed
The following secondary outcome will be assessed.

- 15a. Nasopharyngeal immunoglobulin and other immune responses 1 month and 6 months post-vaccination (and 2 weeks post-dose 3)

#### Psychosocial assessments

Assessments to perform
A psychosocial wellbeing questionnaire

Outcomes assessed
The following secondary outcome will be assessed.

- 11a. Psychosocial assessments at time of dose 1 and 6, 18 and 36 months post-vaccination

#### Assessments specific to long COVID

##### Assessments to perform

Assessments include a longitudinal patient survey, and clinical and laboratory testing.

- A longitudinal patient survey filled in at dose 1, and 2 weeks, 1, 3, 6, 18 and 36 months after dose 1
- A basic laboratory screen including complete blood count with white cell differential, liver and renal function tests, glucose/lipid profile and electrolytes level at or before dose 1, and 1, 6, 18 and 36 months after dose 1/2

The longitudinal patient survey will include an assessment on the types, duration and severity of self-perceived symptoms of patients with prior COVID-19, including:

1. Anxiety/depression
2. Sleep disturbance
3. PTSD
4. Delirium
5. Brain fog
6. Hair loss
7. Skin rash
8. Skipped meals
9. Abdominal pain
10. Diarrhea
11. Chest pain
12. Muscle pains
13. Joint pain
14. Fever
15. Sore throat
16. Persistent cough
17. Hoarse voice
18. Loss of taste/smell
19. Shortness of breath
20. Headache
21. Fatigue
22. Cough
23. Palpitations
24. Others

Dates of onset and resolution, weekly occurrence and severity will be captured on the electronic survey. Use of concomitant medications will also be asked.

##### Outcomes assessed

The following secondary measures will be assessed.

- 13a. Percentage of types, duration and severity of long COVID-19 abnormalities at time of dose 1 and 1, 6, 18 and 36 months post vaccination

##### *18b Plans to promote participant retention and complete follow-up, including list of any outcome data to be collected for participants who discontinue or deviate from intervention protocols*

###### Participant incentive

100 HKD will be given to each participant at each visit. Disbursement and receipt of the incentive will be documented.

###### Failure to report reactions on electronic diary 7 days post-vaccination

A message will be sent to all participants to remind them to fill out the electronic diary and report reactions daily. A second message will be sent to participants who failed to fill out the electronic diary for one day the next day. Participants who fail to fill out the electronic diary for 2 days will be called by trial staff for clarification, and delayed/non-reporting, contact and reason will be documented.

###### Absence at scheduled visits

Every effort will be made to contact participants who do not turn up at a scheduled time, and visits may be rescheduled within the window for each visit whenever possible. Absence, contact and reason for absence will be documented.

Participants absent at one scheduled visit and unreachable by 3 phone calls and a letter to their last known address within 2 weeks after their unattended scheduled visit will be considered lost to follow-up and withdrawn from the study.

###### Data collection from participants who discontinue or deviate from intervention protocol or the study

Data collection will be continued regardless of any deviation from intervention protocol on any participants who have received at least one dose of any vaccines. Data and samples already collected from withdrawn or discontinued participants will be retained and analysed. The analysis of trial data will be performed by modified intention-to-treat and per-protocol analysis as specified in statistical analysis plan. Missing data will be managed as mentioned in 20c.

###### Discontinuation or withdrawal of participants from the study

Participants may withdraw from the study at any time at their request. Reasons for withdrawal by participants or discontinuation of participants by investigators will be documented.

### Data management

*19 Plans for data entry, coding, security, and storage, including any related processes to promote data quality (eg, double data entry; range checks for data values). Reference to where details of data management procedures can be found, if not in the protocol*

##### Data entry

Single data entry will be performed and cross-checked by trial staff. Range checks will be performed for data values.

##### Data storage

Hard copies of any raw data and samples collected or generated in the performance of the trial will be securely retained for a period of 15 years in the University of Hong Kong or overseas collaborators. Only pseudonymized samples and no personal identification will be shared with overseas collaborators. Soft copies of any data collected or generated in the performance of the trial will be retained for a period of 15 years in secure backed-up hard drives. Any raw data and samples collected or generated in the study period will be destroyed 15 years after the conclusion of the study.

##### Data security

All physical data will be stored in locked cabinets, temporarily in the locations for the trial and in the Department of Paediatrics and Adolescent Medicine and the School of Public Health, The University of Hong Kong for long term storage. Digital data will be stored in password-protected encrypted databases accessible only to trial staff and investigators.

### Statistical methods

*20a Statistical methods for analysing primary and secondary outcomes. Reference to where other details of the statistical analysis plan can be found, if not in the protocol*

Detailed statistical methods applied at each analysis can be found in the statistical analysis plan.

*20b Methods for any additional analyses (eg, subgroup and adjusted analyses)*

Detailed statistical methods applied at each analysis can be found in the statistical analysis plan

*20c Definition of analysis population relating to protocol non-adherence (eg, as randomised analysis), and any statistical methods to handle missing data (eg, multiple imputation)*

Detailed statistical methods applied at each analysis can be found in the statistical analysis plan

### Methods: Monitoring

#### Data monitoring

*21a Composition of data monitoring committee (DMC); summary of its role and reporting structure; statement of whether it is independent from the sponsor and competing interests; and reference to where further details about its charter can be found, if not in the protocol. Alternatively, an explanation of why a DMC is not needed*

3 groups will be responsible for data monitoring.

- Steering committee of the trial
- Institutional Review Board approving this study, i.e. IRB of HKU/HA HKWC
- Regulatory authority approving this study, i.e. Department of Health, HKSARG

As required by the Institutional Review Board and Department of Health, severe adverse events will be reported promptly according to the mandated timelines.

A data monitoring committee independent of the investigators is not considered necessary as the investigators declare no competing interests with the manufacturers or inventors of the vaccine candidates trialed.

*21b Description of any interim analyses and stopping guidelines, including who will have access to these interim results and make the final decision to terminate the trial*

##### Interim analysis

Interim analysis of primary outcomes permitted by the timeline will be carried out after half to all of the participants for that study arm have reached each timepoint. Interim analysis may also be conducted when a severe adverse event or a severe COVID-19 infection that may be relevant to study vaccines is flagged.

Adverse reactions, adverse events, pregnancy and COVID-19 in participants will be subject to ongoing contemporaneous review.

##### Stopping a study arm

Stopping rule for a study arm include any of the following:

1. A participant vaccinated develops a severe adverse event/reaction that is possibly related to the intervention, or for which there are no plausible alternative explanation
2. A participant vaccinated develops COVID-19 infection that necessitates intensive care or fatal COVID-19 infection, that is plausibly a result of vaccine-enhanced disease or prophylactic futility of the study vaccine, or for which there are no plausible alternative explanation

When an event that may cause either rule to be met is flagged by an investigator, the Steering Committee will review all available evidence related to the event, and all data collected from the affected participant. The Steering Committee will then confirm if a stopping rule is met, and order the pause temporarily or halt the study arm.

During the pause or halt, only recruitment of new participants and further intervention will be stopped for that arm. Other aspects of the trial including follow-up, blood taking, reactogenicity and safety reporting will continue throughout the study period. Activities of other study arms will remain unaffected.

The study arm may be resumed at the discretion of the Steering Committee, given the level of risk for other participants is anticipated to remain low.

##### Stopping the entire study

The entire study may be paused temporarily or halted when more than one study arm is stopped or halted, or for other practical reasons that prevent the continuation of the study. Other aspects of the trial including follow-up, blood taking, reactogenicity and safety reporting may continue throughout the study period, at the judgement of the Steering Committee.

#### Harms

*22 Plans for collecting, assessing, reporting, and managing solicited and spontaneously reported adverse events and other unintended effects of trial interventions or trial conduct*

##### Reaction to study article

Adverse reactions will be solicited and managed as described in previous sections.

##### Adverse events

Adverse events will be solicited and managed as described in previous sections.

##### Severe adverse events

Severe adverse events will be solicited and managed as described in previous sections.

#### Auditing

*23 Frequency and procedures for auditing trial conduct, if any, and whether the process will be independent from investigators and the sponsor*

##### Responsibility for study auditing

Study audit will be conducted by the Steering Committee monthly to review the conduct of the study, and evaluate any deviations from the study protocols reported that may jeopardize the safety and privacy of participants, and scientific results of the study. Study audits will also be conducted by the Institutional Review Board and regulatory authorities approving this study if needed.

##### Deviation from study protocol by investigators

Medication errors
Medication errors will be documented as adverse events. Examples of medication errors include:

- Administration of vaccines exposed to incorrect handling practices that may affect the safety of participants, e.g. contaminated, expired or stored outside recommended temperatures
- Administration of incorrect vaccines
- Administration of incorrect dosage

Participants administered a potentially harmful vaccine or dosage will be closely monitored and followed up by trial staff for any adverse events. Medication errors will be promptly reported to the Steering Committee within the same day.

##### Assessment errors

Assessment errors need not be documented as adverse events, unless those that exposed participants to harm. Adverse events related to assessment errors should be deemed unrelated to study interventions. Examples of assessment errors that should be documented as adverse events and reported to the Steering Committee within the same day include:

- Injury to participants during blood-taking or taking of nasopharyngeal swab
- Exposure of participants to potential harm or injury during visits

Assessment errors that do not expose participants to harm should be reported to investigators in charge of the assessment promptly and recorded for later review by the Steering Committee. Examples include:

- Improper handling of samples that disqualify samples from being assessed
- Improper performance of testing procedures that disqualify the results of the test

##### Data handling errors

Data handling errors or other potential data breaches discovered by the Data Management Committee should be promptly reported to and investigated by the Steering Committee, and appropriate bodies including affected participants and regulatory authorities should be notified as soon as possible.

### Ethics and dissemination

#### Research ethics approval

##### *24 Plans for seeking research ethics committee/institutional review board (REC/IRB) approval*

Ethics approval is pending at the Institutional Review Board of The University of Hong Kong/Hospital Authority Hong Kong West Cluster. Study procedures beginning with the eligibility screen of the potential study participants will start after ethics approval.

This study will be performed as approved by the IRB, universal ethical principles such as the Declaration of Helsinki, applicable ICH GCP guidelines, and applicable laws and institutional regulations.

#### Protocol amendments

##### *25 Plans for communicating important protocol modifications (eg, changes to eligibility criteria, outcomes, analyses) to relevant parties (eg, investigators, REC/IRBs, trial participants, trial registries, journals, regulators)*

###### Communication with investigators

Important protocol modifications including the updated protocol and summary of changes will be promptly communicated to all investigators.

###### Communication with Institutional Review Board

Approval of important protocol modifications proposed including the updated protocol and summary of changes will be requested to the IRB before implementation.

###### Communication with trial participants

Important protocol modifications that affect the conduct of trial participants will be communicated to participants in a timely manner.

###### Communication with trial registry

Important protocol modifications including the updated protocol and summary of changes will be promptly communicated to the trial registry.

###### Communication with journals

Journals will be provided with the updated protocol and summary of changes at time of manuscript submission.

##### Communication with the Department of Health, HKSARG

Approval of important protocol modifications proposed including the updated protocol and summary of changes will be requested to the regulatory body for clinical trials before implementation as required by law.

#### Consent or assent

*26a Who will obtain informed consent or assent from potential trial participants or authorised surrogates, and how (see Item 32)*

##### Consent

Trial staff will explain the study to potential participants in the language preferred by the participants. Information required to be communicated to the participants will be included in the information sheet. Trial staff and investigators should attempt to answer all queries from potential participants, and given enough time to consider their participation. An interpreter will be provided when needed.

Consent will be obtained from competent adults aged 18 years or above in written format. Consent for an underage participant will be obtained from a parent or other legally accepted representative of the child.

##### Assent

Assent will be obtained from participants aged 7-17 in written format, confirming they have a basic understanding of the study.

*26b Additional consent provisions for collection and use of participant data and biological specimens in ancillary studies, if applicable*

Not applicable.

#### Confidentiality

*27 How personal information about potential and enrolled participants will be collected, shared, and maintained in order to protect confidentiality before, during, and after the trial*

Investigators will comply with all applicable laws and institutional regulations related to data protection before, during and after the trial. Data management procedures as detailed elsewhere in this protocol will be followed. Data breaches will be promptly evaluated by the Steering Committee as detailed elsewhere in this protocol, and will be reported as required by law.

#### Declaration of interests

*28 Financial and other competing interests for principal investigators for the overall trial and each study site*

Principal investigator Yu Lung Lau declares the following interests: received funding on the vaccine and infectious diseases-related projects from vaccine industry, including GlaxoSmithKline (sponsored vaccine trials from 2004 to 2021), Merck, Sharp & Dohme (trials and an epidemiological study between 1999-2004), Wyeth (vaccine trial in 2002), and Medimmune Inc. (vaccine trial in 2004).

Other investigators declare no competing interests.

#### Access to data

*29 Statement of who will have access to the final trial dataset, and disclosure of contractual agreements that limit such access for investigators*

Final participant-level trial dataset will only be available to members of the trial's Steering Committee. Disclosure of the pseudonymized participant-level trial dataset may be requested to the Institutional Review Board of HKU/HKWC.

Contractual agreements to limit such access for investigators are not applicable.

#### Ancillary and post-trial care

*30 Provisions, if any, for ancillary and post-trial care, and for compensation to those who suffer harm from trial participation*

There are no provisions for ancillary or post-trial care. Compensation for injuries related to vaccination may be provided by the HKSAR Government outside of this trial, where applicable.

#### Dissemination policy

*31a Plans for investigators and sponsor to communicate trial results to participants, healthcare professionals, the public, and other relevant groups (eg, via publication, reporting in results databases, or other data sharing arrangements), including any publication restrictions*

##### Early disclosure

Trial results may be made public in preprint servers and via mass media whenever appropriate. Participant-level data will not be disclosed.

##### Publishing in academic journals

Trial results may be published in peer-reviewed academic journals, and reported according to EQUATOR Network guidelines where appropriate. Participant-level data will not be disclosed in academic publications. Authorship will be designated in accordance with the International Committee of Medical Journal Editors guidance.

##### Disclosure to ethics committee, regulatory bodies and trial registry

Trial results will be communicated to IRB, regulatory bodies and trial registry as required.

#### *31b Authorship eligibility guidelines and any intended use of professional writers*

##### Eligibility

Authorship will be designated following the International Committee of Medical Journal Editors guidance.

##### Professional writers

Not applicable.

#### *31c Plans, if any, for granting public access to the full protocol, participant-level dataset, and statistical code*

##### Full protocol

Full protocol will be released with journal articles arising from this study, excluding any confidential information.

##### Participant level dataset

Pseudonymized participant level dataset may be released upon request to the IRB of HKU/HKWC, after the conclusion of the study and publication of trial results.

##### Statistical code

Statistical code, where applicable, will be released with journal articles arising from this study.

End of protocol.
