## Supplementary material for "Superior antibody and membrane protein-specific T cell responses to CoronaVac by intradermal versus intramuscular routes in adolescents": Statistical Analysis Plan

**Study title: To compare the reactogenicity and immunogenicity of the recommended COVID-19 vaccines in young adolescents and children in Hong Kong**

| | |
|---|---|
| Protocol Identifier | COVAC01 |
| SAP Version No. | 2.0 |
| Date | April 1, 2022 |

COVID-19 Vaccination in Adolescents and Children
兒童及青少年新冠病毒疫苗研究

HKU Med LKS Faculty of Medicine
Department of Paediatrics & Adolescent Medicine
香港大學兒童及青少年科學系

C2i Centre for Immunology & Infection
免疫與感染研究中心

### Table of Contents


Table of Contents 2
1 Version History 5
2 Introduction 6
2.1 Study Objectives 6
2.2 Study Design 7
2.2.1 Healthy adolescents 7
2.2.3 Patients living with immunodeficiency or other paediatric diseases 7
3 Endpoints and baseline variables 8
3.1 Primary endpoints 8
3.1.1 Reactogenicity endpoints 8
3.1.1.1 Adverse reactions 8
3.1.2 Immunogenicity endpoints 9
3.1.2.1 anti-S IgG levels 9
3.1.2.2 Surrogate neutralizing antibody inhibition level 9
3.1.2.3 Neutralizing titers (by plaque reduction neutralization assay) 9
3.1.2.4 IgG avidity 10
3.1.2.5 IgG Fc receptor binding antibody 10
3.1.2.6 Th1 and Tc1 response 10
3.2 Secondary endpoints 11
3.2.1 Adverse events 11
3.2.2 Vaccine efficacy prediction 11
3.2.3 anti-N and N-CTD IgG levels 11
3.2.4 Correlates of immunogenicity 11
3.3 Participant characteristics 12
4 Analysis sets 13
4.1 Total enrolled population 13
4.2 Immunogenicity populations 13
4.3 Reactogenicity and safety populations 15
5 Methodology and hypotheses 16
5.1 Hypotheses and decision rules 16
5.1.1 Immunogenicity hypothesis 16

5.1.2 Sample size 17
5.1.3 Multiplicity consideration 17
5.2 General methods 18
5.2.1 Categorical variables 18
5.2.2 Continuous variables 18
5.2.2.1 Geometric means 18
5.2.2.2 Geometric mean fold rises/changes 18
5.2.2.3 Geometric mean ratios 18
5.2.2.4 Geometric mean fold rise ratios 18
6 Analysis and summary 19
6.1 Primary analysis 19
6.1.1 Reactogenicity endpoints 19
6.1.1.1 Adverse reactions 19
6.1.2 Immunogenicity endpoints 20
6.1.2.1 S and S-RBD binding IgG levels 20
6.1.2.2 Surrogate neutralizing antibody inhibition levels 20
6.1.2.3 Neutralizing antibody titers 20
6.1.2.4 S binding IgG avidity index 21
6.1.2.5 Anti-S FcgRIIIa-binding IgG levels 21
6.1.2.6 S (and N and M)-responsive Th1 and Tc1 cell frequencies 21
6.1.2.7 GMR of S and S-RBD binding IgG, surrogate neutralizing antibody inhibition level, neutralizing antibody titers, IgG avidity index and FcgRIIIa-binding IgG in participants 11-17 years of age to those 18 years of age or above 21
6.1.2.8 GMR of S (and N and M)-responsive Th1 and Tc1 cell frequencies in participants 11-17 years of age to those 18 years of age or above 22
6.1.2.9 GMR of S and S-RBD binding IgG, surrogate neutralizing antibody inhibition level, and neutralizing antibody titers, IgG avidity index and FcgRIIIa-binding IgG in participants 11-17 years of age receiving 2 doses of BNT162b2 to those receiving 2/3 doses of CoronaVac 22
6.1.2.10 GMR of total S responsive Th1 and Tc1 cell frequencies in participants 11-17 years of age receiving 2 doses of BNT162b2 to those receiving 2/3 doses of CoronaVac 22
6.1.2.11 GMR of S and S-RBD binding IgG, surrogate neutralizing antibody inhibition level, and neutralizing antibody titers, IgG avidity index and FcgRIIIa-binding IgG in participants receiving intradermal CoronaVac to intramuscular CoronaVac 23

6.1.2.12 GMR of total S, N and M responsive Th1 and Tc1 cell frequencies in participants 11-17 years of age receiving intradermal CoronaVac to intramuscular CoronaVac 23
6.2 Secondary endpoints 23
6.2.1 Adverse events 23
6.2.2 Vaccine efficacy prediction 24
6.2.3 N and N-CTD binding IgG levels 24
6.2.4 GMR of N and N-CTD binding IgG in participants 11-17 years of age to those 18 years of age or above receiving CoronaVac 24
6.2.5 Correlates of immunogenicity 25
6.2.6 GMR of S and S-RBD binding IgG, surrogate neutralizing antibody inhibition level, and neutralizing antibody titers, IgG avidity index and FcgRIIIa-binding IgG in participants 11 years or above of age living with diseases to those healthy receiving a particular vaccine 25
6.2.7 GMR of responsive Th1 and Tc1 cell frequencies in participants 11 years or above of age living with diseases to those healthy receiving a particular vaccine 25
6.3 Subgroup analysis 26
6.4 Baseline and other summaries and analyses 26
6.4.1 Baseline 26
6.4.1.1 Demographics 26
6.4.1.2 Health characteristics 26
6.4.2 Study completion and participant disposition 27
6.4.2.1 Participant disposition summary 27
6.4.2.2 Blood sampling and e-Diary completion 27
6.4.3 Intervention 27
6.4.3.1 Vaccination timing and administration 27
6.4.4 Prior/concomitant vaccination and medications 27
7 Analysis timeline 27
7.1 Interim analysis 1 (Nov 2021) 27
7.2 Interim analysis 2 (Jan 2022) 28
7.3 Interim analysis 3 (May 2022) 28
7.4 Interim analysis 4 (May 2022) 28

### 1 Version History

This is the second version of the statistical analysis plan for study protocol COVAC01, adding the details of the pre-specified preliminary analysis for healthy adolescent/child participants and adults (subgroup CA) 1 month after the second or third doses of BioNTech's BNT162b2 and SinoVac's CoronaVac. Longitudinal immunogenicity analysis describing the decay after vaccination over three years will also be specified for various timepoints. Patients with immune disorders or other pediatric conditions will also be studied and their immunogenicity outcomes compared against healthy counterparts.

### 2 Introduction

This is the second version of the statistical analysis plan for study protocol COVAC01, adding the details of the pre-specified preliminary analysis for healthy adolescent/child participants and adults (subgroup CA) 1 month after the second or third doses of BioNTech's BNT162b2 and SinoVac's CoronaVac. Longitudinal immunogenicity analysis describing the decay after vaccination over three years will also be specified for various timepoints. Patients with immune disorders or other pediatric conditions will also be studied and their immunogenicity outcomes compared against healthy counterparts. Interim analyses are performed when all participants in a particular subgroup have completed assessments at each timepoint.

#### 2.1 Study Objectives

*Primary objectives*

1. Report the frequencies of reactogenicity within the 7 days after each vaccine injection;
2. Compare frequencies of reactions among the vaccines;
3. Report the short-term and long-term (up to 36 months post-vaccination) immunogenicity outcomes including spike protein IgG and related markers, neutralizing antibody titres and B and T cell immune responses;
4. Compare immunogenicity outcomes between children and adults, such that the geometric mean values of immunological parameters are statistically non-inferior (0.6 of adult value) in children, and differences between vaccines in children

*Secondary objectives*

5. Describe adverse events and serious adverse events during the study period;
6. Explore any parameters or markers predictive and correlative of immune responses to vaccination;
7. Short-term and long-term (up to 36 months post-vaccination) longitudinal differences in non-spike protein antibody titres for participants receiving CoronaVac;
8. Short-term and long-term (up to 36 months post-vaccination) longitudinal differences in type I and II T cell immune responses between vaccines and between children and adults
9. Differences in short-term and long-term (up to 36 months post-vaccination) longitudinal differences in spike protein IgG and related markers, neutralizing antibody titres and cellular immune responses against emerging viral variants between vaccines, and between children and adults;
10. Vaccine breakthroughs as documented by N and ORF8 antibodies in subjects receiving BNT162b2, or ORF8 in subjects receiving CoronaVac, with estimation of potential immunological correlates of protection against infection.
11. Explore the psychosocial impact of COVID-19 vaccination in adolescents
12. Impact of pre-existing conditions on safety, reactogenicity and immunogenicity of vaccines
13. Impact of vaccination on adolescents with previous COVID-19 and long COVID

14. Short-term and long-term (up to 36 months post-vaccination) longitudinal differences in SARS-CoV2 whole blood immune responses between vaccines and between children and adults
15. Short-term and long-term (up to 36 months post-vaccination) longitudinal differences in SARS-CoV2 nasopharyngeal immune responses between vaccines and between children and adults
16. Predict vaccine efficacy based on established immune correlates of protection

#### 2.2 Study Design

##### 2.2.1 Healthy adolescents

This is a single-center, immunobridging study in 200 healthy adolescents aged 11-17 years and their parents or other adult controls, in which participants elect to be immunized against COVID-19 with BioNTech BNT162b2 or SinoVac's CoronaVac. A variety of immunogenicity outcomes will be measured and bridged to their healthy parents to demonstrate respective non-inferiority of the vaccines in adolescents to adults. Reactogenicity and safety will also be described. Participants will be followed-up for a period of 3 years.

##### 2.2.3 Patients living with immunodeficiency or other paediatric diseases

Adolescents living with immunodeficiencies or other paediatric diseases will be recruited in disease subgroups. Safety, reactogenicity and immunogenicity will be described as healthy adolescents. Two or three doses of the vaccines will be given as the trialled primary series. Immunogenicity outcomes will be compared with those of healthy adolescents over 3 years.

### 3 Endpoints and baseline variables

#### 3.1 Primary endpoints

##### 3.1.1 Reactogenicity endpoints

###### 3.1.1.1 Adverse reactions

For all participants receiving one dose or more under our study protocol, excluding those who already completed immunization prior to joining the study, we solicit prespecified local and systemic reactions for 7 days after immunization with each dose (days 0 to 7). Participants are asked to report these reactions daily and will be reminded if they missed reporting.

Solicited reactions are either local or systemic, and they include:

- Pain at injection site
- Pruritus at injection site
- Swelling, erythema and induration at injection site
- Headache
- Fatigue
- Myalgia
- Nausea
- Diarrhea
- Vomiting
- Arthralgia
- Cough
- Chills
- Fever
- Decreased appetite
- Rhinorrhea
- Nasal congestion
- Sore throat
- Abdominal pain

For listed reactions that are present, participants are to fill in related information as specified for the reaction (fever and swelling, erythema and induration at injection site), or determine the severity according to the following scale:

1. Mild: tolerable, not affecting daily activities
2. Moderate: performance of some daily activities affected
3. Severe: performance of some daily activities prevented

For fever, we determine the severity according to body temperature reported by participant:

1. Mild: 38.0-38.4 degrees Celsius
2. Moderate: 38.5-38.9 degrees Celsius
3. Severe: 39.0 degrees Celsius or higher

For swelling, erythema and induration at injection site, we determine the severity according to the following:

1. Mild: diameter 2.0 to 4.9 cm
2. Moderate: diameter 5.0 to 9.9 cm
3. Severe: diameter 10.0 cm or longer

We analyze the presence and absence of each adverse reaction by maximum severity and use of antipyretic medication on any day from days 0-7 after each dose.

- Presence on any day refers to reported presence on any day between days 0-7.
- Absence on any day refers to no reported presence on any day between days 0-7, either reported absence on all days or reported absence on any day and missing report on other days.
- Missing refers to a continued missing report on all days between days 0-7

##### 3.1.2 Immunogenicity endpoints

###### 3.1.2.1 anti-S IgG levels

We measure SARS-CoV2 full S and S-RBD binding antibody level in sera as a primary immunogenicity endpoint. We report results as OD450 value. Values below the limit of detection will be taken as 0.5 of the lower limit of detection. **Geometric** mean OD450 values will be calculated for each timepoint in each subgroup.

###### 3.1.2.2 Surrogate neutralizing antibody inhibition level

We measure SARS-CoV2 S-RBD antibody ACE2-binding inhibition level in sera as a primary immunogenicity endpoint. We report results as level of inhibition (%). Values below the limit of quantification will be taken as 0.5 of the lower limit of quantification. **Geometric mean** inhibition percentage will be calculated for each timepoint in each subgroup.

###### 3.1.2.3 Neutralizing titers (by plaque reduction neutralization assay)

We measure SARS-CoV2 50% and 90% neutralizing titers by plaque reduction neutralization assay (PRNT50) against a virus isolate obtained early in the pandemic (BetaCoV/Hong Kong/VM20001061/2020) as a primary immunogenicity endpoint. Values below the limit of detection will be taken as 0.5 of the lower limit of detection. Values higher than the upper limit of detection (highest dilution tested) will be imputed as the next theoretical dilution. Geometric mean titers will be calculated for each timepoint in each subgroup.

###### 3.1.2.4 IgG avidity

We measure SARS-CoV2 full S binding antibody avidity in sera as a primary immunogenicity endpoint. We report results as avidity index (%). **Geometric mean** percentages will be calculated for each timepoint in each subgroup.

###### 3.1.2.5 IgG Fc receptor binding antibody

We measure SARS-CoV2 full S FcgRIIIa-binding antibody concentration in sera as a primary immunogenicity endpoint. We report results as OD450 value. Values below the limit of detection will be taken as 0.5 of the lower limit of detection. **Geometric mean** OD450 values will be calculated for each timepoint in each subgroup.

###### 3.1.2.6 Th1 and Tc1 response

We measure the frequency of responsive Th1/Tc1 cells among all peripheral blood mononuclear cells (PBMC) in peripheral heparinized blood as a primary immunogenicity endpoint.

Cell types are defined with the following surface markers and intracellular cytokine staining on flow cytometry against stimulation by all SARS-CoV2 peptide pools for CoronaVac participants or only S peptide pool for BNT162b2 participants:

- Responsive Th1 cells are defined as CD3+ CD4+ IFN-g+ or IL-2+ cells in PBMC
- Responsive Tc1 cells are defined as CD3+ CD8+ IFN-g+ or IL-2+ cells in PBMC

The results will be reported as percentage of parent population: percentage of CD4 T cells or CD8 T cells, as applicable.

Geometric mean percentages will be calculated for each timepoint in each subgroup.

#### 3.2 Secondary endpoints

##### 3.2.1 Adverse events

For all participants receiving one dose or more under our study protocol, excluding those who already completed immunization prior to joining the study, we evaluate unsolicited, volunteered local and systemic adverse events for 28 days after immunization with each dose (days 0 to 28) before receipt of the next dose (at least 21 days for BNT162b2 first dose).

For all participants receiving one dose or more under our study protocol, including those who already completed immunization prior to joining the study, we evaluate unsolicited, volunteered severe adverse events from joining the study, until 36 months after completing immunization. Severe adverse events are defined as adverse events involving any of the following conditions.

1. Hospitalization
2. Life-threatening illness
3. Death
4. Disability or permanent damage, including birth defect of participants' offspring

##### 3.2.2 Vaccine efficacy prediction

We predict vaccine efficacy for recipients of each vaccine based on neutralizing antibody correlate of protection, as published by Khoury et al in Nature Medicine, 2021. Geometric mean neutralizing titers in participants receiving either vaccines will be normalized against that of a set of house convalescent controls as fold of convalescent titers, and protective efficacy against symptomatic disease will be predicted based on the published model.

##### 3.2.3 anti-N and N-CTD IgG levels

We measure SARS-CoV2 full N and N-CTD binding antibody levels in sera as a primary immunogenicity endpoint. We report results as OD450 value. Values below the limit of detection will be taken as 0.5 of the lower limit of detection. Geometric mean OD450 values will be calculated for each timepoint in each subgroup.

##### 3.2.4 Correlates of immunogenicity

Correlations between primary immunogenicity outcomes will be evaluated by Pearson correlation coefficients after natural log transformation, with a more stringent significance level with lower alpha to account for multiple comparison testing. Relationships between immunogenicity outcomes and baseline variables such as age, sex and haematological parameters will be explored by multiple linear regression by vaccine type and age subgroup.

#### 3.3 Participant characteristics

Demographic variables obtained include age at time of dose 1 (in years), sex (male or female), medical history (generally healthy, pre-existing COVID-19 or immunodeficient), pre-existing conditions (coded according to medDRA) and concomitant medications.

### 4 Analysis sets

#### 4.1 Total enrolled population

| Population | Description |
|---|---|
| Enrolled | All participants with a signed informed consent form, screened in as included by study physician and received at least one dose of study vaccine |

#### 4.2 Immunogenicity populations

| Population | Description |
|---|---|
| Single-dose, healthy evaluable immunogenicity | All eligible participants who<br>● Generally healthy without any immunocompromise<br>● received at least one dose of the vaccine,<br>● has proven seronegative or not known to have had previous COVID-19 infection,<br>● with valid and determinate relevant immunogenicity result for the particular analysis<br>● from a blood sampling taking place within the evaluable window for the post-dose 1 acute time-point (no more than 3 days earlier or later than day 21 for arm B or day 28 for arm C, and before dose 2), and<br>● no important protocol deviations as determined by investigators |
| Single-dose healthy expanded immunogenicity | All eligible participants who<br>● Generally healthy without major immunocompromise<br>● Received one dose of the vaccine,<br>● Has proven seronegative or unknown at baseline,<br>● with valid and determinate relevant immunogenicity result for the particular analysis at least 14 days post-dose 1 but before dose 2 |
| Single-dose, all evaluable immunogenicity | All eligible participants who<br>● received at least one dose of the vaccine,<br>● with valid and determinate relevant immunogenicity result for the particular analysis<br>● from a blood sampling taking place within the evaluable window for the post-dose 1 acute time-point (no more than 3 days earlier or later than day 21 for arm B or day 28 for arm C, and before dose 2), and<br>● no important protocol deviations as determined by investigators |
| Single-dose all expanded immunogenicity | All eligible participants who<br>● Received one dose of the vaccine,<br>● With valid and determinate relevant immunogenicity result for |

| | |
|---|---|
| | the particular analysis at least 14 days post-dose 1 but before dose 2 |
| Two-dose, healthy evaluable immunogenicity | All eligible participants who<br>● Generally healthy without any immunocompromise<br>● received two doses of the vaccine within evaluable window for the vaccine (days 21-28 for arm B, days 28-35 for arm C)<br>● has proven seronegative or unknown at baseline, and<br>● with valid and determinate relevant immunogenicity result for the particular analysis<br>● from a blood sampling taking place within the evaluable window for the post-dose 2 acute time-point (within day 14-42 post-dose 2 and before any further doses)<br>● No important protocol deviations as determined by investigators |
| Two-dose healthy expanded immunogenicity | All eligible participants who<br>● Generally healthy without major immunocompromise<br>● Received two doses of the vaccine,<br>● Has proven seronegative or unknown at baseline,<br>● with valid and determinate relevant immunogenicity result for the particular analysis between 7-56 days post-dose 2 |
| Two-dose, all evaluable immunogenicity | All eligible participants who<br>● received two doses of the vaccine within evaluable window for the vaccine (days 21-28 for arm B, days 28-35 for arm C)<br>● with valid and determinate relevant immunogenicity result for the particular analysis<br>● from a blood sampling taking place within the evaluable window for the post-dose 2 acute time-point (within day 14-42 post-dose 2 and before any further doses)<br>● No important protocol deviations as determined by investigators |
| Two-dose all expanded immunogenicity | All eligible participants who<br>● Received two doses of the vaccine,<br>● With valid and determinate relevant immunogenicity result for the particular analysis between 7-56 days post-dose 2 |
| Three-dose, healthy evaluable immunogenicity | All eligible participants who<br>● Generally healthy without any immunocompromise<br>● received three doses of the vaccine; dose 3 at least 84 days after dose 1<br>● has proven seronegative or unknown at baseline, and not known to be infected<br>● with valid and determinate relevant immunogenicity result for the particular analysis<br>● from a blood sampling taking place within the evaluable window for the post-dose 3 acute time-point (within day 13-42 post-dose 3 and before any further doses)<br>● No important protocol deviations as determined by investigators |
| Three-dose healthy | All eligible participants who |

| | |
|---|---|
| expanded immunogenicity | ● Generally healthy without major immunocompromise<br>● Received three doses of the vaccine; dose 3 at least 56 days after dose 1<br>● Has proven seronegative or unknown at baseline, and not known to be infected<br>● with valid and determinate relevant immunogenicity result for the particular analysis between 6-56 days post-dose 3 |
| Three-dose, all evaluable immunogenicity | All eligible participants who<br>● received three doses of the vaccine; dose 3 at least 84 days after dose 1<br>● with valid and determinate relevant immunogenicity result for the particular analysis<br>● from a blood sampling taking place within the evaluable window for the post-dose 3 acute time-point (within day 13-42 post-dose 3 and before any further doses)<br>● No important protocol deviations as determined by investigators |
| Three-dose all expanded immunogenicity | All eligible participants who<br>● Received three doses of the vaccine,<br>● With valid and determinate relevant immunogenicity result for the particular analysis between 6-56 days post-dose 3 |

#### 4.3 Reactogenicity and safety populations

| Population | Description |
|---|---|
| Healthy safety | All eligible participants who<br>● generally healthy without any major immunocompromise<br>● received at least one dose of the vaccine<br>● has proven seronegative or unknown at baseline and not known to be infected<br>● reported any safety or reactogenicity data |
| All safety | All eligible participants who<br>● received at least one dose of the vaccine<br>● reported any safety or reactogenicity data |

### 5 Methodology and hypotheses

#### 5.1 Hypotheses and decision rules

##### 5.1.1 Immunogenicity hypothesis

A primary objective of the study is to prove or disprove the hypothesis that immunogenicity of either vaccines are non-inferior in healthy adolescents/children to that in healthy adults at the acute dose 2 timepoint (around 1 month post-dose 2). We test the hypothesis in two-dose and three-dose healthy evaluable immunogenicity populations for each of the following specified tests.

H0: $\ln(\mu2) - \ln(\mu1) < \ln(0.6)$

$\ln(0.6)$ refers to a non-inferiority margin of 1.67 fold, while $\ln(\mu2)$ and $\ln(\mu1)$ refer to the natural log of the geometric mean of the following immunogenicity assessments performed at the acute dose 2 timepoint from recipients of respective vaccines aged between 5-10 or 11-17 years and 18 or above. If the lower limit of the 95% confidence interval for mean ratio is >= 0.6, the respective non-inferiority objective is met.

1. Anti-S binding antibody level
2. Anti-S-RBD binding antibody level
3. Surrogate neutralizing antibody inhibition level
4. Neutralizing antibody titer
5. Anti-S antibody avidity index
6. Anti-S FcgRIIIa-binding antibody level
7. Total responsive Th1 (to S, and N and M if applicable)
8. Total responsive Tc1 (to S, and N and M if applicable)
9. Anti-N binding antibody level
10. Anti-N-CTD binding antibody level

In addition, we also test a hypothesis that immunogenicity of a single dose of BNT162b2 in adolescents is non-inferior to that of two doses of BNT162b2 in adults.

For healthy children and adolescents receiving intradermal vaccination, we test also a second hypothesis that superior immunogenicity outcomes can be detected in intradermal route over intramuscular route in participants of the same age group, as well as for non-inferiority.

H0: $\ln(\mu2) - \ln(\mu1) <= \ln(1)$

##### 5.1.2 Sample size

When comparing the peak GM immunogenicity outcomes of children with that of adults, or between vaccine types, a sample size of 61 in each group would assure that a two-sided test with alpha=0.05 has 99% power to detect an effect size with a Cohen's d value=0.78, corresponding to 1.67-fold difference on the linear scale, or 0.51 after natural log transformation, between 2 groups and a standard deviation (SD) of 0.65 on the natural log scale within each group. For assays with higher technical requirements such as PRNT, 66 evaluable adolescents/children and 16 evaluable adults tested would achieve 80% power to detect the same non-inferiority margin with the same alpha and SD; lower sample sizes may be applied on practical grounds. The statistical assumptions on effect size referred to the trial design and results by Frenck et al in NEJM (2021) which found a GMR of 1.76 (95% CI 1.47-2.10) for PRNT50 in adolescents to young adults after 2 doses of BNT162b2. For the proportion of participants with a positive result in immunogenicity outcomes or ARs, 110 adolescents would yield a 95% chance to detect the true value within 7.5% of the measured percentage, assuming a prevalence of 80%. Recruitment of 120 participants were targeted per vaccine regimen and age group to accommodate for attrition.

Reference: Frenck RW et al. Safety, Immunogenicity, and Efficacy of the BNT162b2 Covid-19 Vaccine in Adolescents. N Engl J Med (2021); 385:239-250.

##### 5.1.3 Multiplicity consideration

Not considered.

#### 5.2 General methods

Confidence intervals for all endpoints will be 95% (2-sided) unless otherwise noted.

##### 5.2.1 Categorical variables

We describe categorical variables with percentages, numerator (n) and denominator (N), and 95% CI using the Clopper-Pearson method. For differences in binary data between groups, 95% CI will be derived from Fisher exact test.

##### 5.2.2 Continuous variables

We describe continuous variables with case number (n), mean, median, standard deviation, minimum and maximum values.

###### 5.2.2.1 Geometric means

Geometric means and the corresponding 95% CI will be calculated for some immunogenicity outcomes, including neutralizing antibody titers and responsive T cell frequencies.

###### 5.2.2.2 Geometric mean fold rises/changes

Geometric means of a later time point may be divided by that of an earlier time point, and limited to participants with non-missing values at both timepoints.

###### 5.2.2.3 Geometric mean ratios

Geometric mean ratio will be calculated to compare some immunogenicity outcomes between two groups.

###### 5.2.2.4 Geometric mean fold rise ratios

Geometric mean fold rise ratio will be calculated to compare fold change of some immunogenicity outcomes between two groups at two time points.

### 6 Analysis and summary

#### 6.1 Primary analysis

##### 6.1.1 Reactogenicity endpoints

###### 6.1.1.1 Adverse reactions

To satisfy the primary objective of describing the reactogenicity, we analyze reactogenicity data based on the following:

- Estimand: Percentage of participants reporting a particular solicited reaction by maximum severity, in total for all grades
- Analysis set: (healthy) safety population
- Time-point: after each dose on days 0-7
- Statistics: percentage (and 2-sided Clopper-Pearson 95% CI), n and N; each reaction (total) compared across vaccines by Fisher's exact test
- Figure: stacked bar chart showing percentage of participants reporting a particular reaction by maximum severity in each age category and vaccine type

##### 6.1.2 Immunogenicity endpoints

For immunogenicity endpoints, primary analyses of primary endpoints (including non-inferior hypotheses) are conducted in the evaluable immunogenicity populations. Secondary analysis will also be performed in the expanded population.

###### 6.1.2.1 S and S-RBD binding IgG levels

- Estimands: geometric mean value
- Analysis set: 1/2/3-dose evaluable and expanded immunogenicity populations
- Time points: before dose 1, before dose 2, 1 month after dose 2, 2 weeks after dose 3, 6, 18 and 36 months after vaccination
- Statistics: Geometric mean; % with value above LLOD
- Figure: column scatter dot plot with geometric mean and CI95 error bars
- Intercurrent events: Values below the LLOD will be set to 0.5 of the cut-off/limit for analysis.

###### 6.1.2.2 Surrogate neutralizing antibody inhibition levels

- Estimands: geometric mean value
- Analysis set: 1/2/3-dose evaluable and expanded immunogenicity populations
- Time points: before dose 1, before dose 2, 1 month after dose 2, 2 weeks after dose 3, 6, 18 and 36 months after vaccination
- Statistics: Geometric mean; % with value above LLOQ
- Figure: column scatter dot plot with geometric mean and CI95 error bars
- Intercurrent events: Values below the LLOQ will be set to 0.5 of the cut-off/limit for analysis.

###### 6.1.2.3 Neutralizing antibody titers

- Estimands: geometric mean value
- Analysis set: 1/2/3-dose evaluable and expanded immunogenicity populations
- Time points: before dose 1, before dose 2, 1 month after dose 2, 2 weeks after dose 3, 6, 18 and 36 months after vaccination
- Statistics: Geometric mean; % with value above LLOD
- Figure: column scatter dot plot with geometric mean and CI95 error bars
- Intercurrent events: Values below the LLOD will be set to 0.5 of the cut-off/limit for analysis.

###### 6.1.2.4 S binding IgG avidity index

- Estimands: geometric mean value
- Analysis set: 1/2/3-dose evaluable and expanded immunogenicity populations
- Time points: before dose 1, before dose 2, 1 month after dose 2, 2 weeks after dose 3, 6, 18 and 36 months after vaccination
- Statistics: Geometric mean
- Figure: column scatter dot plot with geometric mean and CI95 error bars
- Intercurrent events: N/A.

###### 6.1.2.5 Anti-S FcgRIIIa-binding IgG levels

- Estimands: geometric mean value
- Analysis set: 1/2/3-dose evaluable and expanded immunogenicity populations
- Time points: before dose 1, before dose 2, 1 month after dose 2, 2 weeks after dose 3, 6, 18 and 36 months after vaccination
- Statistics: Geometric mean; % with value above LLOD
- Figure: column scatter dot plot with geometric mean and CI95 error bars
- Intercurrent events: Values below the LLOD will be set to 0.5 of the cut-off/limit for analysis.

###### 6.1.2.6 S (and N and M)-responsive Th1 and Tc1 cell frequencies

- Estimands: geometric mean frequency, geometric mean fold rise
- Analysis set: 1/2/3-dose evaluable and expanded immunogenicity populations
- Time points: before dose 1, before dose 2, 1 month after dose 2, 2 weeks after dose 3, 6, 18 and 36 months after vaccination
- Statistics: Geometric mean; % with value above cut-off
- Figure: column scatter dot plot with geometric mean and CI95 error bars
- Intercurrent events: Values below cut-off will be set to 0.5 of cut-off/limit for analysis

###### 6.1.2.7 GMR of S and S-RBD binding IgG, surrogate neutralizing antibody inhibition level, neutralizing antibody titers, IgG avidity index and FcgRIIIa-binding IgG in participants 11-17 years of age to those 18 years of age or above

- Estimands: geometric mean ratios
- Analysis set: 1/2/3-dose evaluable and expanded immunogenicity populations
- Time points: 1 month after doses 1 and 2, 2 weeks after dose 3, 6, 18 and 36 months after vaccination; including cross-comparison of 1 month after dose 1 in adolescents and 1 month after dose 2 in adults (and 2 weeks after dose 3 in adults and 1 month after dose 2 in adolescents receiving SinoVac)
- Statistics: exponentiated difference in mean and CI95 on natural log scale for geometric means and two-sided CI95 (or one-sided CI97.5). For testing non-inferiority (one month after dose 2)

- Figure: forest plot
- Intercurrent events: Values below the cut-off/lower limit of detection/quantification may be set to 0.5 of the cut-off/limit for analysis if applicable for the assay.

###### 6.1.2.8 GMR of S (and N and M)-responsive Th1 and Tc1 cell frequencies in participants 11-17 years of age to those 18 years of age or above

- Estimands: geometric mean ratio
- Analysis set: 1/2/3-dose evaluable and expanded immunogenicity populations
- Time points: 1 month after doses 1 and 2, 2 weeks after dose 3, 6, 18 and 36 months after vaccination
- Statistics: exponentiated difference in mean and CI95 on natural log scale for geometric means and two-sided CI95 (or one-sided CI97.5). For testing non-inferiority (one month after dose 2)
- Figure: forest plot
- Intercurrent events: Values below the cut-off/lower limit of detection/quantification may be set to 0.5 of the cut-off/limit for analysis if applicable for the assay.

###### 6.1.2.9 GMR of S and S-RBD binding IgG, surrogate neutralizing antibody inhibition level, and neutralizing antibody titers, IgG avidity index and FcgRIIIa-binding IgG in participants 11-17 years of age receiving 2 doses of BNT162b2 to those receiving 2/3 doses of CoronaVac

- Estimands: geometric mean ratio
- Analysis set: 2/3-dose evaluable and expanded immunogenicity populations
- Time points: 1 month after dose 2, 2 weeks after dose 3, 6, 18 and 36 months after vaccination; including cross-comparison of two doses of BNT162b2 and three doses of CoronaVac
- Statistics: exponentiated difference in mean and CI95 on natural log scale for geometric means and two-sided CI95 (or one-sided CI97.5).
- Figure: forest plot
- Intercurrent events: Values below the cut-off/lower limit of detection/quantification may be set to 0.5 of the cut-off/limit for analysis if applicable for the assay.

###### 6.1.2.10 GMR of total S responsive Th1 and Tc1 cell frequencies in participants 11-17 years of age receiving 2 doses of BNT162b2 to those receiving 2/3 doses of CoronaVac

- Estimands: geometric mean ratio
- Analysis set: 2/3-dose evaluable and expanded immunogenicity populations
- Time points: 1 month after dose 2, 2 weeks after dose 3, 6, 18 and 36 months after vaccination; including cross-comparison of two doses of BNT162b2 and three doses of CoronaVac

- Statistics: exponentiated difference in mean and CI95 on natural log scale for geometric means and two-sided CI95 (or one-sided CI97.5).
- Figure: forest plot
- Intercurrent events: Values below the cut-off/lower limit of detection/quantification may be set to 0.5 of the cut-off/limit for analysis if applicable for the assay.

###### 6.1.2.11 GMR of S and S-RBD binding IgG, surrogate neutralizing antibody inhibition level, and neutralizing antibody titers, IgG avidity index and FcgRIIIa-binding IgG in participants receiving intradermal CoronaVac to intramuscular CoronaVac

- Estimands: geometric mean ratio
- Analysis set: 2/3-dose evaluable and expanded immunogenicity populations
- Time points: 1 month after dose 2, pre-dose 3, 2 weeks after dose 3, 6, 18 and 36 months after vaccination
- Statistics: exponentiated difference in mean and CI95 on natural log scale for geometric means and two-sided CI95 (or one-sided CI97.5).
- Figure: forest plot
- Intercurrent events: Values below the cut-off/lower limit of detection/quantification may be set to 0.5 of the cut-off/limit for analysis if applicable for the assay.

###### 6.1.2.12 GMR of total S, N and M responsive Th1 and Tc1 cell frequencies in participants 11-17 years of age receiving intradermal CoronaVac to intramuscular CoronaVac

- Estimands: geometric mean ratio
- Analysis set: 2/3-dose evaluable and expanded immunogenicity populations
- Time points: 1 month after dose 2, pre-dose 3, 2 weeks after dose 3, 6, 18 and 36 months after vaccination
- Statistics: exponentiated difference in mean and CI95 on natural log scale for geometric means and two-sided CI95 (or one-sided CI97.5).
- Figure: forest plot
- Intercurrent events: Values below the cut-off/lower limit of detection/quantification may be set to 0.5 of the cut-off/limit for analysis if applicable for the assay.

#### 6.2 Secondary endpoints

##### 6.2.1 Adverse events

We analyze unsolicited adverse events based on the following:

- Estimand: Percentage of participants reporting an adverse event (by medDRA terms)
- Analysis set: (healthy) safety population

- Time-point: after each dose on days 0-28, before the next dose
- Statistics: incidence (events per person), n and N

##### 6.2.2 Vaccine efficacy prediction

We predict vaccine efficacy for adolescents in each vaccine based on neutralizing antibody correlate of protection, as published by Khoury et al in Nature Medicine, 2021. Geometric mean neutralizing titers in adolescents/children receiving either vaccines will be normalized against that of a set of house convalescent controls as fold of convalescent titers, and protective efficacy against symptomatic disease for either vaccine in adolescents/children by whichever route will be predicted based on the published model.

- Estimands: predicted protective efficacy (against symptomatic disease)
- Analysis set: 1/2/3-dose evaluable immunogenicity populations
- Time points: 1 month after dose 1/2 and 2 weeks after dose 3
- Statistics: Range of prediction will be given by CI95 of GMT and of model estimates.

##### 6.2.3 N and N-CTD binding IgG levels

- Estimands: geometric mean value
- Analysis set: 1/2/3-dose evaluable and expanded immunogenicity populations for SinoVac recipients
- Time points: before dose 1, before dose 2, 1 month after dose 2, 2 weeks after dose 3, 6, 18 and 36 months after vaccination
- Statistics: Geometric mean; % with value above LLOD
- Figure: column scatter dot plot with geometric mean and CI95 error bars
- Intercurrent events: Values below the LLOD will be set to 0.5 of the cut-off/limit for analysis.

##### 6.2.4 GMR of N and N-CTD binding IgG in participants 11-17 years of age to those 18 years of age or above receiving CoronaVac

- Estimands: geometric mean ratios
- Analysis set: 1/2/3-dose evaluable and expanded immunogenicity populations
- Time points: 1 month after doses 1 and 2, 2 weeks after dose 3, 6, 18 and 36 months after vaccination; including cross-comparison of 1 month after dose 1 in adolescents and 1 month after dose 2 in adults and 1 month after dose 2 in adolescents and 2 weeks after dose 3 in adults
- Statistics: exponentiated difference in mean and CI95 on natural log scale for geometric means and two-sided CI95 (or one-sided CI97.5). For testing noninferiority.
- Figure: forest plot
- Intercurrent events: Values below the cut-off/lower limit of detection/quantification may be set to 0.5 of the cut-off/limit for analysis if applicable for the assay.

##### 6.2.5 Correlates of immunogenicity

We explore the correlation between immunogenicity outcomes by Pearson correlation after logarithmic transformation. Participants with valid results for both variables will be included for correlation analysis between any two variables.

- Estimands: Pearson correlation coefficient
- Analysis set: 1/2/3-dose evaluable immunogenicity populations. Subgroup analysis is performed by vaccine type, dose number and age group.
- Time points: pre-dose 1, pre-dose 2 and 1 month after dose 2, 2 weeks after dose 3, 6, 18 and 36 months after vaccination
- Statistics: Pearson correlation after logarithmic transformation
- Figure: correlation matrix

##### 6.2.6 GMR of S and S-RBD binding IgG, surrogate neutralizing antibody inhibition level, and neutralizing antibody titers, IgG avidity index and FcgRIIIa-binding IgG in participants 11 years or above of age living with diseases to those healthy receiving a particular vaccine

- Estimands: geometric mean ratio
- Analysis set: 2/3-dose evaluable and expanded immunogenicity populations
- Time points: 1 month after dose 2, 2 weeks after dose 3, 6, 18 and 36 months after vaccination
- Statistics: exponentiated difference in mean and CI95 on natural log scale for geometric means and two-sided CI95 (or one-sided CI97.5).
- Intercurrent events: Values below the cut-off/lower limit of detection/quantification may be set to 0.5 of the cut-off/limit for analysis if applicable for the assay.

##### 6.2.7 GMR of responsive Th1 and Tc1 cell frequencies in participants 11 years or above of age living with diseases to those healthy receiving a particular vaccine

- Estimands: geometric mean ratio
- Analysis set: 2/3-dose evaluable and expanded immunogenicity populations
- Time points: 1 month after dose 2, 2 weeks after dose 3, 6, 18 and 36 months after vaccination
- Statistics: exponentiated difference in mean and CI95 on natural log scale for geometric means and two-sided CI95 (or one-sided CI97.5).
- Intercurrent events: Values below the cut-off/lower limit of detection/quantification may be set to 0.5 of the cut-off/limit for analysis if applicable for the assay.

#### 6.3 Subgroup analysis

As described elsewhere in this protocol, subgroup analyses include:

- age (18 years and above, 11-17 years, 3-10 years, and below 3)
- healthy history, conditions include:
    - Immune diseases
        - Combined defects
        - Humoral defects
        - Immune dysregulation
        - Innate defects
        - Phagocytic defects
    - Renal disorders
        - On immunosuppression only
        - On dialysis
        - Post-kidney transplant
    - Hematological conditions
        - Post-HSCT
        - Acute lymphoblastic leukemia
    - Allergy
        - To PEG-containing drugs
        - To first dose of COVID-19 vaccine
    - Neuromuscular disorders
        - Immune-mediated
        - Muscular dystrophy on immunosuppression
    - Prior COVID-19

#### 6.4 Baseline and other summaries and analyses

##### 6.4.1 Baseline

###### 6.4.1.1 Demographics

Demographics will be summarized in the enrolled and healthy safety population.

###### 6.4.1.2 Health characteristics

Count for a particular medical history (by MedDRA SOC and PT terms) will be listed when present in the enrolled population. Count and percentage for participants with at least one comorbid condition linked to severe COVID-19 illness will be listed in the enrolled and healthy safety population as well.

##### 6.4.2 Study completion and participant disposition

###### 6.4.2.1 Participant disposition summary

Summary of participant disposition will give the count for participants in age subgroups, for each stage of completion (received doses 1, 2 and 3, attended follow-up clinics, withdrew or excluded, with reason) in the enrolled and healthy safety population.

###### 6.4.2.2 Blood sampling and e-Diary completion

Count and percentage of participants who gave blood samples in and out of pre-specified time windows and submitted reactogenicity and safety data will be given for each timepoint in the enrolled and healthy safety population.

##### 6.4.3 Intervention

###### 6.4.3.1 Vaccination timing and administration

Count of participants who received dosing in and out of pre-specified time windows will be given for each dose in the enrolled and healthy safety population.

##### 6.4.4 Prior/concomitant vaccination and medications

Count of participants who were medicated or vaccinated 28 days before and after each dose will be given for each dose for each prior/concomitant medication/vaccine in the enrolled and healthy safety population.

### 7 Analysis timeline

Dependent on the progress of visits, interim analyses will be planned for different subgroups throughout the study period to report results of different timepoints.

#### 7.1 Interim analysis 1 (Nov 2021)

Interim analysis will be performed first in November 2021 when most enrolled participants with kidney diseases, neurological diseases and hematological disorders or history of HSCT have completed post-dose 2 assessment under the age of 18 years for BioNTech.

The analysis will analyze the available immunogenicity data (S-RBD IgG levels and sVNT) by descriptive statistics without hypothesis testing. Reactogenicity and safety will also be described.

#### 7.2 Interim analysis 2 (Jan 2022)

Interim analysis will be performed in Jan 2022 when 100 healthy participants have completed post-dose 2 assessment under the age of 18 years, including at least 50 for PRNT, for each vaccine.

While paediatric patients with prior COVID-19 and severe paediatric/immune conditions have been enrolled in this trial, the analysis of these participants will be separate. Safety, reactogenicity and immunogenicity analyses will focus on healthy participants.

Key objectives of this analysis are to describe the reactogenicity and safety of these vaccines in the pediatric age group, compare the acute immunogenicity of the vaccines in adolescents and adults, and compare the acute immunogenicity of the two vaccines in adolescents. We aim to demonstrate non-inferiority in a range of immunogenicity endpoints in adolescents to adults.

#### 7.3 Interim analysis 3 (May 2022)

Interim analysis will be performed in May 2022 when more than 60 healthy adolescents receiving CoronaVac and 60 receiving BNT162b2 have completed post-dose 3 assessment under the age of 18 years, including at least 20 for PRNT for each vaccine. Reactogenicity and safety will be analyzed. Antibody and T cell responses against Omicron will also be evaluated.

#### 7.4 Interim analysis 4 (May 2022)

Interim analysis will be performed in May 2022 when most participants with pediatric conditions have completed post-dose 3 assessment under the age of 18 years. Participants with kidney diseases, immune diseases, blood disorders and neurological disorders will be separately analyzed. Immune responses will be analyzed according to diseases subtypes, age, and vaccine regimen. Antibody responses against Omicron will also be evaluated. Reactogenicity and safety will be analyzed.
